# Non-invasive multi-cancer detection using DNA hypomethylation of LINE-1 retrotransposons

**DOI:** 10.1101/2024.01.20.23288905

**Authors:** Marc Michel, Maryam Heidary, Anissa Mechri, Kévin Da Silva, Marine Gorse, Victoria Dixon, Klaus von Grafenstein, Charline Bianchi, Caroline Hego, Aurore Rampanou, Constance Lamy, Maud Kamal, Christophe Le Tourneau, Mathieu Séné, Ivan Bièche, Cecile Reyes, David Gentien, Marc-Henri Stern, Olivier Lantz, Luc Cabel, Jean-Yves Pierga, François-Clément Bidard, Chloé-Agathe Azencott, Charlotte Proudhon

**Author notes:** Equal contribution (Marc Michel, Maryam Heidary).

## Abstract

**Purpose:** The detection of circulating tumor DNA, which allows non-invasive tumor molecular profiling and disease follow-up, promises optimal and individualized management of patients with cancer. However, detecting small fractions of tumor DNA released when the tumor burden is reduced remains a challenge.

**Experimental Design:** We implemented a new highly sensitive strategy to detect base-pair resolution methylation patterns from plasma DNA and assessed the potential of hypomethylation of LINE-1 retrotransposons as a non-invasive multi-cancer detection biomarker. The DIAMOND (Detection of Long Interspersed Nuclear Element Altered Methylation ON plasma DNA) method targets 30-40,000 young L1 scattered throughout the genome, covering about 100,000 CpG sites and is based on a reference-free analysis pipeline.

**Results:** Resulting machine learning-based classifiers showed powerful correct classification rates discriminating healthy and tumor plasmas from 6 types of cancers (colorectal, breast, lung, ovarian, gastric cancers and uveal melanoma including localized stages) in two independent cohorts (AUC = 88% to 100%, N = 747). DIAMOND can also be used to perform copy number alterations (CNA) analysis which improves cancer detection.

**Conclusions:** This should lead to the development of more efficient non-invasive diagnostic tests adapted to all cancer patients, based on the universality of these factors.

**Statement of significance:** The DIAMOND assay is a new highly sensitive strategy to detect base-pair resolution methylation patterns of LINE-1 retrotransposons (L1) from plasma DNA. It targets 30-40,000 young L1 scattered throughout the genome, covering about 100,000 CpG sites and is based on a reference-free analysis pipeline. This provided high coverage data using affordable sequencing depth, which is instrumental to achieve high sensitivity and work with minute amounts of cell-free DNA. Resulting machine learning-based classifiers showed powerful discrimination between healthy and tumor plasmas from 6 types of cancers (colorectal, breast, lung, ovarian, gastric cancers and uveal melanoma including localized stages) in two independent cohorts (AUC = 88% to 100%, N = 747). DIAMOND data can also be used to perform copy number alterations (CNA) analysis which improves cancer detection.

## Introduction

Extensive research has shown that tumor genetic alterations can be detected from plasma DNA of patients with cancer ^1–3^. This paved the way for the use of molecular analyses performed from *liquid biopsies* to genotype tumors non-invasively ^4,5^ and demonstrated the potential of circulating tumor DNA (ctDNA) as a marker of cancer progression ^6,7^. It is also a powerful prognostic factor ^8^ enabling detection of tumor masses not perceptible clinically, after surgery or during treatment. These approaches promise optimal management of cancer patients and are currently playing an important role in oncology ^9,10^. However, several technological obstacles still limit their widespread application. Samples collected at early stages of tumor progression, or during and after treatment, may contain less than one mutant copy per milliliter of plasma ^1,11^.

This is below the detection limit of most used technologies, even when testing multiple genetic alterations simultaneously. Moreover, most methods are biased towards preselected recurrent mutations, which do not cover all tumors. We observed in our previous studies ^12–15^ that approximately 25% of patients affected with breast cancer do not display common mutations trackable in plasma DNA, even at advanced stages. Therefore, it is necessary to develop more sensitive and more informative detection tools.

Multiple studies have demonstrated the central role of epigenetic processes in the onset, progression, and treatment of cancer. Epigenetic alterations (i.e., changes in the pattern of chromatin modifications such as DNA methylation and histone modifications) are promising candidates for cancer detection, diagnosis and prognosis ^16,17^. These *extended* markers provide an additional level of information, overlooked by methods that only question genetic alterations ^18^. Aberrant DNA methylation is a hallmark of neoplastic cells ^16^, which combine hypermethylation of a wide range of tumor suppressor genes along with a global hypomethylation of the genome^19^. DNA methylation is a stable modification, which affects a large number of CpG sites per region and per genome and will be key to achieve increased detection sensitivity ^20^. Moreover, the concordance of the methylation status between multiple CpGs of the same region can help detect low frequency anomalies among a heterogeneous population of molecules ^21,22^. Finally, combining several genomic regions allows to capture a wide range of tumor alleles and cover the heterogeneous profiles of cancer patients ^23^.

Previous studies have shown that cellular DNA methylation patterns are conserved in cell-free DNA (cfDNA) and that detection of cancer-specific profiles at the genome-wide scale is feasible ^24–27^. Until now, most studies investigating plasma DNA methylation patterns have targeted a limited number of regions at high depth, using PCR-based methods ^28–30^, or explored genome-wide at low depth with high-throughput sequencing ^24–26,31^. Both approaches have limited sensitivity, as focusing on a few regions does not cover cancer-type and patient variability and low depth cannot detect small fractions of ctDNA. More recent studies, relying on the capture of regions of interest coupled with deep sequencing have investigated the performance of larger numbers of regions at high depth ^21,32–40^. These methods enabled sensitive detection and classification of cancer from plasma DNA. However, since they largely focus on cancer hypermethylation and unique sequences, it involves targeting specific regions for each cancer type. As a result, developing a cost-effective universal pan-cancer test remains a challenge.

Remarkably, cancer-related hypomethylation has been reported in almost all classes of repeated sequences ^41^, from dispersed retrotransposons to clustered satellite repeated DNA, and within multiple forms of cancers ^42^. In particular, this leads to the reactivation of retrotransposons, resulting in the acquisition of genomic instability, chromosomal rearrangements and the production of chimeric transcripts between the transposable element and its adjacent locus.

Hypomethylation of the internal promoter of Long-Interspersed Element-1 (L1) has been described as a hallmark of many human cancers ^42,43^, which can result in the reactivation of intact L1 elements ^44^ and the abnormal production of their transcripts and proteins. Transposition of these competent elements induces DNA double-strand breaks and damages the genome. A recent study identified 4 types of cancer (esophagus, head and neck, lung and colorectal) with a large amount of damage linked to retrotranspositions involving mostly L1s ^45^. Another study identified the transposition event responsible for initiating colorectal cancer by mutating the APC gene ^46^. To obtain a global representation of the hypomethylation occurring during carcinogenesis and to increase sensitivity, we chose to target primate-specific copies of L1 retrotransposons (L1PA). These elements have tens of thousands of copies per cell and are hypomethylated in multiple cancers ^42^. Two studies have explored L1 global methylation profiles from plasma ^47,48^ of lung and colorectal cancers, using qPCR-based methods, but reported a low detection sensitivity, below 70%. Indeed, repeats being inherently difficult to map, detecting their methylation profiles at the single base-pair resolution requires sophisticated downstream analysis.

To overcome this, we have developed a method to detect methylation patterns of primate specific L1 elements (L1PA) from cfDNA, which we named DIAMOND (for **D**etection of Long **I**nterspersed Nuclear Element **A**ltered **M**ethylation **ON** plasma **D**NA). We implemented computational tools to accurately align sequencing data without a reference genome and applied prediction models, trained by machine learning algorithms, integrating patterns of methylation, overall and at the single molecule level. The aim of this study was to assess the potential of circulating DNA methylation changes at L1s as a universal tumor biomarker, and to develop new highly sensitive strategies to detect cancer-specific signatures in blood.

## Results

### Targeting primate-specific LINE-1 elements reveals plasma DNA-methylation patterns genome-wide

We developed a PCR-based targeted bisulfite method coupled to deep sequencing to detect methylation patterns of L1PA elements. We used sodium bisulfite chemical conversion to achieve base-pair resolution analysis and designed a multiplexed PCR based on 8 amplicons covering L1PAs (**Fig. 1A, Table S1, Fig. S1A**). We detected thousands of L1PA elements scattered throughout the genome as shown by the genomic hits obtained from a healthy plasma, an ovarian tumor, and a uveal melanoma tumor sequenced at high depth (**Fig. 1B, Table S2**). We observed similar profiles for the three samples, as well as for healthy and cancer plasmas with standard coverage (**Fig. S1B-E**). This demonstrated the robustness of the approach. Overall, the estimated number of L1PA targets is about 30-40,000 elements per genome including half of the human specific copies (L1HS) and many copies of the other L1PA subfamilies (**Fig. 1C, Table S2**). This represents an estimate of 87-120,000 CpG sites.

**Fig 1.**
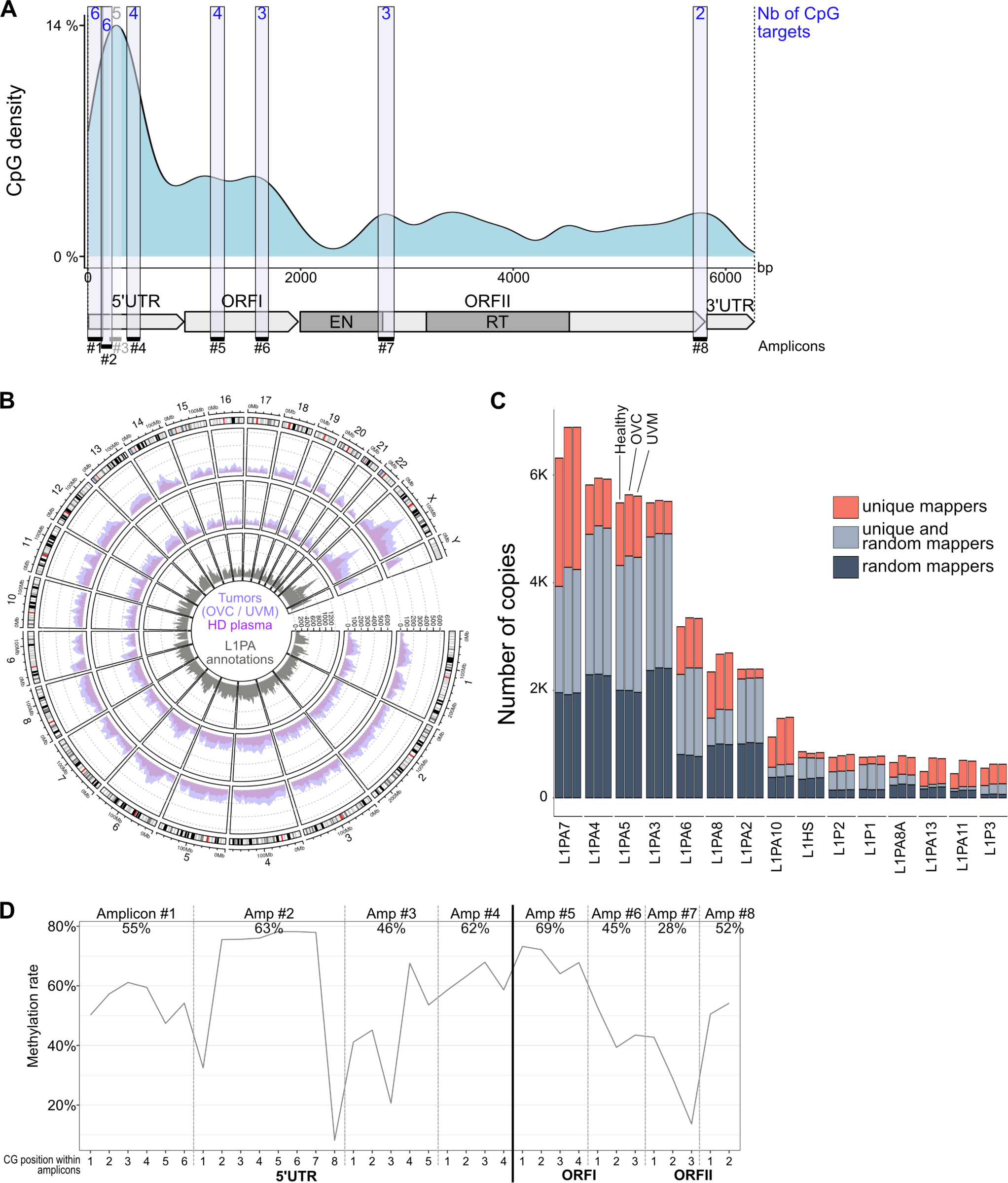
Targeting primate-specific LINE-1 elements reveals plasma DNA-methylation patterns genome-wide. **A.** CpG density along the structure of a human specific LINE-1 (L1HS) element, which contains 95 CpG. The DIAMOND assay targets 30 CpG. Each target amplicon is highlighted by a black bar below the structure. The number of CpG sites detected per amplicon is displayed in blue. **B.** L1PA copy number hit by uniquely and/or randomly mapped reads, obtained from a healthy plasma versus ovarian (OVC, top track) or uveal melanoma (UVM, middle track) tumor tissue samples ‘deep sequenced’ (54M, 44M or 46M reads respectively) over the distribution of L1PA elements annotated in the genome (RepeatMasker on hg38, grey bottom track). **C.** Histogram summarizing the most represented sub-families of L1 targeted by the DIAMOND assay in the 3 ‘deep sequenced’ samples, in descending order (sum of copies across the 3 samples). The colors highlight the relative contribution of L1PA copies hit by reads uniquely mapped, randomly mapped or both. **D.** Methylation pattern observed across the 8 regions targeted along the L1 element in the healthy plasma sample ‘deep-sequenced’. Metaplot showing the average methylation levels at each CpG position. Amplicon limits are delineated with grey dotted lines. The dark line marks the end of the 5’UTR. Average levels per amplicon are indicated.

Following deep sequencing, reads are traditionally mapped back to the genome. However, the majority of sequencing reads from repetitive sequences are assigned randomly during mapping steps and are subsequently lost for classical differentially methylated region (DMR) calling ^49^. We, thus, developed a new computational pipeline to accurately align repetitive sequencing data without using a reference genome (**Fig. S1F**). To perform this, we clustered all good quality reads based on their similarity, extracted representative sequences from the largest clusters and used them for multiple sequence alignment. We then aligned all the reads back onto this custom database. Using such reference-free method, we preserved the majority of our data and could extract the informative CpG sites agnostically. We selected sites with a CG/TG content ≥ 20% including at least 5% of CG to ensure that the position of interest carries some DNA methylation marks. This selection was done on healthy samples to avoid biases related to cancer hypomethylation. We retrieved 35 CpG positions covered by our panel including two additional CpGs with respect to the L1HS consensus annotations, located within amplicon 2 (**Fig. S2A-B**).

As expected, the 5’ end of the L1 copies targeted is heavily methylated ^42,50^, particularly within the 2^nd^ amplicon. We also observed quite high levels in both the 5^th^ amplicon (69% in average, **Fig. 1D**), which covers part of the ORFI, and the last two CpGs of amplicon 8, which is located immediately upstream of the 3’UTR. Amplicon 3, which has the lowest methylation levels within the 5’ end, displayed sequencing data with atypical distributions and showed less robust performances (not shown). Hence, we further eliminated it from the rest of the study, resulting in a total of 30 CpG positions analyzed. Overall, this reference-free method retrieved methylated sites contained by the youngest LINE-1 elements present in the human genome allowing us to study their DNA-methylation levels and motifs from minute amount of DNA such as plasma cfDNA.

### L1PA hypomethylation is detectable from plasma DNA in multiple forms of cancer

We first tested the DIAMOND approach on methylation controls, cancer cell lines and tissue samples. The overall methylation levels demonstrated an extensive L1PA hypomethylation specifically in cancer samples, including colorectal (CRC), ovarian (OVC), breast (BRC) and uveal melanoma (UVM) cancer cell lines as well as OVC, BRC and UVM tumors compared to healthy white blood cells and healthy tissues collected adjacent to ovarian tumors (**Fig. 2A, Table S3**). Next, we tested a cohort of 473 plasma samples including 123 healthy plasma controls and plasma samples from patients with 6 different types of cancer, covering metastatic (M+) and localized (M0) stages (**Table S4**. This includes colorectal and ovarian cancers in which a substantial rate of L1 hypomethylation has previously been reported ^51,52^. We detected a statistically significant L1PA hypomethylation in cfDNA of metastatic colorectal cancer (CRC M+), breast cancer (BRC M+) and uveal melanoma (UVM M+) samples as well as in locally advanced ovarian cancers (OVC M0, stages III) and localized gastric cancers (GAC M0) (**Fig. 2B, Table S3**). The global methylation was not significantly different in metastatic non-small cell lung cancers (LC M+) nor in localized stages of breast cancer (BRC M0). However, focusing strictly on global methylation levels provides only part of the information.

**Fig 2.**
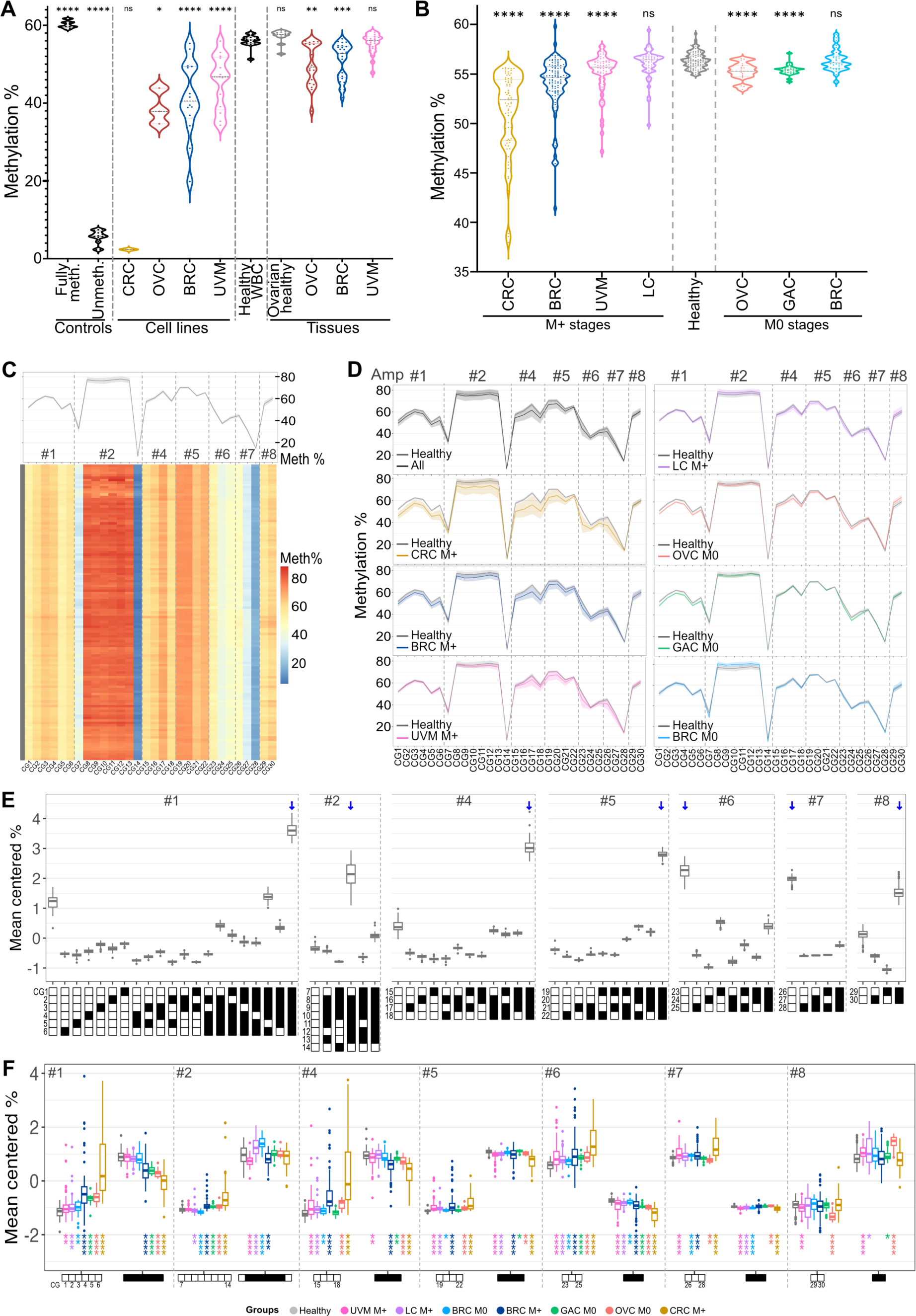
L1PA hypomethylation is detectable from plasma DNA in multiple forms of cancer. **A.** Global DNA methylation of fully methylated (healthy white blood cells (WBC) DNA treated with SssI) and unmethylated (whole-genome amplified healthy WBC DNA) controls, cancer cell lines or tissues. Ovarian healthy tissues were collected next to ovarian tumors. The global methylation levels for each sample correspond to the percentage of CG dinucleotides at each CpG site averaged by the number of CpG sites. Statistical differences between controls, cell lines or tissues and healthy white blood cells (WBC) were computed using Mann–Whitney *U* test (p_Fully_meth._ = 9.97e-07, p_Unmeth._ = 1.86e-06, p_CRC_Cells_ = 0.266, p_OVC_Cells_ = 1.20e-02, p_BRC_Cells_ = 2.47e-06, p_UVM_Cells_ = 6.77e-05, p_Healthy_OVC_Tissues_ = 0.063, p_OVC_Tissues_ = 8e-03, p_BRC_Tissues_ = 4.10e-04, p_UVM_Tissues_ = 0.88, **Table S3**). **B.** Global DNA methylation in cancer plasma including metastatic stages (M+) and non-metastatic stages (M0) as well as healthy donor plasmas. Statistical differences between each cancer subgroup and healthy samples were computed using Mann–Whitney *U* test (p_CRC_M+_ = 1.27e-29, p_BRC_M+_ = 3.79e-19, p_UVM_M+_ = 8.29e-06, p_LC_M+_ = 0.655, p_OVC_M0_ = 1.94e-05, p_GAC_M0_ = 4.28e-08, p_BRC_M0_ = 9.10e-01, **Table S3**). Black dotted lines represent the median. **C.** Methylation level at each targeted CpG sites (x-axis), for each healthy sample (y-axis) depicted as a heatmap. CpG numbers are indicated. The metaplot represents the average methylation levels of the population. Amplicon numbers are indicated. **D.** Differential methylation levels between healthy samples and patients for each type of cancer represented as metaplots. **E.** Proportion of methylation motifs, called haplotypes, for each amplicon (mean centered per amplicon). Only the most important features are represented (see Fig. 3F and methods). Blue arrows highlight the most abundant haplotype in each amplicon. **F.** Mean centered abundance of the most important haplotypes with the highest co-methylation patterns (mostly fully methylated or fully unmethylated molecules) in cancer subgroups compared to healthy donors. Statistical significances were computed using Mann–Whitney *U* test on raw haplotype proportions (**Table S5**).

We further computed the levels of methylation at each CpG target (*n*=30) for these plasma samples and observed specific patterns of methylation along the L1 structure, which are robustly conserved among the 123 healthy donors (**Fig. 2C**). When considering all cancer samples together, we observed a clear difference with the methylation of healthy samples. We detected a steady hypomethylation through all CpG targets except for the two sites within amplicon 8 (**Fig. 2D**). This is also true for metastatic colorectal cancers (CRC M+), breast cancers (BRC M+) and uveal melanoma (UVM M+). Clear hypomethylation is also observable for localized gastric (GAC M0) and ovarian (OVC M0) cancers, in particular at amplicon #1, #4 and #6, while the differences are less striking for localized breast cancers (BRC M0) and metastatic non-small cell lung cancers (LC M+). The distinction between most cancers and healthy samples were dependent on multiple CpG positions belonging to different amplicons along L1s, as shown by PCA analysis (**Fig. S2C**). The least discriminating positions were located within amplicon 8, which is consistent with the metaplots shown in **Fig. 2D**.

Next, we analyzed the motifs of methylation at the molecule level, which provide a more detailed signal. These *haplotypes* correspond to true patterns of methylation of adjacent CpGs, detected for each amplified DNA molecule. This was achieved by the incorporation of unique molecular identifiers (UMIs) into the library (**Fig. S1A**). Based on the combination of the 30 CpG targets divided into their 7 amplicons, we extracted a total of 372 unique features (**Fig. S2D**). We observed highly robust representation profiles of haplotypes among the 123 healthy samples (**Fig. 2E**). For most amplicons, the fully methylated molecules were the most represented, as expected for healthy controls. However, we observed a high proportion of totally unmethylated haplotypes in amplicon #6 and #7. This can be explained by the fact that older L1 copies are often truncated in 5’ and less regulated by DNA methylation, leading to the capture of molecules with lower DNA methylation in 3’. Nevertheless, several intermediate patterns were also among the most important features and were found to be differentially represented in healthy and cancer samples (**Table S5**). Fully methylated haplotypes were significantly under-represented in most cancer subgroups and in most amplicons (**Fig. 2F**). On the contrary, fully unmethylated haplotypes were over-represented in most cancer subgroups and in most amplicons. This is also well illustrated by the PCA analysis shown in **Fig. S2E**, underlining the contribution of highly methylated haplotypes towards the healthy group versus the lowly methylated haplotypes separating cancer samples (middle panel). This separation involves haplotypes from all amplicons (right panel).

Next, we compared the methylation profiles of tumor and plasma paired samples (OVC = 10, **Fig. S3A-F**; BRC = 16, **Fig. S3G-K**) by calculating the correlation between their methylation differences relative to the mean methylation of healthy donor plasmas. We observed a better correlation with methylation haplotype portions than with single CG methylation features (**Fig. S3B-C** and **Fig. S3H-I, Table S6**). These results demonstrate that L1 hypomethylation can robustly be observed from cancer plasma DNA at the level of single CpG sites and more importantly at the level of methylation haplotypes.

### L1PA hypomethylation-based classifiers recognize samples from multiple forms of cancer

We then trained classification models using random forests, with the 30 features corresponding to the levels of methylation at each CpG target or the 372 features corresponding to the proportions of haplotypes, or both, and assessed their performances to automatically separate healthy from tumor plasmas. By testing all cancer samples without cancer-type specification, the methylation of L1PA elements showed an extremely good ability to discriminate between healthy and tumor plasmas, with an overall area under the curve (AUC) of 94-95% for the 3 types of features (**Fig. 3A, 3C, S4A-B**). Next, we trained distinct models to estimate the performances for each cancer type and/or dissemination stage (M0 vs M+). These models were extremely performant in metastatic colorectal and breast cancers but also stage III ovarian cancers and localized gastric cancers, with nearly perfect classifications and AUCs between 98-100% (**Fig. 3B-C, S4A-B**). Additionally, we observed excellent performances for metastatic lung cancers and uveal melanoma and more importantly for localized stages of breast cancer (AUC_BRC_M0_ = 92-95%). These models provide very good sensitivities at 99% specificity (**Fig. 3D**), in particular for CRC M+, BRC M+, OVC M0, GAC M0 and BRC M0. The latter is one of the most difficult cancer to detect non invasively, as reported in previous liquid biopsy multi-cancer tests ^11,39,40^.

**Fig 3.**
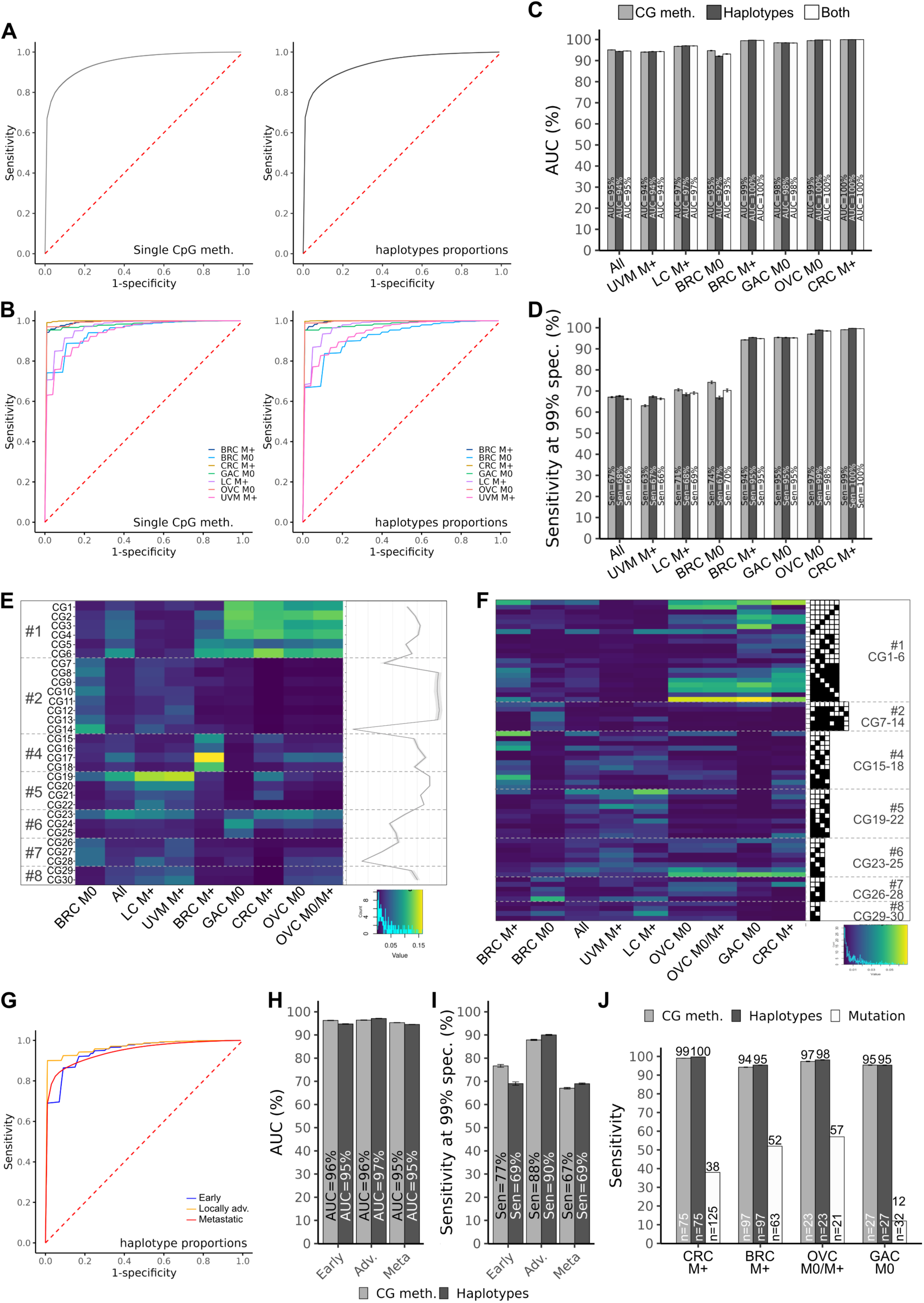
L1PA hypomethylation-based classifiers recognize samples from multiple forms of cancers. **A-B.** Receiver Operating Characteristic (ROC) curves obtained for plasma samples classification using single-CpG methylation levels (n=30) or haplotype proportions (n=372) with the ‘all cancers’ model (**A**) or the ‘cancer-types’ models (**B**). All classifications include 5000 stratified random repetitions of learning on 60% of the samples and testing on the 40% left, with undersampling for classes equilibrium (results with and without undersampling are presented in **Fig. S3A-B**). N_CRC_M+_ = 75, N_BRC_M+_ = 97, N_LC_M+_ = 50, N_UVM_M+_ = 55, N_OVC_M+_ = 4 (included only in ‘all cancers’ testing), N_OVC_M0_ = 18, N_GAC_M0_ = 27, N_BRC_M0_ = 23 tested versus 123 healthy donors. ROC curves shown are obtained by averaging the sensitivity and specificity of each repetition of learning. **C-D.** Performances for classifiers using single CpG methylation levels (grey), haplotype proportions (black) or both (white) presented as AUCs (**C**) or sensitivities at 99% specificity (**D**). Average AUCs are computed from the 5000 AUCs generated by each repetition of learning. Bars indicate 95% CI. **E-F.** Importance (mean decrease in impurity) of the features used by the classifiers depicted as clustered heatmaps. The features correspond to the CpG targets (**E**) or the haplotypes (**F**). Only the most important haplotypes (feature importance level >1%) are shown. **G.** ROC curves obtained for plasma samples classification with the 3-stage model, using haplotype features. **H-I.** Performances for the 3-stage classifiers using single CpG methylation levels (grey) or haplotype proportions (black) presented as AUCs (**H**) or sensitivities at 99% specificity (**I**). Early stages (I/II, N=31), locally advanced stages (III, N=30) and metastatic stages (IV, N=281) **J.** Cancer detection rates with the methylation-based DIAMOND assay (haplotypes and CG methylation) vs common recurrent mutations for samples assessed in previous studies (^13,14,55,56^) or with NGS (**Table S6**).

Overall, we observed similar results using single-CpG methylation levels, haplotype proportions or both features. This can be explained by the high correlation observed between the 2 types of features (**Fig. S4C**). We compared these performances for each subgroup with the classification rates extracted from the model ‘all’, testing all cancer samples together. We observed similar AUCs for most cancers, but overall, they were better classified with their ‘expert’ cancer specific models (**Fig. S4D-E**). This shows that certain specificities of cancer type may confuse the current ‘all’ model and affect the sensitivities at 99% specificities.

Subsequently, we evaluated the importance of the features used by our classifiers (**Fig. 3E-F**). CpG positions displayed different patterns in the various cancer subgroups that can be informative for distinct cancer types or stages (**Fig. 3E)**. Nonetheless, we identified features which are common to many types of cancer such as most CpGs of amplicon 1 and the first CpG of amplicon 6. Other features seemed to be characteristic of specific subgroups, such as CG7-14 which are the most important features for sorting localized stages of breast cancer (BRC M0) or CG15-18, in particular CG17, which are part of the top features for metastatic breast cancers (BRC M+). Besides their dissemination status, the different important features detected in the BRC M0 and M+ subgroups may result from the fact that they were composed of different breast cancer subtypes, with 100% of hormone dependent (HR+ HER2-) cancers in the BRC M+ subgroup and a mixture of subtypes, including also triple negative (TNBC) and HER2+ cancers, in the BRC M0 subgroup (**Table S7)**. Indeed, we observed that slightly different hypomethylation patterns in HR+ HER2-BRC versus TNBC (**Fig. S4F-H**). We observed that haplotype features showed consistent patterns with important CG features (**Fig. 3F**). However, haplotypes provide a more detailed view of the methylation patterns with a strong importance of the most methylated or non-methylated molecules. We still observed that some methylation intermediates are important for cancer detection (ex: in amplicon #1 in CRC M+ and GAC M0, #2 in BRC M0, #4 in BRC M+ and other subgroups, #5 in LC M+ and UVM M+, #7 in OVC, #8 in BRC M0 and LC M+). BRC M+ and M0 subgroups cluster together, showing that even if they are not highly similar, they are closer to each other than to other cancer types. These breast cancer specificities are worth exploring further in the future. Overall, this suggests that L1PA methylation alterations detected from cfDNA vary in different types and stages of cancer.

To estimate the ability of DIAMOND to detect cancer at early stages of the disease, we build classifiers for 3 stage classes gathering all cancer types: early stages (I/II, N=31), locally advanced stages (III, N=30) and metastatic stages (IV, N=281). Classifications were highly performant for all 3 stage categories (AUC_Early_ = 95%, AUC_Adv._ = 97%, AUC_Meta_ = 95%; **Fig. 3G-H, S4I, S4D**) with a mean sensitivity of 70% for early stages, (Sen_Early_ = 70%, Sen_Adv._ = 90%, Sen_Meta_ = 69%; **Fig. 3I, S4E**).

Next, we analyzed whole genome bisulfite sequencing (WGBS) from healthy and cancer plasma from two recently published studies ^53,54^ to evaluate if we can retrieve L1PA hypomethylation signal with non-targeted methods (**Fig. S5A**, see methods). Consistent with our findings, we observed statistically significant hypomethylation in cancer samples (**Fig. S5B-H**). These whole genome approaches were nevertheless not as efficient as the DIAMOND targeted approach to automatically detect cancer from L1PA hypomethylation as they cover less well the regions of interest (**Fig. S5A, S5F, S5I**). Indeed, they involve mapping on a reference genome leading to loss of data. Moreover, L1PA methylation measured with DIAMOND largely outperforms methods based on the detection of mutations. In comparison, the identification of the same tumor samples via the detection of frequent recurrent mutations, which is commonly used in the clinic, does not exceed 57% for ovarian cancer (**Table S8**), 38% for colon cancer ^55^ and 52% for metastatic breast cancer ^13,14^ (**Fig. 3J**). We particularly achieved remarkable performance on the cohort of 27 localized gastric cancers with a detection rate of 95% of true positive as compared to 12% for mutation screening ^56^. This is mostly due to the fact that methylation changes occur in virtually all cancer patients, unlike recurrent mutations.

### Multi-cancer classification performances are reproducible on an independent cohort

To validate the DIAMOND approach, we tested a second independent cohort consisting of 214 patients affected with the same types of cancers as in the first cohort, excluding uveal melanoma and non-metastatic gastric cancers, along with 60 healthy donors (**Fig. 4A**). First, we confirmed that the methylation patterns along the L1 structure were highly reproducible between healthy donors from cohorts 1 and 2, at the level of single-CpG targets (**Fig. 4B**) but also for haplotype proportions (**Fig. 4C, Table S9**). While methylation at single-CpG within cancer subgroups showed slightly more variability (**Fig. S6A**), global methylation levels were quite reproducible between the two cohorts, showing similar distributions and no statistical differences (**Fig. 4D, Table S10**), except for non-metastatic ovarian cancers. There was an important heterogeneity among the OVC M0 samples of cohort 2, which clustered into two distinct groups, while cohort 1 was more homogeneous (**Fig. S6B**). Notably, no correlation was found with available clinico-histopathological parameters (age, staging, CA125 level, mutational status, treatment or response to therapy). Differential haplotype proportions between healthy and cancer subgroups were also mostly conserved (**Fig. S6C, Table S11**). Overall, the method showed good reliability with the 7-amplicon panel used and good robustness in detecting L1 methylation levels and changes.

**Fig 4.**
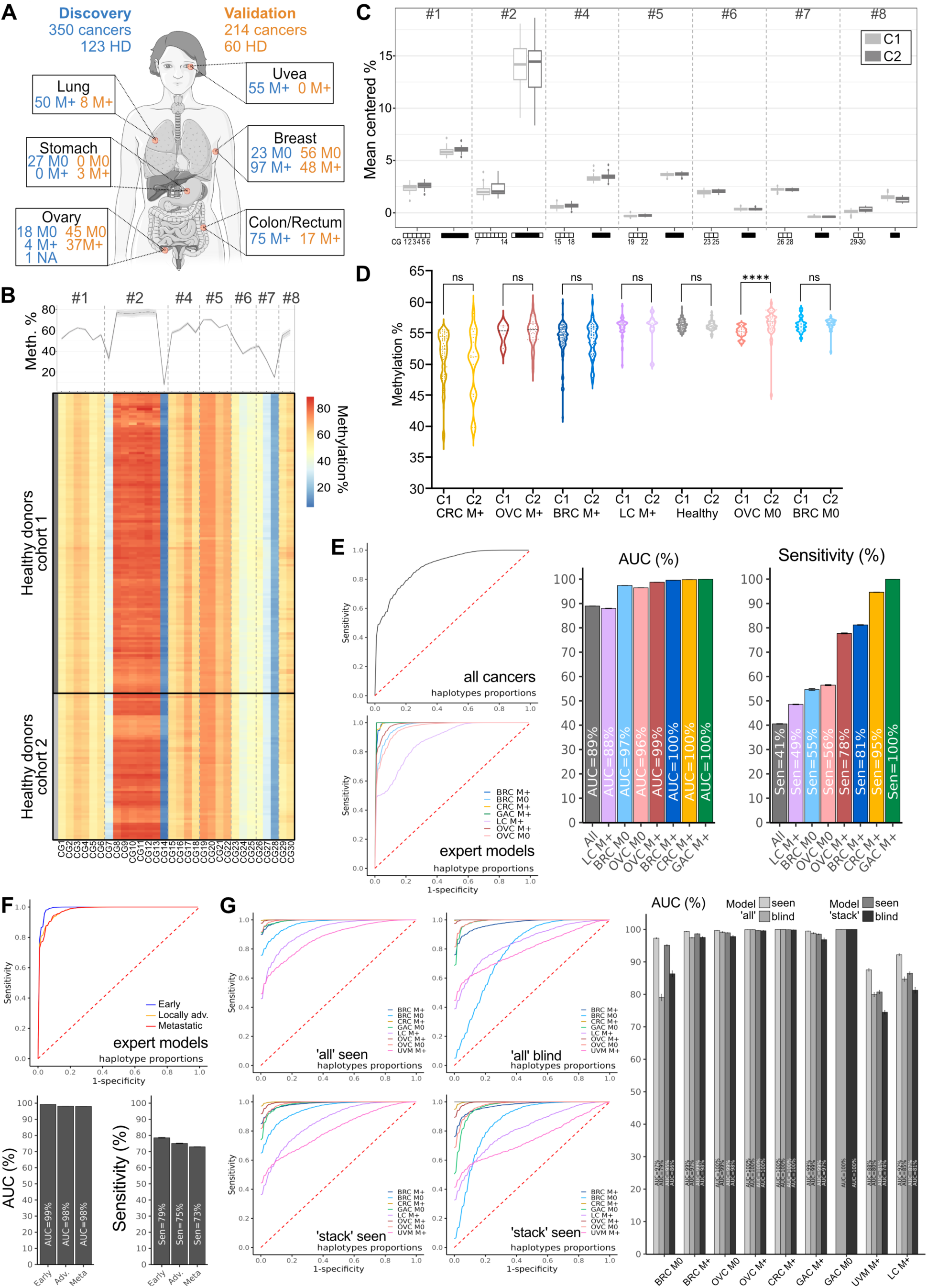
Multi-cancer classification performances are reproducible on an independent cohort. **A.** Number of patients and healthy donors (HD) in the discovery cohort (C1) and in the validation cohort (C2) for each cancer type and dissemination stage (non-metastatic: M0 vs metastatic: M+, NA: stage not available). Generated using Servier Medical Art. **B.** Methylation level at each targeted CpG sites (x-axis), for each healthy sample (y-axis) from C1 vs C2, depicted as a heatmap. No clustering is done on the data, which comes ordered by targeted CpG site on the x-axis (amplicon numbers are indicated). The metaplots represent the average levels for donors of C1 versus C2 at each CpG site. **C.** Mean centered abundance of the most important haplotypes, with the highest co-methylation patterns, in healthy donors from C1 vs C2 (Statistical differences computed using Mann–Whitney *U* test are available in **Table S9**). **D.** Comparison of the global levels of methylation in C1 vs C2. Methylation levels are calculated as explained previously in Fig. 2. The p-values are computed using Mann–Whitney *U* test (p_CRC_M+_ = 0.680, p_OVC_M+_ = 0.816, p_BRC_M+_ = 0.783, p_LC_M+_ = 0.596, p_Healthy_ = 0.316, p_OVC_M0_ = 4.74e-05, p_BRC_M0_ = 0.132, **Table S10**). Black doted lines represent the median. **E.** Performances for validation classifiers using haplotype features presented as ROC curves, AUCs and sensitivities at 99% specificity obtained with the ‘all’ cancers model or the ‘expert’ models for cancer subgroups. All classifications include 5000 stratified random repetitions of learning on the whole discovery cohort and testing on the whole validation cohort without undersampling. ROC curves shown are obtained by averaging the sensitivity and specificity of each repetition of learning. Average AUCs are computed from the 5000 AUCs generated by each repetition of learning. Bars indicate 95% CI. **F.** Performances for 3-stage ‘expert’ classifiers: early stages (I/II, N_C1_=31, N_C2_=38), locally advanced stages (III, N_C1_=30, N_C2_=54) and metastatic stages (IV, N_C1_=281, N_C2_=113) presented as mean ROC curves, AUCs, or sensitivities at 99% specificity. **G**. Performances for integrated models (‘all’ or ‘stack’) when training for the specific group tested (seen) versus when not training for this subgroup (blind). These classifications have been performed on the whole sample set including C1 and C2.

Since age-related changes in DNA methylation have been described ^57,58^ and that the healthy donors included in the study are younger overall than the cancer patients (**Fig. S7A**), we have investigated whether there was an effect on the methylation patterns we studied. We found a significant but very small effect which appeared much smaller that the effect of disease status (**Fig. S7B-C**). This small effect was tending towards an increase in methylation with age (**Fig. S7D**), and we observed similar patterns and differences between healthy and cancer samples when adjusting for the age (**Fig. S7E-G, Table S12**). Furthermore, we observed similar performances in age-matched and non-age-matched cohorts extracted from C2 (**Fig. S7H-J**), demonstrating that age is not a confounding factor.

To validate our classifiers, we trained models on the entire first cohort and evaluated them on the second set of independent samples. The results showed excellent classification performances with an overall AUC of 88% when testing all cancers together with no annotations of their histological types and AUC between 88%-100% for the cancer subgroup ‘expert’ models with sensitivities at 99% specificity between 49%-100% (**Fig. 4E**) including 55% for localized breast cancers. It was, however, lower for metastatic lung cancer with a mean sensitivity of 49%. We observed that haplotype models were more robust compared to single-CpG methylation rates (**Fig. S6D-G**). This could be explained by the fact that haplotypes capture true methylation patterns at the molecule level, enabling to discard noise caused by experimental variability for example. Next, we applied the same validation method, training on C1 and testing on C2, for the 3-stage ‘expert’ classifiers and observed great classification performances with a mean AUC of 99% and a mean sensitivity of 79% for early stages (**Fig. 4F**). When comparing the performances for each subgroup with the classification rates extracted from the model ‘all’, we observed similar AUCs for most cancers, but lower rates for the BRC M0 subgroup and the early-stage group (**Fig. S8A**), which is mostly composed of BRC M0 (**Fig. S8B**).

To further investigate the generalization of our marker, we have also tested the ability of DIAMOND to detect cancers of a type or subgroup for which the model is not trained. To do so, we have combined the data from cohorts C1 and C2 and trained on the whole data set, systematically removing one cancer subgroup or one cancer type and testing specifically for this subgroup or type (see Methods, **Fig. 4G, S8C**). We observed performances similar to when the subgroup is included in the train set, which demonstrates the universality of L1PA hypomethylation. Classification performance is noticeably lower for the BRC M0 subgroup when the model still performs well for OVC M0 and GAC M0. This may be because the BRC M0 subgroup contains the earliest stages of both cohorts (**Fig. S8B**) and the model is less well trained for these stages when removing it. To improve this, we have developed another integrated model that is trained for all cancer types and stages together but also incorporates the probabilities from the ‘expert’ models, defining profiles for various stages and dissemination status. This ‘stack’ model provides a unique prediction for cancer versus healthy status for each sample at once, independently of their cancer type / dissemination status, and allows to balance for the cancer characteristics used to train the model by including them all. We have then extracted the performances per cancer subgroups out of this integrated model, which reached better performances to classify BRC M0 and early-stages samples (see Methods, **Fig. 4G, S8A**).

This demonstrates the robustness of cancer detection by probing L1PA hypomethylation from plasma DNA with the DIAMOND assay and its ability to detect early stages. Finally, we evaluated if the extent of L1PA hypomethylation was associated with survival. In the validation cohort, we compared patients with high methylation to those with low methylation levels (above or below the median threshold - **Fig. S8D**) and observed that more pronounced hypomethylation is clearly associated with shorter survival (**Fig. S8E**). This effect is thought to mainly reflect the tumor burden, which directly impacts the fraction of tumor DNA circulating in the blood, but it remains an interesting non-invasive tumor marker.

### DIAMOND data contain signal to infer the tumor burden, which improves cancer detection

When looking at the overall methylation rates, we detected significantly more hypomethylation for more advanced stages of the disease, in particular in metastatic stages compared to localized stages (**Fig. 5A, Table S13**). However, there was no significant differences between metastatic tumor tissues and primary tissues (**Fig. 5B, Table S14**), which confirmed that L1PA methylation alteration is an early event in carcinogenesis ^59,60^ and also affects early-stage cancers. The differences observed in the blood (**Fig. 5A**) reflect the ctDNA fraction, which is known to be directly influenced by the tumor burden and the stage of the disease ^1,9^. This demonstrates the quantitative potential of DIAMOND, which could help quantify the tumor burden and monitor the disease.

**Fig 5.**
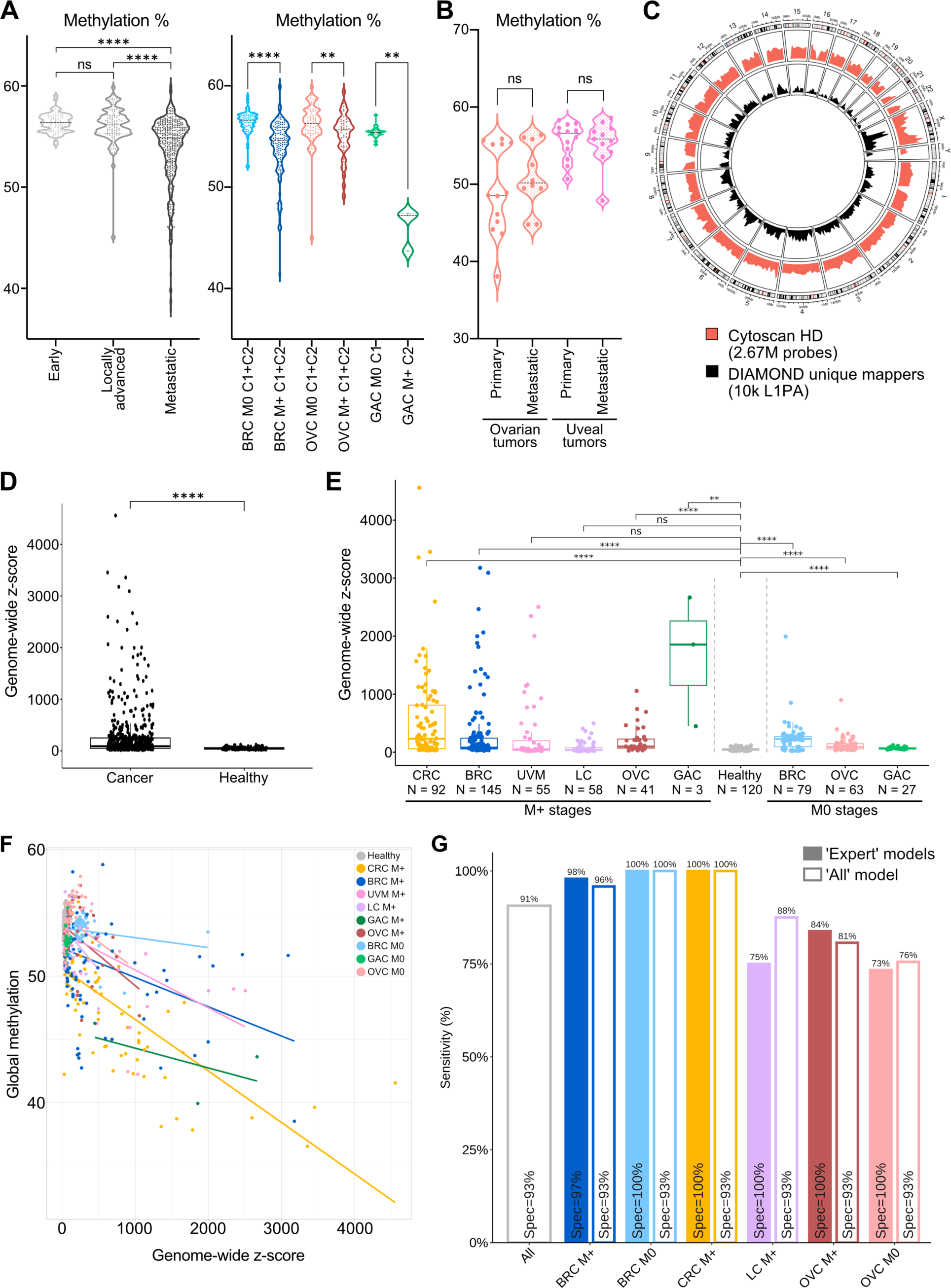
DIAMOND data contain signal to infer the tumor burden, which improves cancer detection. **A-B.** Comparison of the average levels of methylation observed in localized vs metastatic plasma samples (**A**, p_Early/Adv._ = 0.327, p_Adv./Meta._ = 3.14e-11, p_Early/Meta._ = 2.82e-14; p_BRC_M0/M+_ = 1.3e-18, p_OVC_M0/M+_ = 0.006, p_GAC_M0/M+_ = 0.005, **Table S13**) or in primary vs metastatic tissues (**B**, p_OVC_ = 0.257, p_UVM_ = 0.820, **Table S14**). **C**. L1PA unique hits obtained for 15 breast cancer cell lines compared to the distribution of Cytoscan probes distributed throughout the human genome. **D.** Genome-wide z-score for all cancer (N = 564) vs healthy plasma samples (N = 120, 63 of the total 183 HDs are used as references to compute the z-score and are not displayed here, p = 1.21e-20). **E.** Genome-wide z-score by cancer subgroups vs healthy samples. The p-values are computed using Mann–Whitney *U* test (p_CRC_M+_ = 2.05e-18, p_BRC_M+_ = 1.01e-18, p_UVM_M+__= 0.169, p_LC_M+_ = 0.769, p_OVC_M+__= 1.84e-11, p_GAC_M+_ = 0.003, p_BRC_M0_ = 5.12e-17, p_OVC_M0_ = 1.09e-12, p_GAC_M0_ = 8.40e-06, **Table S15**) **F.** Correlation analysis for genome-wide z-score versus global methylation (r_overall_ = −0.62, p = 1.25e-69). **G.** Performances of the 2-step model incorporating CNA with DNA methylation analysis (Classification is done as follow: Proba_Cancer_ ≤ Threshold C1 AND GZ-score ≤ 121: prediction = Healthy; Proba_Cancer_ > Threshold C1 OR GZ-score > 121: prediction = Cancer, see methods).

A recognized marker to non-invasively estimate the tumor burden and the fraction of ctDNA is the aneuploidy or copy number alterations (CNA) ^61,62^, a hallmark of cancer genomes ^63^. Given that DIAMOND hits are dispersed throughout the genome (**Fig. 1B**), we investigated the possibility to use our data to perform CNA analysis. The mFast-SeqS approach had previously used a PCR-based L1PA targeting as a prescreening tool to estimate the fraction of ctDNA ^64,65^. This was done on native DNA whereas our data resulted from bisulfite-treated DNA.

We first tested this approach on 15 breast cancer cell lines that were also assessed by CytoScan HD microarrays for aneuploidy. DIAMOND provided an average of 78 000 uniquely mappable reads per cell line, corresponding to around 10 000 L1PA copies precisely located in the genome. These L1PA hits homogeneously overlapped with regions covered by CytoScan probes along the genome (**Fig. 5C**). We computed z-scores, quantifying copy number alterations, at the level of chromosome arms as previously described (^65,66^, see methods) and obtained similar results to those found with CytoScan arrays (**Fig. S9A**). We observed low alteration scores for the normal-like breast cell line HTERT-HME1 and good correlations between the 2 methods for the majority of the cell lines (**Fig. S9B**).

Next, we computed genome-wide z-scores in healthy and cancer plasma samples and observed high alteration scores specifically in cancer samples (**Fig. 5D**). Cancer subgroups z-scores mirrored global hypomethylation profiles (**Fig. 5E, 2B and 4D, Table S15**), both reflecting tumor burden and ctDNA fractions available. However, global methylation rates and z-scores were only moderately anti-correlated (**Fig. 5F**), demonstrating that these are partially independent markers that can provide distinct signals (**Fig. S9C**).

To obtain a final classification labelling each sample as healthy or cancer, we used a 2-step categorization incorporating CNA analysis, which improved cancer detection. We used the probability of the cancer prediction provided by the methylation-based validation models (‘expert*’* and ‘all*’* models), applying a threshold identified on the discovery cohort, followed by a reclassification of samples which presented a z-score > 121, as cancer. This cut-off value was deduced from a cross-validation applied on C1 (see methods and **Fig. S9C**). This classifier achieved high sensitivities with specificities between 97-100% for the ‘expert’ models and 93% for the integrated ‘all’ model in 4 distinct cancer types (BRC, CRC, LC, OVC, **Fig. 5G**) and could be applied as is in the clinic. This was particularly promising for localized breast cancer with a sensitivity of 100% with both model types.

## Discussion

In this study, we established a robust proof of concept that targeting hypomethylation of retrotransposons from cell-free DNA is a sensitive and specific biomarker to detect multiple forms of cancer non-invasively. Repetitive regions provide genome-wide information as they hold half of the CpG sites present in the human genome ^67^. Hypomethylation of L1 elements, which is a common feature of multiple forms of cancer, is a universal marker and helps cover the heterogeneous profiles of cancer patients in a single test. Previous methylation studies have left these regions aside as they are inherently difficult to map, and DMR analysis is commonly performed on mapped data. However, repeats were previously used to profile aneuploidy and one of the first reported liquid biopsy cancer detection test was based on serum DNA integrity calculated with the ratio of short over long Alu fragments ^68^.

Here, we developed a new pipeline to detect methylation profiles at repeats with a single base-pair resolution, without resorting to mapping on a reference genome. This allowed us to rescue unmappable sequences (such as polymorphic copies that are not annotated in the reference genomes) or sequences that are difficult to map (such as multi-mappers) and retain most of our data. This is instrumental in achieving high sensitivity. The DIAMOND assay demonstrated high performance in detecting cancer samples and we established its feasibility in six different cancer types, including three at localized stages. It outperforms mutation screening, as it covers virtually all patients and presents a promising marker for pan-cancer detection.

Previous cfDNA multi-cancer tests enabled sensitive detection and classification of multiple forms of cancers such as the CancerSeek test, based on the combination of mutations and proteins detection^11,69^ and more recently combined with aneuploidy profiling using representation of Alu sequences^70^ or the cfDNA methylation test Galleri from GRAIL, which targets 1,100,000 CpGs within unique regions ^39,40^. In comparison, DIAMOND targets about 100,000 CpG sites within 30-40,000 copies of LINE-1 using a unique set of probes for all these copies and the various cancer type tested. Very recently, several multi-cancer detection methods based on retrotransposons were published. This includes the detection of the circulating L1 protein ORF1p ^60^ as well as the probing of the representation of all type of repeats from Annapragada and colleagues ^71^. Another study demonstrated that cfDNA cleavage profiles at Alu sequences reflect their methylation status and can be used to detect liver cancer and nasopharyngeal carcinoma ^72^.

The DIAMOND assay interrogates specifically the methylation status of LINE-1 retrotransposons to obtain a global representation of the hypomethylation occurring during carcinogenesis and assess the potential of circulating DNA methylation changes at L1PA elements as a universal tumor biomarker. Our aim was to develop a new highly sensitive strategy to detect cancer-specific signatures in blood. Overall, DIAMOND reached great performances for multi-cancer detection using ‘expert’ models for specific cancer subgroups or integrated models including all types of cancers. Lower detection rate in metastatic lung cancer may be related to the fact that these samples seem to have a low tumor burden as indicated by their genome-wide z-scores (**Fig. 5E**). However, integrating other regions with cancer-specific methylation changes could help improve detecting this type of cancer. The DIAMOND assay provides methylation profiles from minute amount of cfDNA, down to a few nanograms, with high precision and high coverage using an affordable sequencing depth.

Stronger L1PA hypomethylation was associated with shorter overall survival demonstrating its prognostic value. We therefore anticipate that our method has the potential to be applied for the development of routine clinical tests. While our integrated models provide proof of concept for cancer detection with blood screening in asymptomatic patients, the ‘expert’ models could also be useful to help diagnosis when we suspect the cancer location or to perform disease follow up.

To push the DIAMOND assay towards a clinically applicable test, we also demonstrated that DIAMOND data can be used to perform copy number alterations analysis which improves cancer detection. We integrated this analysis in a classifier providing ‘healthy’ or ‘cancer’ labels for each sample and reached a detection of 91% of true positives for all cancers together and in particular a 100% sensitivity with 100% specificity for localized breast cancer with the BRC M0 expert model.

Further testing with a larger number of samples covering earlier stages, including an independent cohort of gastric non-metastatic cancers, more subtypes and different types of cancer will enable to consolidate and expand these findings. Moreover, this will strengthen the classification models, which will perform better with more samples for training and testing. In the current study, we did not control for ethnic origin of samples tested. The potential impact of L1 polymorphism linked to ancestry should be estimated in future studies. It will also be important to study the impact of other conditions, such as auto-immune diseases, which may lead to the detection of L1 hypomethylation in blood. The recent study on the detection of circulating L1 ORF1p in cancer by Taylor and colleagues^60^ demonstrated a high specificity and no sign of L1 reactivation in blood of patients with auto-immune disease, indicating that it might be a cancer-specific phenomenon. DIAMOND analysis could further be used to infer the tumor burden and monitor the disease to better detect minimal residual disease and the relapse early. However, the impact of treatments on methylation status should be investigated first.

Overall, we developed a *turnkey* analysis method that identifies tumor plasmas across multiple types of cancer with the same marker. This approach offers an optimized balance between the number of targeted regions and sequencing depth, which could extensively improve the sensitivity of ctDNA detection in a cost-effective manner and improve management of patients with cancer.

## Methods

### Materials

#### Cell lines

Cell lines screened in **Fig. 2A** are the following: CRC (HCT116); OVC (SKOV, Caov3, ES-2); BRC (MDA-MB453, SKBR3, MDA-MB361, HCC202, ZR75.1, HCC70, BT474, MDA-MB231, Cal51, MDA-MB157, BT20, MCF7, HCC1954, HCC1569, HCC38); UVM (MP38, MP41, MP46, MP65, MM28, Mel285, Mel270, 92.1, Mel202, omm2.5, Mel290, mm66, omm1).

#### Tissue and plasma samples

Archived tissue samples (ovarian adjacent tumor tissues, ovarian primary and metastatic tumors, breast tumors and uveal melanoma tissues) were retrieved from the Pathology department of Institut Curie. Healthy white blood cells and healthy plasma were collected from blood of healthy donors through the French blood establishment (agreement #16/EFS/031) under French and European ethical practices. Blood samples from patients treated at the Institut Curie (Paris, France) were collected, after written informed consent, as part of the following studies: resectable metastatic colorectal cancers from the Prodige14 trial (approved by a French Personal Protection Committee – “CPP-Comité de Protection des Personnes Sud Méditerranée IV” and registered in ClinicalTrials.gov under NCT01442935); non-small cell lung cancer and metastatic HR+ HER2-breast cancer from the ALCINA study (approved by a French Personal Protection Committee and registered in ClinicalTrials.gov under NCT02866149; treatment-naïve ovarian cancer or triple-negative breast cancer patients eligible for surgery or neoadjuvant chemotherapy from the SCANDARE study (approved by the French National Agency for the Safety of Medicines and Health Products “ANSM - Agence National de Sécurité du Médicament”, a French Personal Protection Committee and registered in ClinicalTrials.gov under NCT03017573); multiple-types of metastatic cancers from the SHIVA02 study (approved by the French National Agency for the Safety of Medicines and Health Products “ANSM - Agence National de Sécurité du Médicament”, a French Personal Protection Committee and registered in ClinicalTrials.gov under NCT03084757), non-metastatic operable gastric cancers and advanced uveal melanoma from CTC-CEC-ADN study (approved by a French Personal Protection Committee and registered in ClinicalTrials.gov under NCT02220556). Additional archived samples were also retrieved from the biobank of the Institut Curie, patients having provided informed consent for research use. All samples were obtained in accordance with the ethical guidelines, with the principles of Good Clinical Practice and the Declaration of Helsinki. This study was approved by the Internal Review Board and Clinical Research Committee of the Institut Curie. Blood samples were collected at the time of inclusion, before the start of the treatment, in EDTA tubes. Plasma was isolated within 4 h, to ensure a good quality of cfDNA, by centrifugation at 820 g for 10 min, followed by a second centrifugation of the supernatant at 16,000 g for 10 min and stored at −80°C until use.

## Methods

### Preparation of DNA from cell lines and tissues and cfDNA

Isolation of DNA from cell lines and healthy white blood cells (buffy coats) was performed using the QIAamp DNA Mini Kit or QIAamp DNA Blood Mini Kit (Qiagen) according to the manufacturer’s instructions. DNA from cryopreserved and formalin-fixed paraffin embedded (FFPE) tumor tissues was extracted using a classical phenol chloroform protocol and the NucleoSpin® FFPE DNA kit (Macherey Nagel), respectively. cfDNA was extracted from 2 ml of plasma using the automated QIAsymphony Circulating DNA kit (Qiagen), the Maxwell RSC ccfDNA LV plasma kit (Promega) or manual QIAamp circulating nucleic acid kit (Qiagen), according to the manufacturer’s instructions, and eluted in 60 μl, 75 μl or 36 μl, respectively. We verified that the extraction method did not impact our results (**Fig. S10A-B**). Isolated DNA was quantified by Qubit® 2.0 Fluorometer using dsDNA HS Assay Kit (Thermo Fisher Scientific) according to the manufacturer’s instructions and stored at −20°C until use.

### Bisulfite conversion

We used sodium bisulfite-based chemical conversion to achieve base-pair resolution analysis, which is crucial to address methylation levels at single CpG dinucleotides and the co-methylation of multiple CpG sites to determine methylation *haplotypes* (methylation state of successive CpG sites). Bisulfite treatment of the isolated genomic DNA (up to 200 ng) from the cancer tissues, cancer cell lines and buffy coats was performed using an EZ DNA Methylation-Gold Kit™ (Zymo Research, CA, USA), following the manufacturer’s instructions. Bisulfite treatment of cfDNA (isolated from 2 ml of plasma) was performed using the Zymo EZ DNA Methylation-Lightning Kit™ (Zymo Research, CA, USA), according to the manufacturer’s instructions. Bisulfite-treated DNA was stored at −80°C and further used to build a sequencing library. We have compared the methylation profiles obtained with bisulfite conversion or enzymatic conversion (NEBNext® Enzymatic Methyl-seq Conversion Module, ^73^) followed by amplification with the DIAMOND targets and deep sequencing. We observed similar methylation profiles (**Fig. S11**). We have tested various starting quantities of DNA (100ng, 10ng, 5ng) of white blood cells extracted from healthy donors (buffy coats). We have also compared cfDNA (10ng) and DNA from MCF7 breast cancer cell lines (100ng) know to display hypomethylation at L1PA elements. We observed that the methylation profiles along the 30 CpG targets are similar when starting with 100ng (**Fig. S11A**) but, in our hands, the bisulfite conversion seems more robust with decreasing stating DNA quantities (**Fig. S11B-C**). We thus observe some differences with the cfDNA samples at 10ng (**Fig. S11D**). We could still detect very similar hypomethylation profiles in MCF7 breast cancer cell lines (**Fig. S11E**). This shows that EM-seq could be used to profile L1PA methylation changes but the same conversion should be used throughout the study.

### Primer design

Eight primer pairs were designed using the LINE-1 Human Specific (L1HS) consensus sequence from Repbase (**Fig. 1A**). Although 5’UTR (promoter region) is CpG-rich and common target for methylation quantitation, L1PA copies are frequently 5’-truncated. Therefore, primers were also designed for ORFI and ORFII to target more L1PA elements and improve the sensitivity of our assay. All primers were designed for plus strand of bisulfite converted DNA, using the MethPrimer or PyroMark Softwares. Targeted regions contained 2-7 CpG targets and ranged from 101bp to 150bp, to better capture cfDNA fragments, which have a mean size of 167bp ^74^, (**Table S1**). Primers were methylation-independent, encompassing 0 to 2 CpGs (none toward the 5′ end), and were degenerated to target both the methylated and unmethylated states. They contained Fluidigm universal CS (common sequence) tags at their 5′ ends. We incorporated a 16 N (random nucleotides) as unique molecular identifiers (UMI) between the target-specific sequence and the CS2 in the reverse primers for signal deconvolution to detect true low frequency alterations and for reducing errors. As LINE-1 hold thousands of copies per genome, a high number of distinct UMIs is essential for unique barcoding of each target molecule. The 16 N stretch between the target-specific sequence and the CS1 in forward primers was used to increase diversity of sequencing libraries and improve sequencing quality. All primers were obtained from Eurogentec (RP-cartridge purification method).

### Preparation of targeted bisulfite sequencing libraries

To limit batch effects, sequencing and library preparation batches contained both cancer samples and healthy donors. We also specifically processed the validation cohort C2 with equilibrated number of healthy and cancer samples distributed in 5 experimental batches. Sequencing libraries were prepared using three PCR steps (**Fig. S1A**): 1) target-specific linear amplification for UMI assignment, 2) target-specific exponential amplification and 3) barcoding PCR for sample identification. Each library was prepared in two individual reactions (due to the overlap of amplicon 2 with other primers), including: *I)* Multiplex PCR amplification of 7 probes (Amplicon 1, 3, 4, 5, 6, 7, 8), and *II)* Single PCR amplification of amplicon 2.

UMI assignment for multiplex reaction was performed using Platinum™ Multiplex PCR kit Master Mix (Thermofisher, Life Technologies SAS) in a 25 μL reaction containing 1x Platinum™ Multiplex PCR Master Mix, 0.01-0.06 µM mix of reverse primers and up to 5 ng bisulfite-converted DNA at the following thermocycling conditions: 95°C for 5 min followed by 1 cycle at 95°C for 30 s, 58°C for 90 s, 72°C for 40 s. UMI assignment for single reaction was performed using Hot Star Taq Plus DNA Polymerase (Qiagen) in a 25 μL reaction containing 1x Taq PCR Buffer, 0.65 U Hot Star Taq (5U/μL), 0.2 μM dNTPs, 1.5 mM MgCl2, 0.1 μM amplicon 2 reverse primer, up to 4 ng of bisulfite-converted DNA at the following thermocycling conditions: 95°C for 10 min followed by 1 cycle at 94°C for 60 s, 58°C for 30 s, 72°C for 40 s. To ensure complete removal of the reverse primers and dNTPs, each 25 μL reaction was treated with 50U of Exonuclease I and 10U of FastAP Thermosensitive Alkaline Phosphatase (Thermo Fisher Scientific) at 37°C for 1 h and heat-inactivated at 80°C for 15 min.

Target-specific exponential amplification for multiple reaction was performed using Platinum™ Multiplex PCR kit Master Mix in a 50 μL reaction containing 1x Platinum™ Multiplex PCR Master Mix, 0.01-0.06 µM mix of forward primers, 0.2 μM CS2 reverse primer and 20 μL of purified PCR product at the following thermocycling conditions: 95°C for 5 min followed by 28 cycles at 95°C for 30 s, 58°C for 90 s, 72°C for 30 s followed by a 10 min incubation at 72°C. Target-specific exponential amplification for single reaction was performed using Hot Star Taq Plus DNA Polymerase in a 25 μL reaction containing 1x Taq PCR Buffer, 0.65 U Hot Star Taq (5U/μL), 0.2 μM dNTPs, 1.5 mM MgCl2, 0.2 μM amplicon 2 forward primer, 0.2 μM CS2 reverse primer and 8 ul of purified PCR product at the following thermocycling conditions: 95°C for 10 min, 25 cycles at 94°C for 60 s, 58°C for 30 s, 72°C for 30 s and 10 min at 72°C.

PCR products of multiplex and single reaction were pooled together after quantification by qPCR and purified using Agencourt AMPure XP (Beckman Coulter) at 1.2x ratio according to the manufacturer’s protocol. Purified DNA was eluted in 30 ul of water. Barcoding PCR was performed using universal fluidigm primers. 25 μL of purified pooled PCR product, 1x Phusion HF Buffer, 1 U Phusion Hot Start II DNA Polymerase (Thermo Fisher Scientific), 0.2 μM fluidigm primer, and 0.2 mM dNTPs were mixed in the final volume of 50 μL and amplified with the following conditions: 98 °C for 2 min, followed by 20-25 cycles of 98 °C for 10 s, 62°C for 30 s, and 72 °C for 30 s followed by a 5 min incubation at 72°C. The amplified product was purified by two consecutive AMPure XP steps using *1)* a low concentration of AMPure XP beads (0.6x – 0.7x ratio) where the beads containing the larger fragments are discarded and supernatant collected (reverse purification) and *2)* higher beads concentration (1.1x – 1.2x ratio) where the beads containing fragments of interest were collected and purified according to the manufacturer’s protocol. Size-selected libraries were eluted in 15 μL of low-EDTA TE buffer. The libraries were quantified with Qubit HS DNA kit (Thermo Fisher Scientific), qualified with nano-electrophoresis (TapeStation, Agilent), and pooled equimolarly for sequencing. Sequencing was performed on Illumina HiSeq rapid run mode or NovaSeq (PE 30bp, 170bp).

### Preprocessing of the reads

For each sample, FASTQ files containing raw sequences, composed by the following parts: CS1, forward UMI, forward primer, insert, reverse primer, reverse UMI, and CS2 (**Fig. S1A**) were first filtered for reads quality (average >Q20 per read) and then demultiplexed (i.e., cut using *atropos* v1.1.31) using forward and reverse primer sequences. FASTA files were created per primer-set, containing inserts and reverse UMIs for deduplication, as they are unique for each input DNA molecule. Inserts and reverse UMI were then filtered on expected sizes (with a tolerance of ± 5 bases for the inserts). Filtered inserts and UMIs sequences were concatenated and deduplicated using *vsearch* v2.15.2. Reverse UMIs were then trimmed and resulting inserts from all samples were aggregated into a single FASTA file per primer-set.

### Clustering, extraction of representative sequences and global alignment

Using *vsearch* (with the following parameters: --cluster_fast <inputFasta> --notrunclabels -- fasta_width 0 --iddef 4 --id 0 --qmask none --clusterout_sort --consout <referenceFasta>), a clustering based on sequence identity was applied to each FASTA file, or a subset of 20 million reads randomly chosen if a given file comprised more. The 10 largest clusters’ representative sequences were isolated in separate files. Using *mafft* v7.508 (with the following parameters: -- globalpair --maxiterate 1000), the 10 representative sequences were aligned pairwise resulting in a reference database for each primer-set. Lastly, using *mothur v*1.48.0 (with the following parameters: #align.seqs(candidate=<inputFasta>, template=<referenceFasta>, align=needleman, match=1, mismatch=-1, gapopen=-1, gapextend=0)) on each primer-set FASTA file, all sequences from all samples were aligned to the corresponding reference.

### CG calling, methylation levels and haplotypes extraction

To call CpG dinucleotides of interest, a sliding window of 2 bp was used on all aligned sequences to determine the distribution of dinucleotides along each amplicon target. A first threshold of ≥ 20% of CG/TG dinucleotides was used to select potential CpG sites. A second threshold was applied to eliminate dinucleotide with ≥95% TG and select position with at least 5% methylation rate. From the aligned sequences, the patterns of methylation were extracted and compiled into either average levels of methylation at each previously identified CpG sites, or proportions of methylation haplotypes for each sample.

### Machine learning-based classification models

The resulting data (represented as average levels of methylation per CpG site or proportions of methylation haplotypes or both) were used to do supervised learning of statistical models using the random forest classifier algorithm ^75^ from Python package scikit-learn ^76^, with the following hyperparameters: n_estimators=300, criterion=’gini’, max_depth=None, min_samples_split=2, min_samples_leaf=1, min_weight_fraction_leaf=0.0, max_features=’sqrt’, max_leaf_nodes=None, min_impurity_decrease=0.0, bootstrap=True, oob_score=False, warm_start=False, class_weight=None, ccp_alpha=0.0, max_samples=None.

The rational for choosing random forest over other learning methods was driven by three main factors: *1)* it is less prone to overfitting ^75^; *2)* it shows excellent performance even when the quantitative relationship between features and observations is biased in favor of the former, such as when using methylation haplotypes data representation ^77^; *3)* random forests also inherently return measures of variable importance ^75^, such as mean decrease in impurity, which greatly facilitate the interpretability of model decisions. The features used to train the models were the average levels of methylation per CG site (n=30), the proportions of methylation haplotypes (i.e., the combinatorial of all the possible methylation status of CG sites within a given amplicon, n=372) or both. No additional transformation nor feature selection was performed on the data.

### Expert and All cancer models

Model classifications were run 5000 times in order to estimate variance and confidence intervals. For the discovery step, in each run, as many samples from each class were randomly drawn to construct a balanced subset of the data ^78^. The samples from these draws were stratified by class and split into 60% for training, 40% for evaluation. For the validation step, we trained the model on the entire cohort 1 and evaluated it on cohort 2. The true and false positive rates for all possible classification threshold were evaluated at each run, with interpolation to generate an average ROC curve with 95% confidence interval for the 5000 runs. 95% confidence interval has been calculated with the following formula: Μ±z*s/√n with M the average of the variable, z the confidence level (z∼1.96 for 95%CI), s the standard deviation, n the number of samples in the variable.

### Blind models

We trained a random forest on haplotypes features, removing one cancer type or subgroup from the training set. The specific cancer type or subgroup is then assessed in the test set. We pooled together the discovery and validation cohorts, training on 2/3 of all the samples-- excluding the cancer type or subgroup to test for-- and testing on the remaining one-third of the samples. The only exception was metastatic gastric cancer: as they are made up of only 3 samples, they were systematically moved to the test set, consequently blinding the model towards GAC M+ samples. We also trained a stacked version with this setup (see below).

### Stacked machine learning model

We developed a model referred to in the paper as “Stack”. This model uses one Random Forest model for each combination of cancer type and metastatic status, known as the “expert sub-model.” Each expert sub-model was trained on one-third of the healthy plasma samples and one-third of the samples matching the cancer subgroup of interest (cancer type and dissemination status). These expert sub-models were then combined into a Random Forest Stack model, which uses both the haplotype features and the probabilities output by each expert sub-model. The final Random Forest Stack model was trained on an additional one-third of the healthy and cancer plasma samples and tested on the remaining samples (which represent one third of the healthy samples and one third of each subgroup).

### Mutation screening for ovarian cancer samples

Ovarian tumor genotyping was performed using the TIGER panel previously developed by Institut Curie^79^, which targets 78 genes or using ‘custom NGS’ with amplicons targeting *Tp53* and *TSC2*, which are the 2 most frequently mutated genes in this group of patients, with TruSeq library constructions for low input material (dual strand technology). After mutations identification, ovarian cancer plasmas were genotyped using custom NGS or ddPCR as previously done^14^. Sequencing was performed on a MiSeq V3-150 (25M) with Paired End 75bd protocol. Results are presented in **Table S8**.

### Whole genome bisulfite sequencing analysis

To see if we could retrieve cancer-associated L1PA hypomethylation in others plasma studies, we analyzed data sets from 2 recent studies profiling cell-free DNA methylation with Whole Genome Bisulfite Sequencing (WGBS) in healthy and cancer patients (Liu *et al.* 2024^53^; Gao *et al.* 2022^54^). Liu *et al.* analyzed methylation at 75 617 CpGs in the whole genome (estimated with Bis-SNP after mapping with Bismark) of 17 healthy donors and 31 cancer patients. We extracted the methylation levels at CpG residing within L1PA families hit by DIAMOND (mostly L1HS-L1PA10, see **Fig. 1C**). To do so, we identified CpG dinucleotides covered by L1HS-L1PA10 elements, and their position within L1 based on the L1HS consensus sequence, and we selected the DIAMOND copies using Bedtools^80^. Gao *et al.*, provided FASTQ files data for 123 breast cancer and 40 healthy patients. In order to avoid breast-cancer subtype effect and age effect, we subsampled the data to generate an age matched cohort with 16 healthy and 15 HR+HER2-M+ patients. We first merged the paired-end reads using Fastp^81^ and mapped them on the hg38 reference genome using *bismark*. At this step, we followed two different approaches to obtain the methylation level at the 30 L1PA CpGs that we studied with DIAMOND. 1) Using *bismark extractor*, we retrieved the methylation levels of all CpGs covered and intersected it with the dinucleotides covered by DIAMOND using Bedtools. Results produced with this approach are subsequently called “Gao et al (2022) Bismark”. 2) Using *mothur*^82^, we aligned the reads that mapped on L1HS-L1PA10 elements on an aggregate of native and converted (CG converted to TG) reference sequences of L1HS. All read that mapped with a score lower than 51% of similarity (score computed by *mothur*) were rejected (this threshold was established from the percentile at 99% of similarity score distribution from 1000 random sequences 167 bp long).

Results generated by this approach are subsequently called “Gao et al (2022) *Mothur*”. Finally, for “Gao et al (2022) Bismark” and “Gao et al (2022) Mothur “, we refined the results to only take in account the copy of L1HS-L1PA10 targeted by DIAMOND. Further refinements were done to only consider L1HS-L1PA3 copies with the second approach and is referred as “L1HS-L1PA3”.

### Survival analysis

Survival analysis has been performed with *survival* and *survminer* R packages - https://cran.r-project.org/web/packages/survival/citation.html, doi:10.32614/CRAN.package.survminer.

### Copy number alterations analysis

Cytoscan HD microarrays: 250 ng of gDNA from 15 breast cell lines (1 normal-like: HTERT-HME1 and 14 cancer cell lines: MDA-MB231, MDA-MB453, HCC1569, BT20, HCC1954, HCC38, MDA-MB361, ZR 75.1, MDA-MB157, MCF7, SKBR3, HCC202, HCC70, BT474) were characterized using Affimetrix/Thermo Cytoscan HD microarrays at the Genomics facility of Institut Curie to profile aneuploidy. To compare with the z-score by chromosome arm, we calculated the mean of Weighted Log2 combining probes by chromosome arms.

DIAMOND CNA: 1) Z-score calculation: prepossessed reads were uniquely mapped on hg38 genome using Bismark (version 0.23.1). As in Belic *et al.* 2015 ^66^, only the reads with an alignment score > 15 were kept. Resulting reads from all amplicons (excluding #2 and #3) were merged and normalized number of reads per chromosome arm (excluding sexual chromosomes X and Y) per sample were calculated with R. Next, the amplifications/deletions score was computed using the following formula:

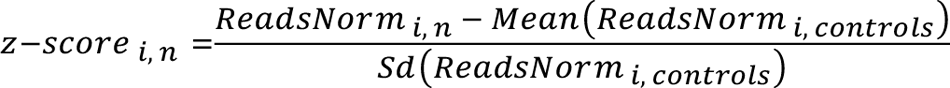

with *i* = a given chromosome arm, *n* = a given sample and *controls* = a set of reference samples (white blood cells from 10 healthy reference samples for the cell lines, 63 healthy plasma from C1 as a reference for cancer and healthy plasma samples). Genome-wide z-scores were computed by summing the squared z-scores of all chromosome arms. 2) Z-score threshold identification: to identify altered versus normal z-scores, we performed 5-fold cross validation of simple cutoff classification model on the discovery cohort (N_Healthy_ = 60, N_Cancer_ = 350) using the genome-wide z-score and calculated the threshold that maximize the sensitivity at 100% specificity.

### 2-step classification for sample labelling

First, we selected the threshold for the probability of the cancer prediction (Proba_Cancer_) on the discovery cohort maximizing the sensitivity for a 99% specificity, per ‘cancer-type’ model. We applied this threshold on the Proba_Cancer_ computed with the validation models and reclassified samples which presented a z-score > 121, as cancer (Proba_Cancer_ ≤ Threshold C1 AND GZ-score ≤ 121: prediction = Healthy; Proba_Cancer_ > Threshold C1 OR GZ-score > 121: prediction = Cancer) see **Tables S16-22.**

## Data Availability

Data have been deposited as methylation matrices (CG% or haplotypes%) on the Zenodo database with the following accession code: https://zenodo.org/uploads/12206227 and as compressed FASTQ files at the European Genome-Phenome Archive at https://ega-archive.org/ under the accession code <u>EGAD50000000646</u>. WGBS sequencing data were downloaded from publicly available database at https://zenodo.org/records/7779198 and from the National Center for Biotechnology Information (https://www.ncbi.nlm.nih.gov) under the accession number PRJNA494975. The code used to analyze the data is available on github: https://github.com/ProudhonLab.

## Acknowledgments

We thank the members of the C.P.’s laboratory for critical reading of the manuscript. We are grateful to D. Bourc’his and her team for hosting us during part of this study. We thank the members of the ICGex NGS platform of the Institut Curie, especially S. Lameiras, V. Raynal and S. Baulande for advice and the non-profit organization “La Vannetaise” for financial support.

## Funding

The NGS facility was supported by ANR-10-EQPX-03 (Equipex) and ANR-10-INBS-09-08 (France Génomique Consortium) grants and by the Cancéropôle Île-de-France.

This research was supported by grants, of which C.P. was recipient, from: The European Research Council (ERC-StG EpiDetect),

The Ligue contre le cancer (RS17-75-75),

The prematuration program of the Centre National pour la Recherche Scientifique (CNRS), The SiRIC 2 Curie program (INCa-DGOS-Inserm_12554),

The DEEP Strive funding (LABEX DEEP 11-LBX0044).

CAA research was supported in part by the French government under management of Agence Nationale de la Recherche as part of the “Investissements d’avenir” program, reference ANR-19-P3IA-0001 (PRAIRIE 3IA Institute).

## Author Contributions

MM, MH and CP designed the study

MM, MH, AM, CH, MG, VD, CB, MS, CR and DG performed the experiments MM, MH, AM, KDS, MG, KVG, CAA and CP analyzed the data.

MM, AM, KDS, MG and KVG performed the statistical analyses.

CH, AR, FCB, JYP, CL, MK, CLT, IB, MHS, OL, LC contributed with identification of clinical samples.

MM, MH, AM, KDS, MG, CAA and CP wrote the manuscript.

All authors participated in revising the manuscript and approved this final version.

## Supplementary materials

**Fig S1.**
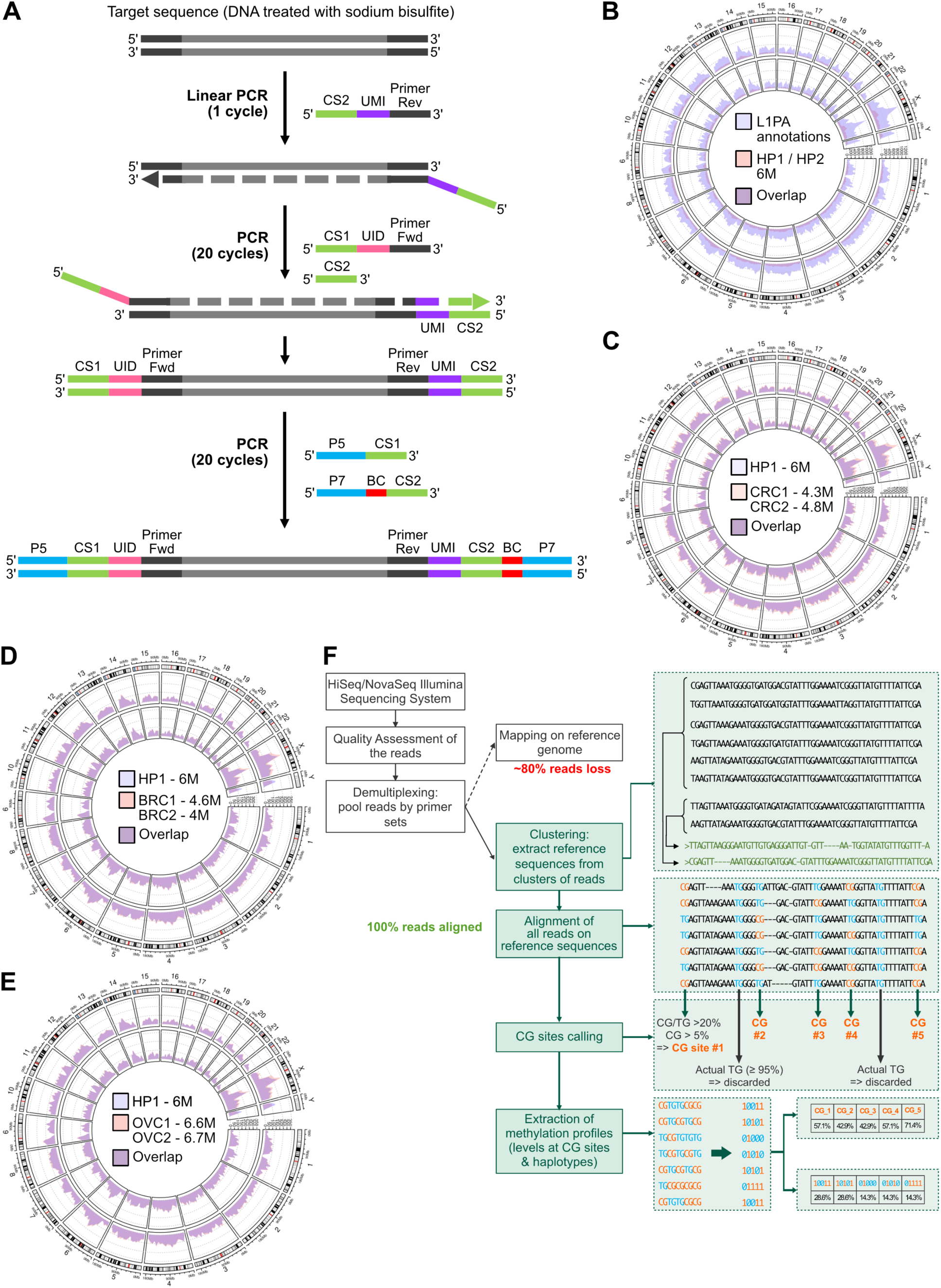
**A.** Scheme of the targeted bisulfite sequencing strategy used to build the DIAMOND assay libraries. The protocol starts by the incorporation of unique molecular identifiers (UMI) via 1 cycle of linear PCR to identify each initial molecule present in the sample. We also incorporated a 2^nd^ set of molecular identifiers (UID) during the 2^nd^ PCR in order to generate libraries with enough nucleotide diversity which is crucial for a successful downstream sequencing (See method section for more details). **B-E.** Hits obtained across the genome using the DIAMOND assay with standard coverage (indicated in million reads – M) in healthy plasma (HP) over the L1PA annotations **(B)**, in HP versus colorectal cancer (CRC) plasmas **(C)**, in HP versus breast cancer (BRC) plasmas **(D)**, and in HP versus ovarian cancer (OVC) plasmas **(E)**, (see also **Table S2**). **F.** Summary flow chart illustrating the pipeline developed for reference-free alignment of sequencing data (see also Methods).

**Fig S2.**
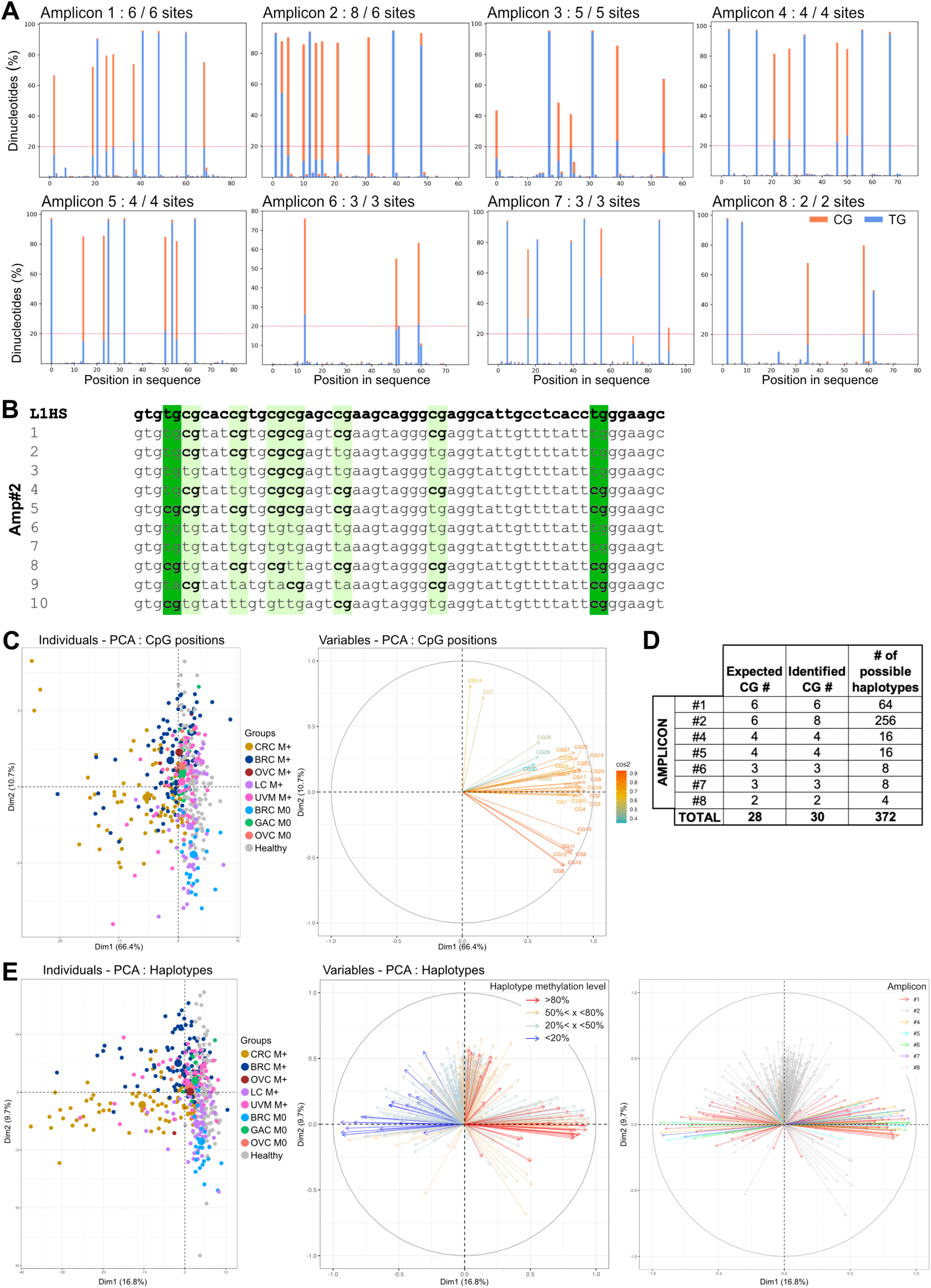
**A.** Proportion of CG and TG dinucleotides within the 8 regions targeted, after reference-free alignment. Number of CpG sites detected vs expected are indicated for each amplicon. The dashed red line represents the first threshold applied to select dinucleotide positions with >20% (GC+TG). A second threshold of >5% CG is applied. **B.** Alignment of 10 representative sequences of the largest clusters obtained for amplicon 2 relative to the L1HS consensus sequence, highlighting 2 additional CpG positions identified (dark green). **C.** Principal component analysis, based on the average methylation level at each CpG position (n=30), showing the distribution of healthy and cancer samples annotated for their cancer subgroups in the two first dimensions (left panel), and the contribution of CpG positions used as components (right panel). **D.** Number of possible haplotype features extracted from the 30 CpGs within the 7-amplicon panel. **E.** Principal component analysis, based on haplotypes proportions (n=372), showing the distribution of healthy controls and cancer samples annotated for their cancer subgroups in the two first dimensions (left panel), and the contribution of haplotypes used as components (middle and right panels). Middle panel highlights 4 groups of methylation levels relative to the haplotype components. The right panel highlights the contribution of the various amplicons.

**Fig S3.**
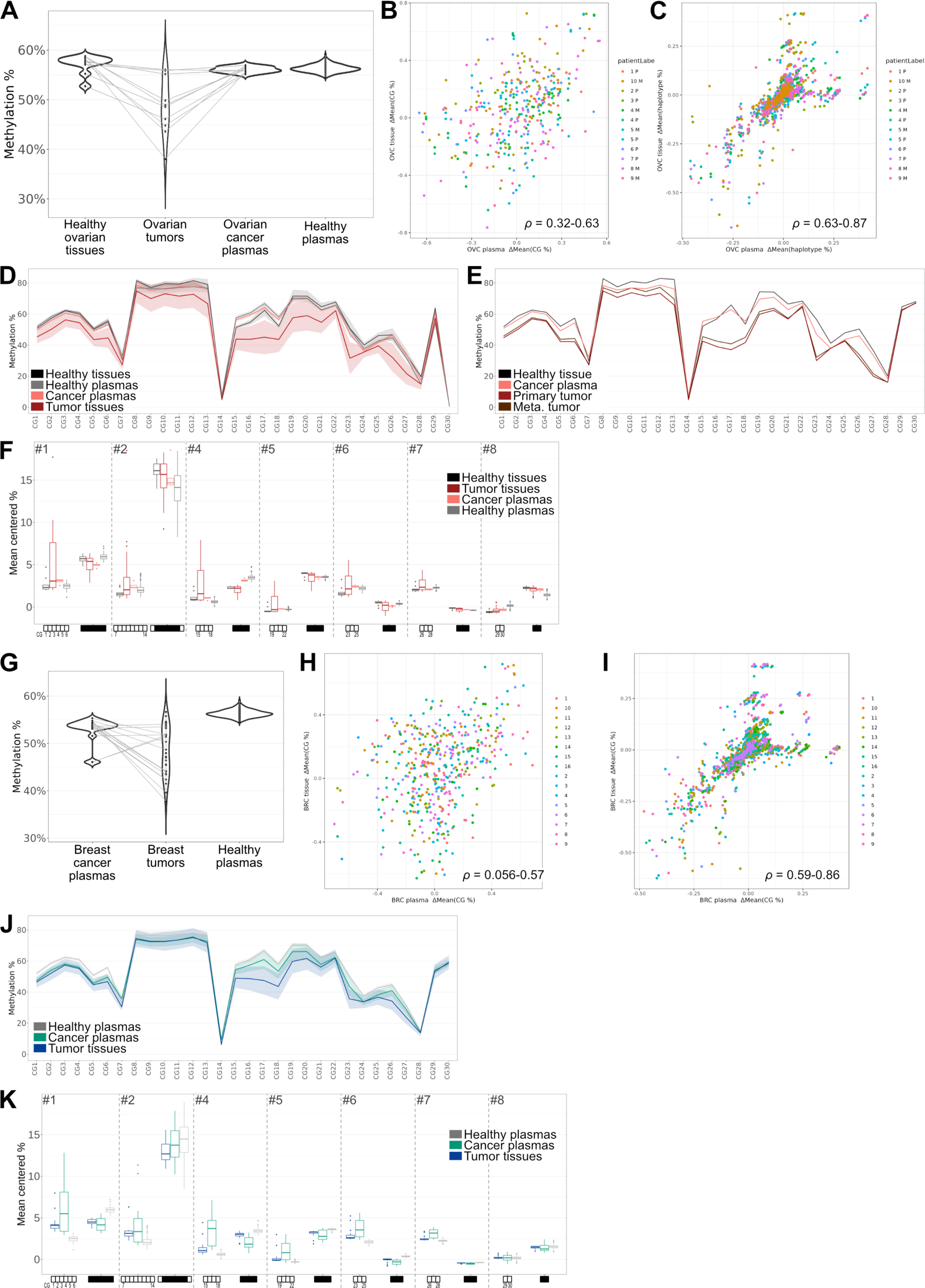
**A-F.** Analysis of ovarian tumor and plasma paired samples. **A.** Global DNA methylation in ovarian paired samples (ovarian healthy tissues, ovarian tumors: 7 primary and 3 metastatic tumors, ovarian cancer plasmas) from 10 patients versus healthy donor plasmas, N=183. **B-C.** Correlation of the methylation differences, relative to the mean methylation of healthy donor plasmas, between tumor and plasma paired samples. Ranges of correlation scores (*ρ*) are indicated. **B.** Correlation for single CpG methylation levels (30 features per sample). **C.** Correlation for methylation haplotype proportions (372 features per sample). **D.** Mean methylation observed at each CpG position across L1PA elements in healthy ovarian tissues, ovarian tumors (primary or metastatic), ovarian cancer plasmas and healthy donor plasmas. **E.** Methylation pattern observed at each CpG position in paired samples from patient with healthy ovarian tissue, primary tumor, metastatic tumor and cancer plasma. **F.** Proportion of fully methylated or fully unmethylated haplotypes, for each amplicon (mean centered per amplicon) in paired ovarian healthy tissues, tumor tissues, cancer plasmas and healthy donor plasmas. **G-K.** Analysis of breast tumor and plasma paired samples. **G.** Global DNA methylation in breast primary tumors and cancer plasmas from 16 patients versus healthy donor plasmas, N=183. **H-I.** Correlation of the methylation differences, relative to the mean methylation of healthy donor plasmas, between tumor and plasma paired samples. Ranges of correlation scores (*ρ*) are indicated. **H.** Correlation for single CpG methylation levels (30 features per sample). **I.** Correlation for methylation haplotype proportions (372 features per sample). **J.** Mean methylation observed at each CpG position across L1PA elements in breast tumors, breast cancer plasmas and healthy donor plasmas. **K.** Proportion of fully methylated or fully unmethylated haplotypes, for each amplicon (mean centered per amplicon) in paired breast tumor tissues, cancer plasmas and healthy donor plasmas. Paired samples and their correlation details are listed in **Table S6.**

**Fig S4.**
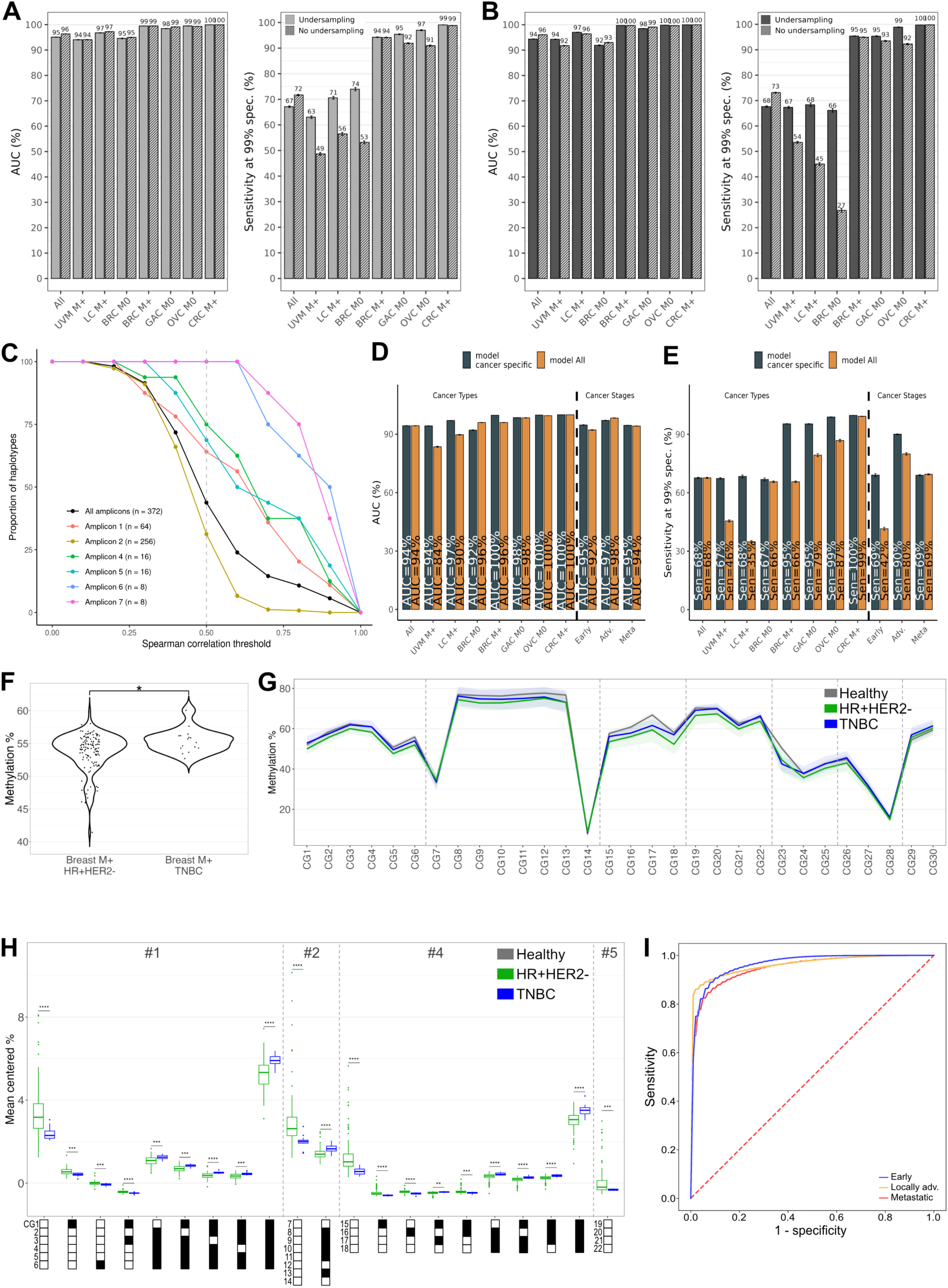
**A-B.** Performances for classifiers using single CpG methylation features (**A**) or haplotype features (**B**) with undersampling (plain bars) or not (hatched bars) presented as AUCs or sensitivities at 99% specificity. **C.** High correlation between haplotype representation and CpG positions shown by the proportion of haplotypes with at least one correlation to CpG sites along spearman rho correlation thresholds. Amplicons 1, 4-8 show high correlation all along. Amplicon 2 shows lower correlations due to its very high number of haplotypes (n = 264). n: number of haplotypes per amplicon. **D-E.** Performances for each subgroup tested separated (black bars) or from the model testing all cancer samples together (orange) using haplotype features presented as AUCs (**D**) or sensitivities at 99% specificity (**E**). **F-H.** Comparison of methylation patterns in hormone dependent (HR+HER2-, N=125) vs triple negative (TNBC N=16) metastatic breast cancers at the overall level (**F**), single CpG level (**G**) or the haplotype level (**H**). BRC M+ from C1 and C2 are analyzed (see **Table S7**). **I**. ROC curves obtained for plasma samples classification with the 3-stage model, using single-CpG methylation rates.

**Fig S5.**
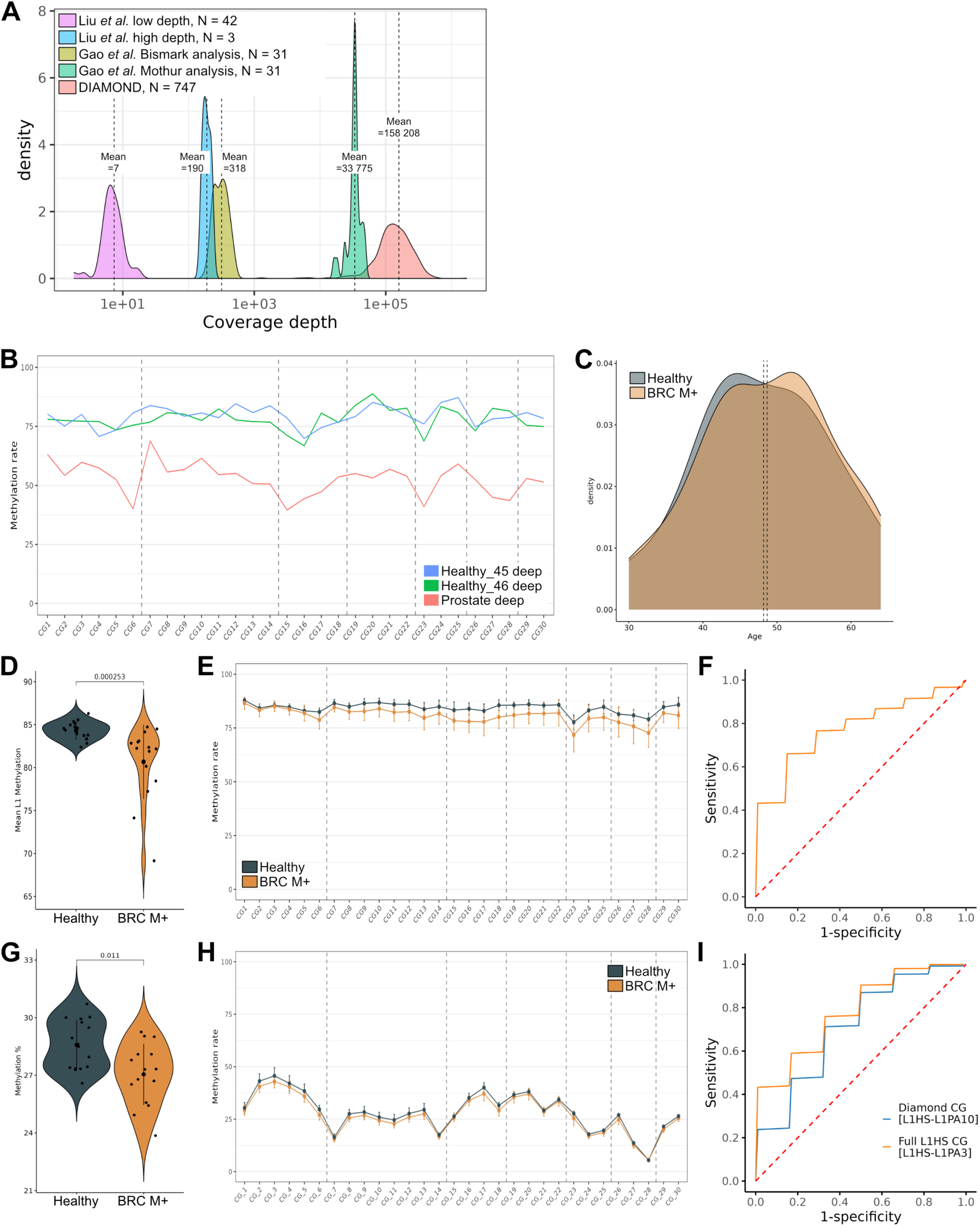
**A-I.** Analyses performed with whole genome bisulfite sequencing (WGBS) data (see methods section for more details). **A.** Mean coverage observed at each 30 CpG position belonging to the L1HS-L1PA10 copies in the various analyses performed from WGBS data and DIAMOND. **B.** Methylation pattern observed across the 30 CpGs targeted with DIAMOND in 2 healthy plasma samples and 1 prostate plasma sample extracted from the WGBS data published in Liu *et al.* ^53^. Methylation data were available as CG% at 75 617 CpGs genome wide. Only these 3 samples presented high enough coverage to profile reliable methylation patterns along L1PA elements (covering 26 135 CpGs). **C-I.** Analysis of WGBS data from Gao *et al.* ^54^, which profiled metastatic hormone dependent breast cancer (BRC M+) and healthy plasma samples. **C.** Age distribution of the age-matched cohort selected and analyzed, composed of 16 healthy and 15 BRC M+ plasmas. **D-F.** Methylation data analyzed with *bismark extractor* (‘Bismark analysis’). This includes positions specifically annotated as CpGs. **D.** Average DNA methylation levels demonstrating significant hypomethylation in BRC M+ (p=2.53e-04). **E.** Methylation patterns observed with WGBS analyzed with *bismark extractor* demonstrating lower methylation in BRC M+ across the 30 CpG of L1PA elements studied with DIAMOND. **F.** ROC curve obtained for healthy versus cancer plasmas classification with single-CpG methylation rates (AUC = 78%). **G-I.** Methylation data analyzed by extracting the CpG positions covered by L1HS-L1PA10 DIAMOND hits independently of the nature of the dinucleotide observed in the WGBS (‘Mothur analysis’). **G.** Average DNA methylation levels demonstrating significant hypomethylation in BRC M+ (p=0.011). **H.** Methylation patterns observed with WGBS analyzed with *mothur* demonstrating lower methylation in BRC M+ across the 30 CpG of L1PA elements studied with DIAMOND. **I.** ROC curves obtained for healthy versus cancer plasmas classification with single-CpG methylation rates, using positions covered by L1PA1-10 elements hit with DIAMOND (AUC = 71%) or by only L1HS, L1PA2 and L1PA3 elements hit with DIAMOND (AUC = 78%), which represent more than half of the data obtained with our targeted method.

**Fig S6.**
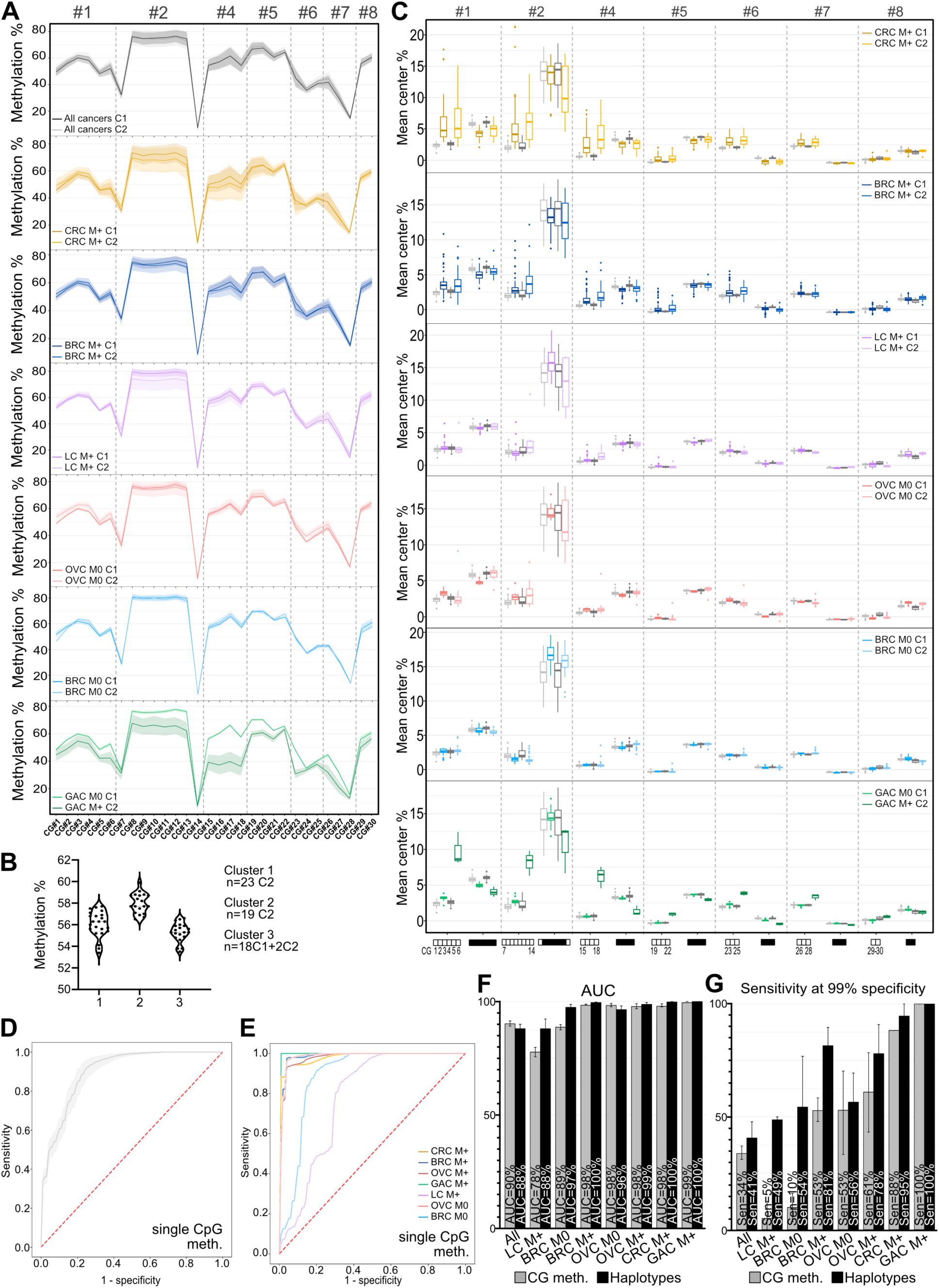
**A.** Comparison of the methylation level at individual CpG sites along the L1 for each cancer subgroup in cohort 1 (C1) versus cohort 2 (C2). Amplicon numbers are indicated. **B.** Clustering analysis on OVC M0 stages of cohort 1 (C1) and cohort 2 (C2) showing a split of C2 into cluster 1 and 2 and separation of the whole cohort 1 in a single cluster (cluster 3) including 2 samples of C2. **C.** Comparison of the haplotype proportions for each cancer subgroup and the healthy controls in cohort 1 (C1) versus cohort 2 (C2). Light grey = healthy donors C1, dark grey = healthy donors C2. Statistical analyses are reported in **Table S11**. **D-E.** ROC curves obtained for plasma samples classification in the validation cohort with the ‘all cancers’ model (**D**) or the ‘cancer-types’ models (**E**) using single CpG methylation features. All classifications include 5000 stratified random repetitions of learning on the whole discovery cohort and testing on the whole validation cohort without undersampling. **F-G**. Performances for validation classifiers using CpG methylation (grey) or haplotypes (black) features presented as AUCs (**F**) or sensitivity at 99% specificity (**G)**. Average AUCs are computed from the 5000 AUCs generated by each repetition of learning. Bars indicate 95% CI.

**Fig S7.**
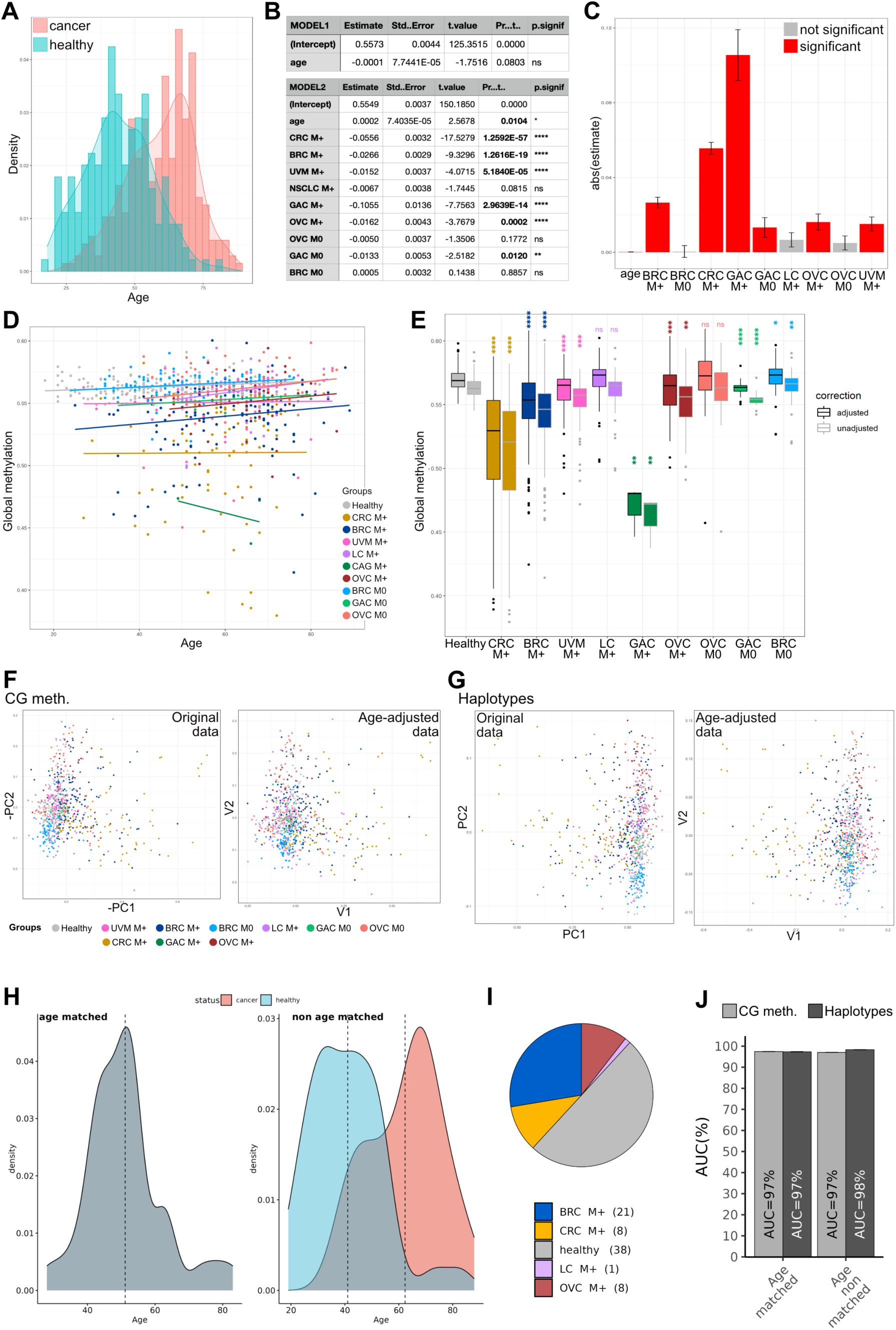
**A.** Age distribution of all cancer vs healthy samples (C1 + C2). **B.** Linear models demonstrating that the age did not predict the methylation patterns alone (Model 1) and had a significant but small effect (see also (**C**)) in combination with the ‘biological class’ representing the cancer subgroups (Model 2). **C.** Impact extent of the parameters tested represented by the absolute values of their Estimate (red: significant, grey: not significant). **D.** Global methylation levels for all samples versus their age. Correlation curves illustrate small impact of age on the methylation patterns in our study, with an increase in methylation with age. **E.** Global methylation levels of original data next to age-adjusted data, organized by subgroups, demonstrating that cancer subgroups versus healthy differences remained similar when adjusting for the age. Statistical differences between each cancer subgroup and healthy samples were computed using Mann– Whitney *U* test (see **Table S12** for detailed p-values and CI). **F-G.** PCA analysis on original data (left panel) or age-adjusted using AC-PCA to adjust for confounding factors conjointly (right panel) for single CpG methylation levels (**F**) or haplotypes (**G**). Adjusting for the age seemed to have limited impact on the data indicating that age is not a confounding factor here. **H.** Age distribution in the age-matched vs non-age-matched cohorts extracted from C2. **I.** Both cohorts are composed of 76 samples including half healthy and the same proportions of M+ cancer types. **J.** Classification performances for ‘all cancer’ model with age-matched vs non-age-matched cohorts presented as AUCs.

**Fig S8.**
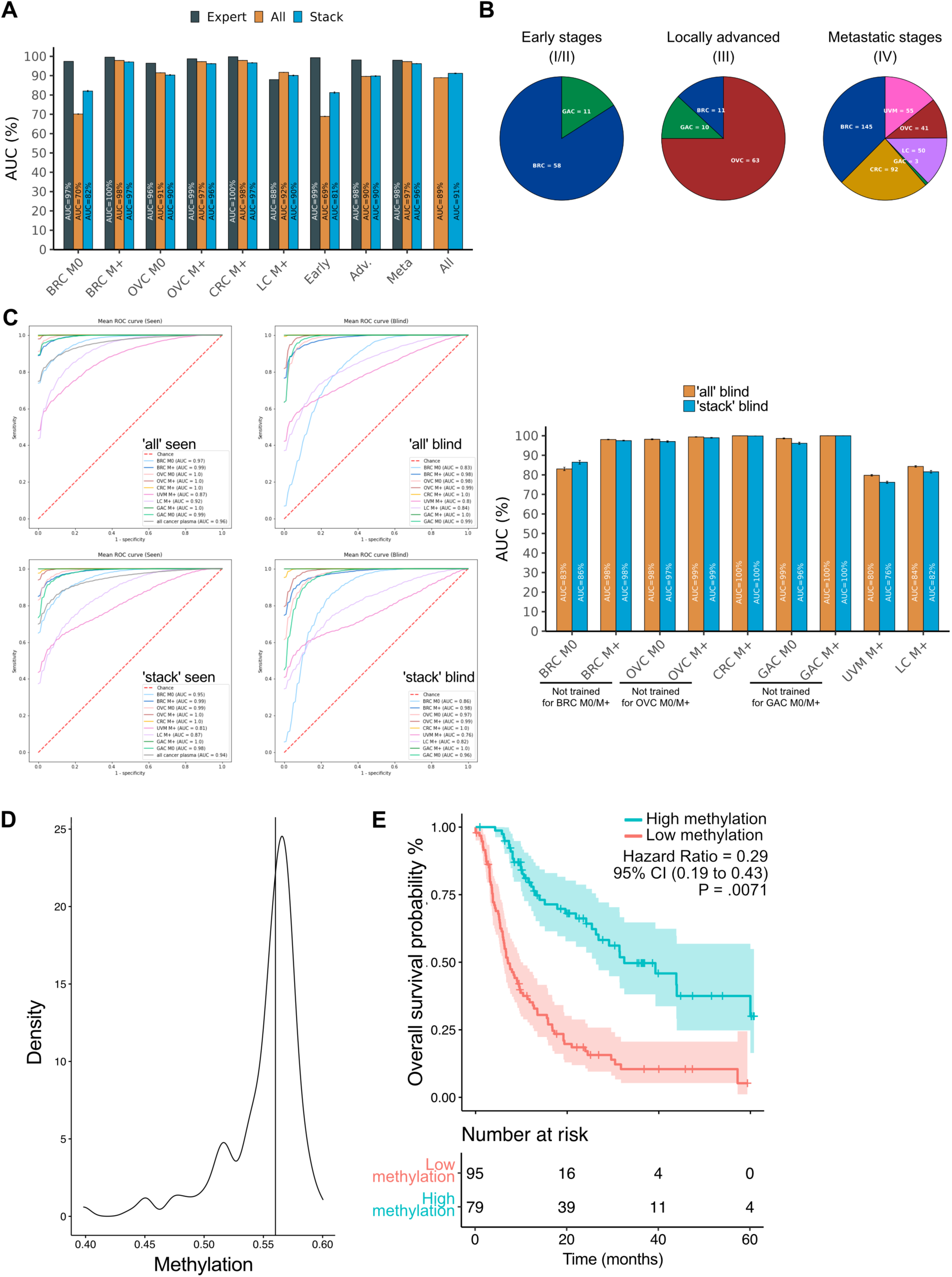
**A.** Comparison of the performances of the ‘expert’ models testing each subgroup separated (black bars), the ‘all’ model testing all cancer samples together (orange) or the ‘stack’ model integrating both models (blue bars) using haplotype features presented as AUCs. **B.** Composition of cancer types per stage-groups (early (I/II), locally advanced (III) or metastatic (IV)). **C.** Performances for integrated models (‘all’ or ‘stack’) when training for the specific cancer type tested (seen) versus when not training for this cancer type overall (blind). These classifications have been performed on the whole sample set including C1 and C2. BRC M0/M+, OVC M0/M+ or GAC M0/M+ have been removed all together. **D.** Distribution of the global methylation levels in patients of the validation cohort. Median = 56%. **E.** Survival analysis comparing patients with high and low methylation levels (N=79 vs N=95 respectively).

**Fig S9.**
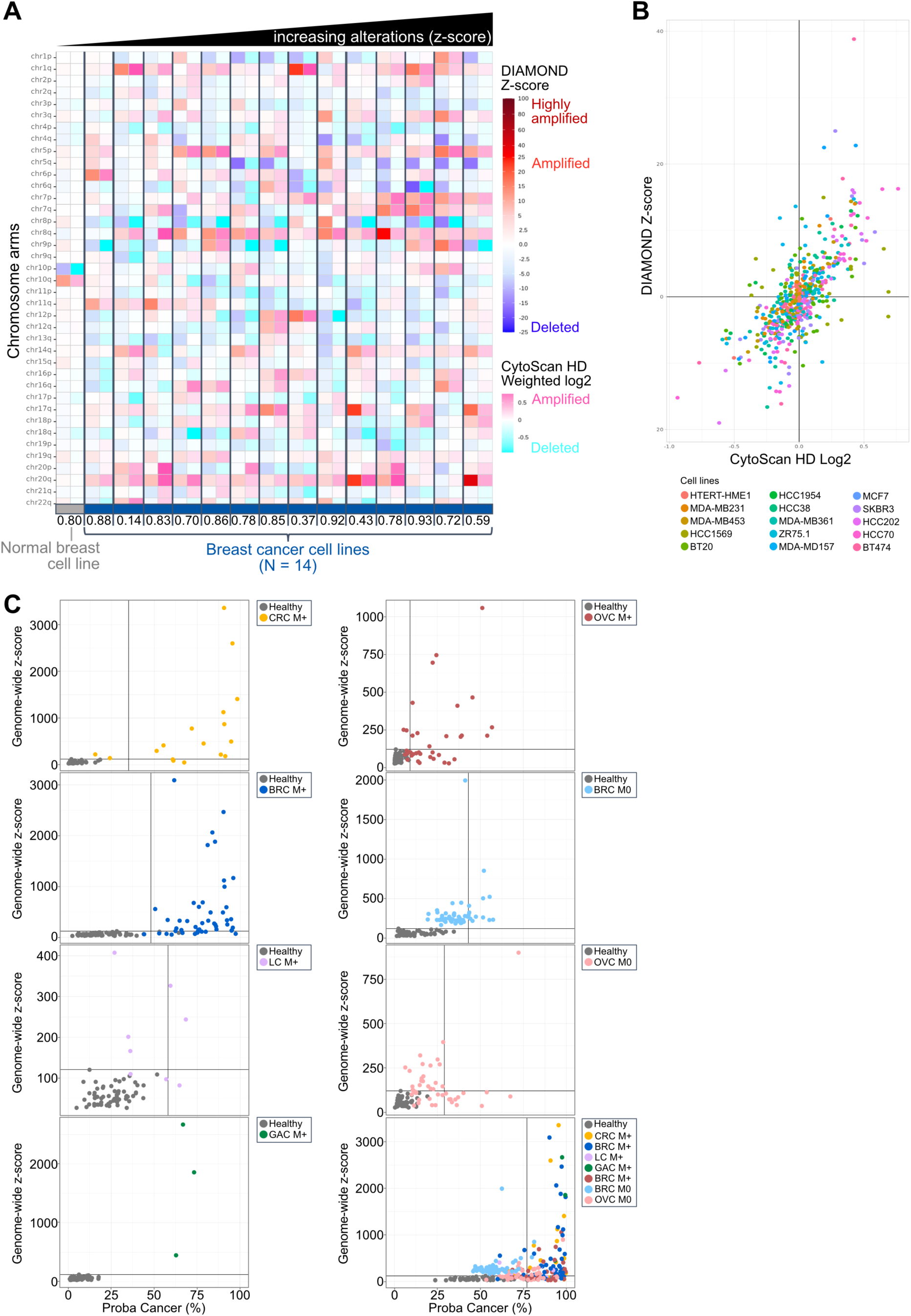
**A.** Heatmap comparing copy number alterations (CNA) profiled by DIAMOND Z-score versus Cytoscan HD microarrays for 1 normal-like breast cell line (HTERT-HME1) and 14 breast cancer cell lines (in order: MDA-MB231, MDA-MB453, HCC1569, BT20, HCC1954, HCC38, MDA-MB361, ZR 75.1, MDA-MB157, MCF7, SKBR3, HCC202, HCC70, BT474) organized by increasing aneuploidy measured by the chromosome arms z-scores. Correlation scores are indicated below the heatmap. **B.** Correlation between CNA measured by CytoScan HD microarrays and DIAMOND, r_overall_ = 0.68, p = 8.73e-80. **C**. CNA as a function of the probability of a sample to be classified as cancer (Proba Cancer) in the validation models. Graphs for cancer subgroups and all cancers together are shown.

**Fig S10.**
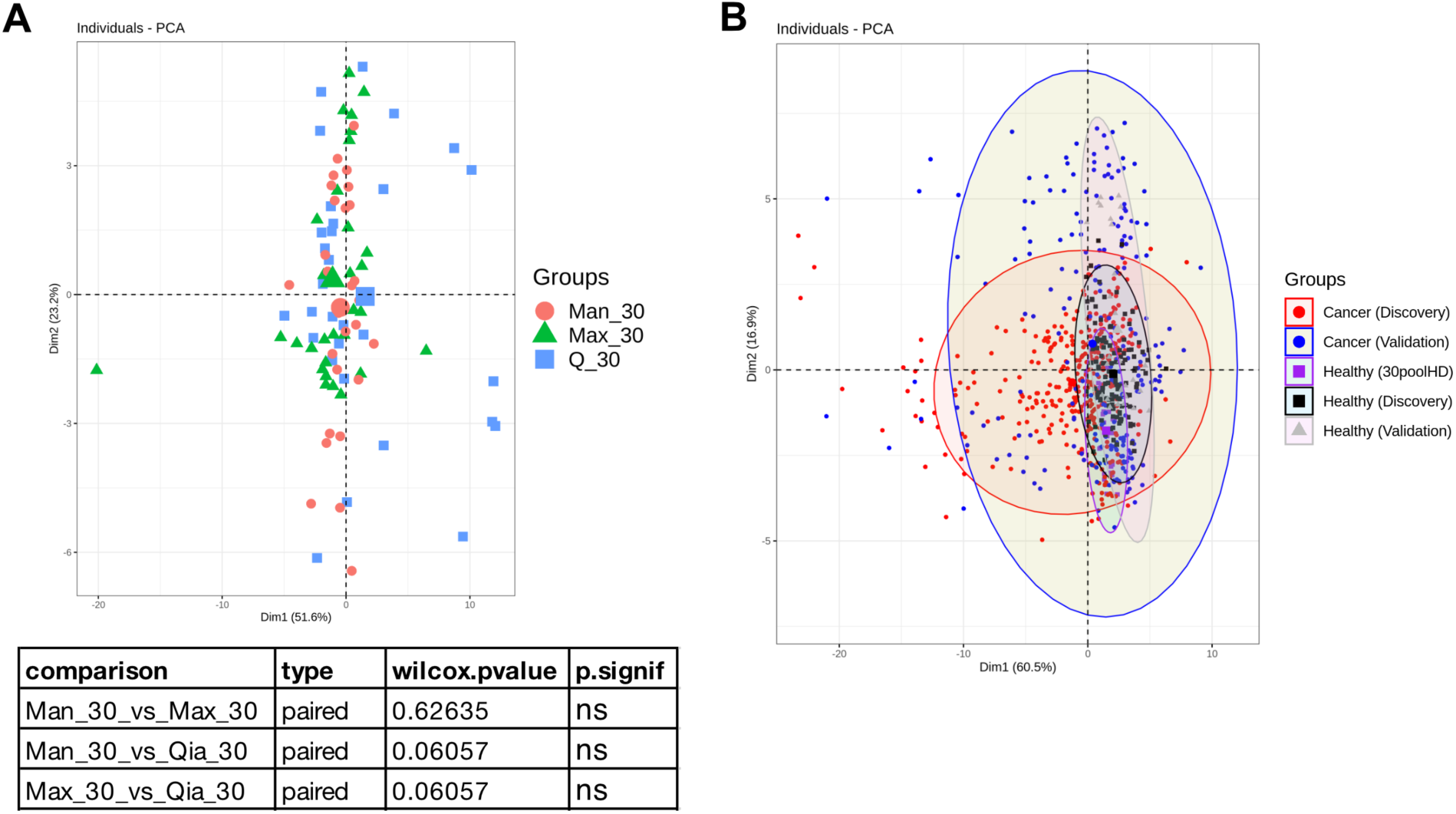
**A.** Principal component analysis (PCA), based on the average methylation level at each CpG position (n=30), showing the distribution of 30 pools of healthy plasma which were each divided into 3 replicates extracted in parallel with the Qiasymphony (Q), Maxwell (Max) and the Manual (Man) kits. The table shows the results of Wilcoxon tests demonstrating no significant differences between the 3 extraction methods. **B.** PCA showing the distribution of the various batches of healthy donors compared to cancer samples of C1 and C2 highlighting that the experimental effect that can be observed between various library construction batches are distributed along dimension 2 when the disease status effect appears in dimension 1 (see also **Fig. S2C, E**).

**Fig S11.**
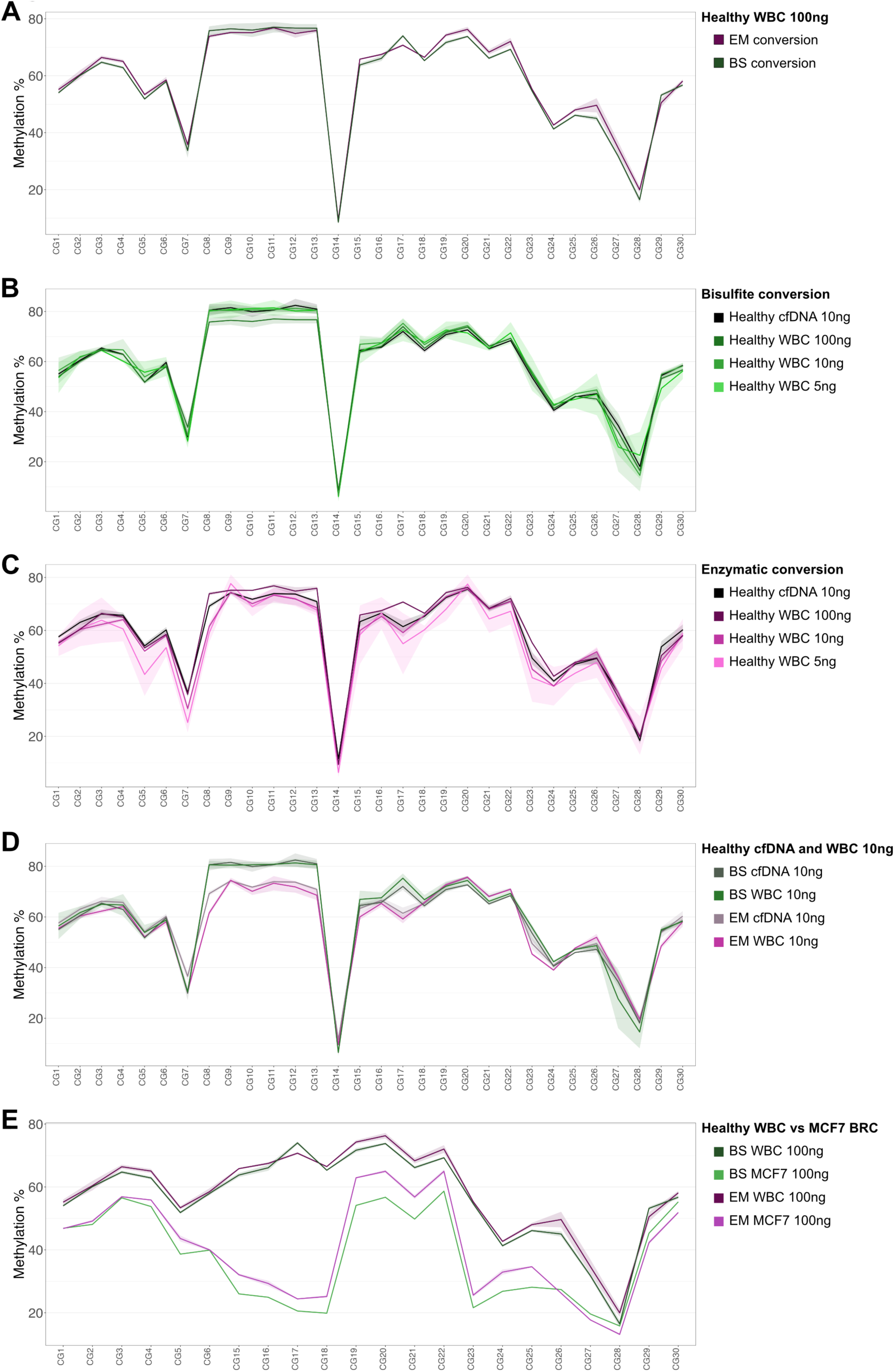
**A-E.** Comparison of the methylation level at individual CpG sites across L1 for samples treated with enzymatic (EM) versus bisulfite (BS) conversion followed by amplification with the DIAMOND targets and deep sequencing or with various DNA starting quantities. **A.** 100ng DNA from healthy white blood cells (WBC) treated with EM versus BS conversion. **B.** 10ng healthy cfDNA vs various DNA input quantities from WBC converted with BS. **C.** 10ng healthy cfDNA vs various DNA input quantities from WBC converted with EM. **D.** 10ng cfDNA or WBC DNA treated with BS versus EM conversion. **E.** DNA from healthy WBC versus MCF7 breast cancer cell lines treated with EM versus BS conversion.

---

**Table S1.**
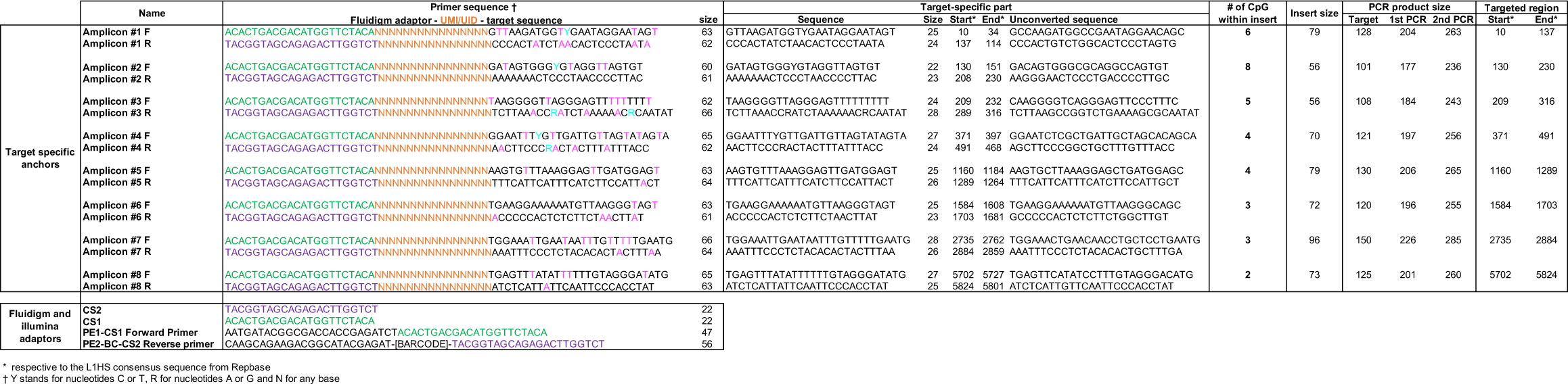
Targeted bisulfite sequencing primers.

**Table S2.** Target copies and CpG sites relative to the number of sequencing reads.

| Sample | Biological class | # of reads per sample | Deduplicated length filtered inserts | # of CpG sites covered | # of bp covered | # of L1 copies with unique mappers | # of L1 copies with random mappers | # of L1 copies with unique and random mappers |
| --- | --- | --- | --- | --- | --- | --- | --- | --- |
| A1072S1 | Ovarian tumor | 44 382 229 | NA | 125 137 | 6 262 236 | 27 125 | 28 921 | 40 504 |
| A1072S2 | Uveal melanoma tumor | 45 833 528 | NA | 125 072 | 6 249 451 | 27 113 | 28 817 | 40 449 |
| A1072S3 | Healthy plasma | 53 982 906 | NA | 109 700 | 5 433 427 | 24 285 | 27 596 | 37 457 |
| A1190S134 | BRC plasma | 4 637 807 | 2 756 162 | 87 826 | 4 076 579 | 17 188 | 18 430 | 29 063 |
| A1190S136 | BRC plasma | 4 071 125 | 2 455 668 | 85 416 | 3 966 291 | 16 703 | 18 093 | 28 469 |
| A1179S01 | CRC plasma | 4 335 884 | 2 815 280 | 90 001 | 4 360 428 | 19 133 | 19 843 | 31 697 |
| A1179S04 | CRC plasma | 4 885 606 | 3 162 942 | 91 262 | 4 374 819 | 19 079 | 19 614 | 31 415 |
| A1146S5 | OVC plasma | 6 632 707 | 3 912 875 | 89 757 | 4 182 581 | 17 871 | 18 553 | 29 202 |
| A1146S12 | OVC plasma | 6 785 341 | 4 454 098 | 90 343 | 4 136 316 | 17 535 | 18 269 | 28 523 |
| D473S375 | HD plasma | 5 979 285 | 1 571 767 | 84 617 | 3 846 076 | 15 888 | 17 266 | 26 811 |
| D473S376 | HD plasma | 5 933 799 | 1 458 718 | 82 466 | 3 756 894 | 15 383 | 17 016 | 26 239 |

**Table S3.** Statistical results of global methylation differences between healthy samples and cancer subgroups using Mann–Whitney U test.

| .y. | group1 | group2 | n1 | n2 | p | p.signif | Figure |
| --- | --- | --- | --- | --- | --- | --- | --- |
| Global methylation | Healthy WBC | Fully methylated controls | 14 | 13 | 0.000001 | **** | 2A |
| Global methylation | Healthy WBC | Unmethylated control | 14 | 12 | 0.000002 | **** | 2A |
| Global methylation | Healthy WBC | Colorectal_cancer_cellLine | 14 | 1 | 0.266000 | ns | 2A |
| Global methylation | Healthy WBC | Ovarian_cancer_cellLine | 14 | 3 | 0.012000 | * | 2A |
| Global methylation | Healthy WBC | Breast_cancer_cellLine | 14 | 15 | 0.000002 | **** | 2A |
| Global methylation | Healthy WBC | Uveal_melanoma_cellLine | 14 | 13 | 0.000068 | **** | 2A |
| Global methylation | Healthy WBC | Healthy_ovarian_tissue | 14 | 11 | 0.063000 | ns | 2A |
| Global methylation | Healthy WBC | Ovarian_cancer_tissue | 14 | 28 | 0.008000 | ** | 2A |
| Global methylation | Healthy WBC | Breast_cancer_tissue | 14 | 25 | 0.000410 | *** | 2A |
| Global methylation | Healthy WBC | Uveal_cancer_tissue | 14 | 24 | 0.880000 | ns | 2A |
| Global methylation | Healthy plasmas | BRC M+ plasmas | 123 | 97 | 0.000000 | **** | 2B |
| Global methylation | Healthy plasmas | BRC M0 plasmas | 123 | 23 | 0.910000 | ns | 2B |
| Global methylation | Healthy plasmas | CRC M+ plasmas | 123 | 75 | 0.000000 | **** | 2B |
| Global methylation | Healthy plasmas | LC M+ plasmas | 123 | 50 | 0.655000 | ns | 2B |
| Global methylation | Healthy plasmas | OVC M0 plasmas | 123 | 18 | 0.000019 | **** | 2B |
| Global methylation | Healthy plasmas | GAC M0 plasmas | 123 | 27 | 0.000000 | **** | 2B |
| Global methylation | Healthy plasmas | UVM M+ plasmas | 123 | 55 | 0.000008 | **** | 2B |

**Table S4.**
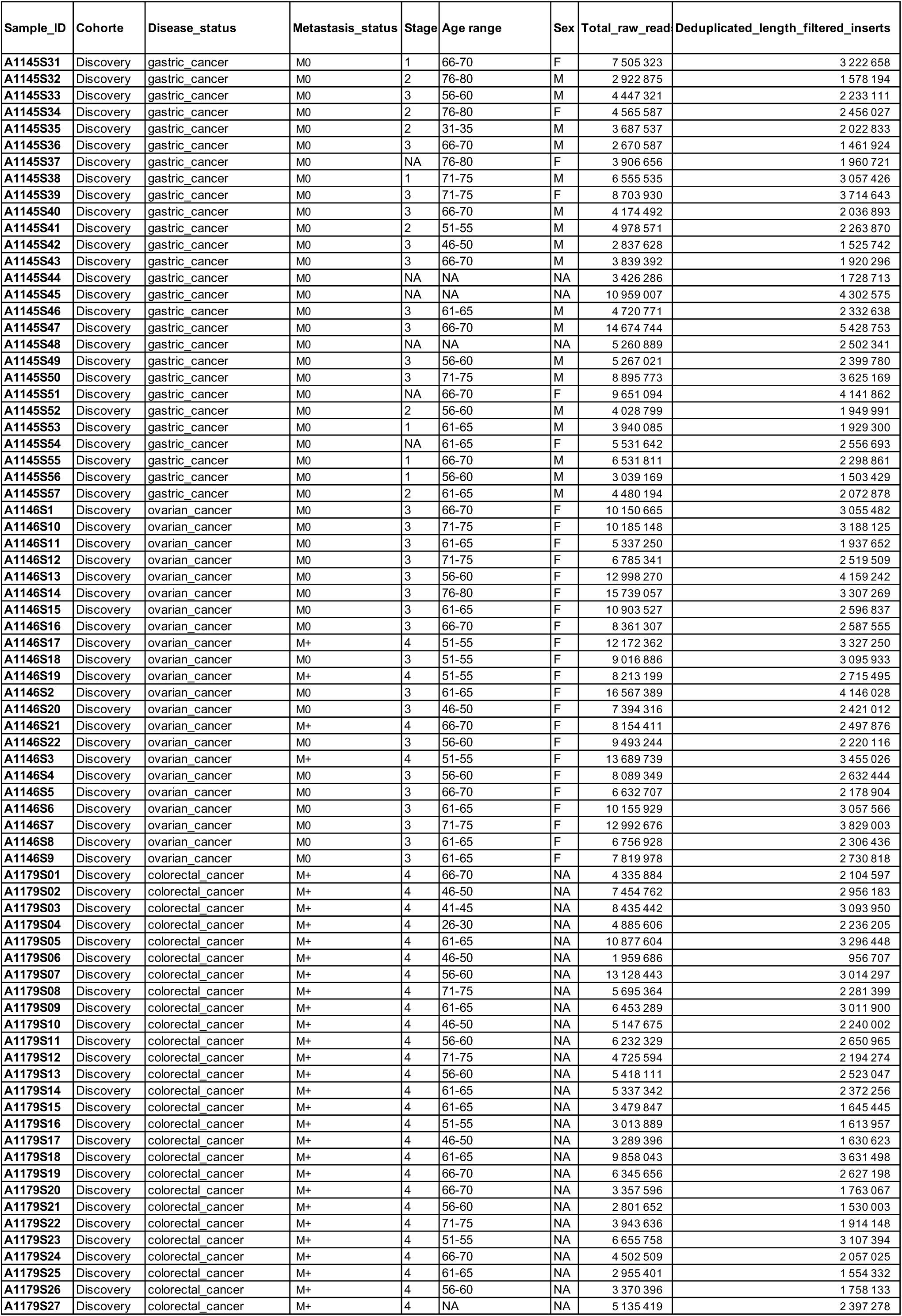
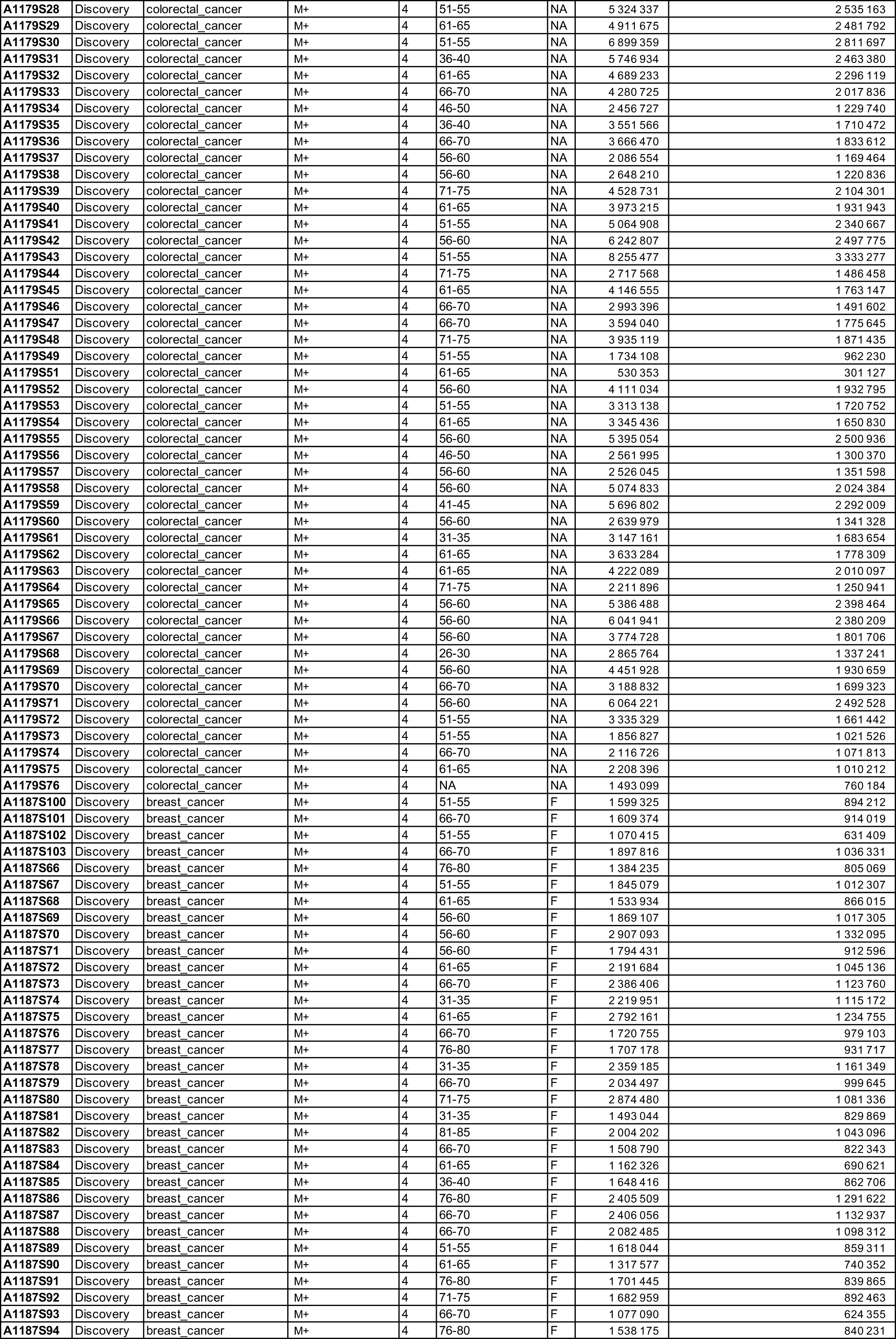
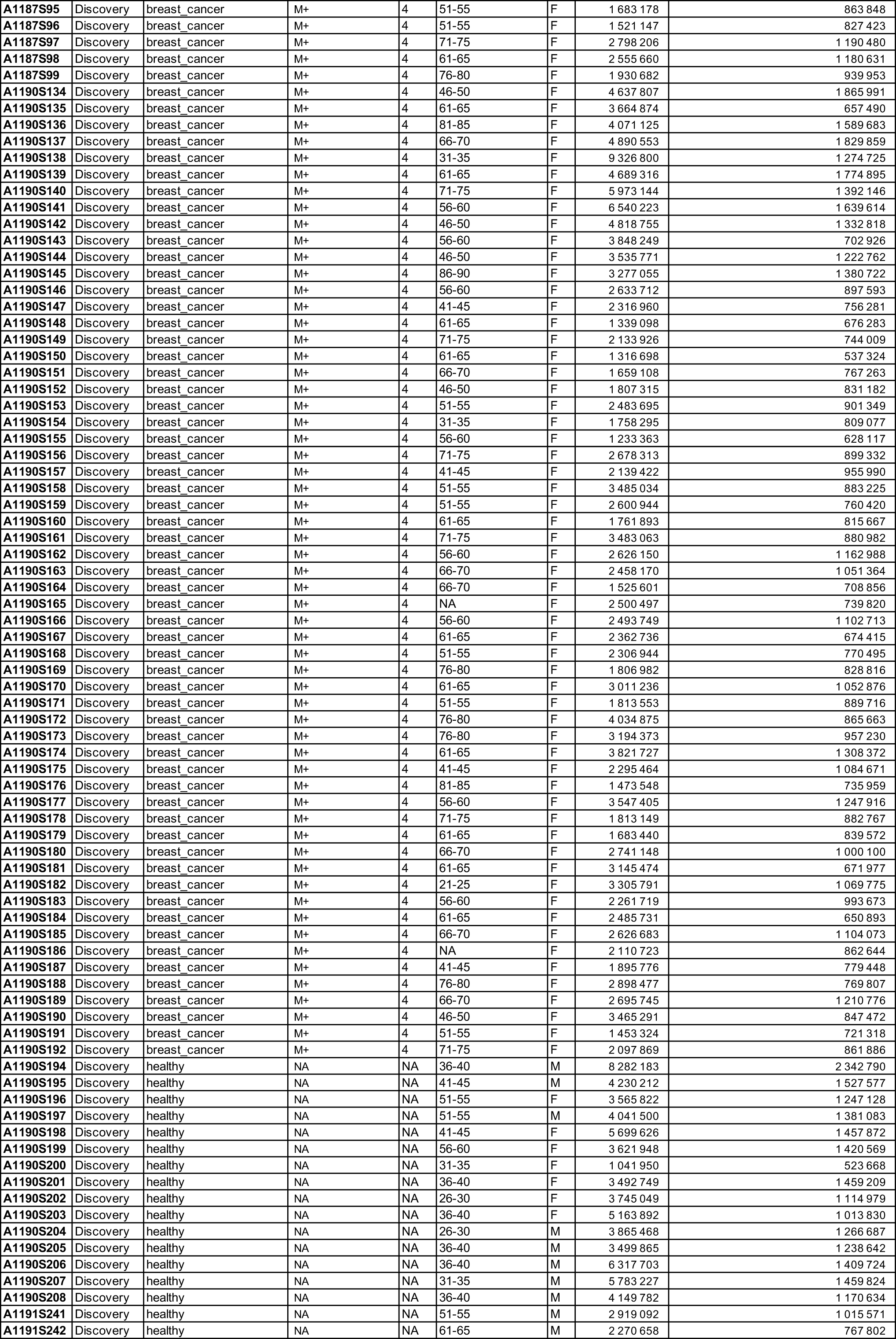
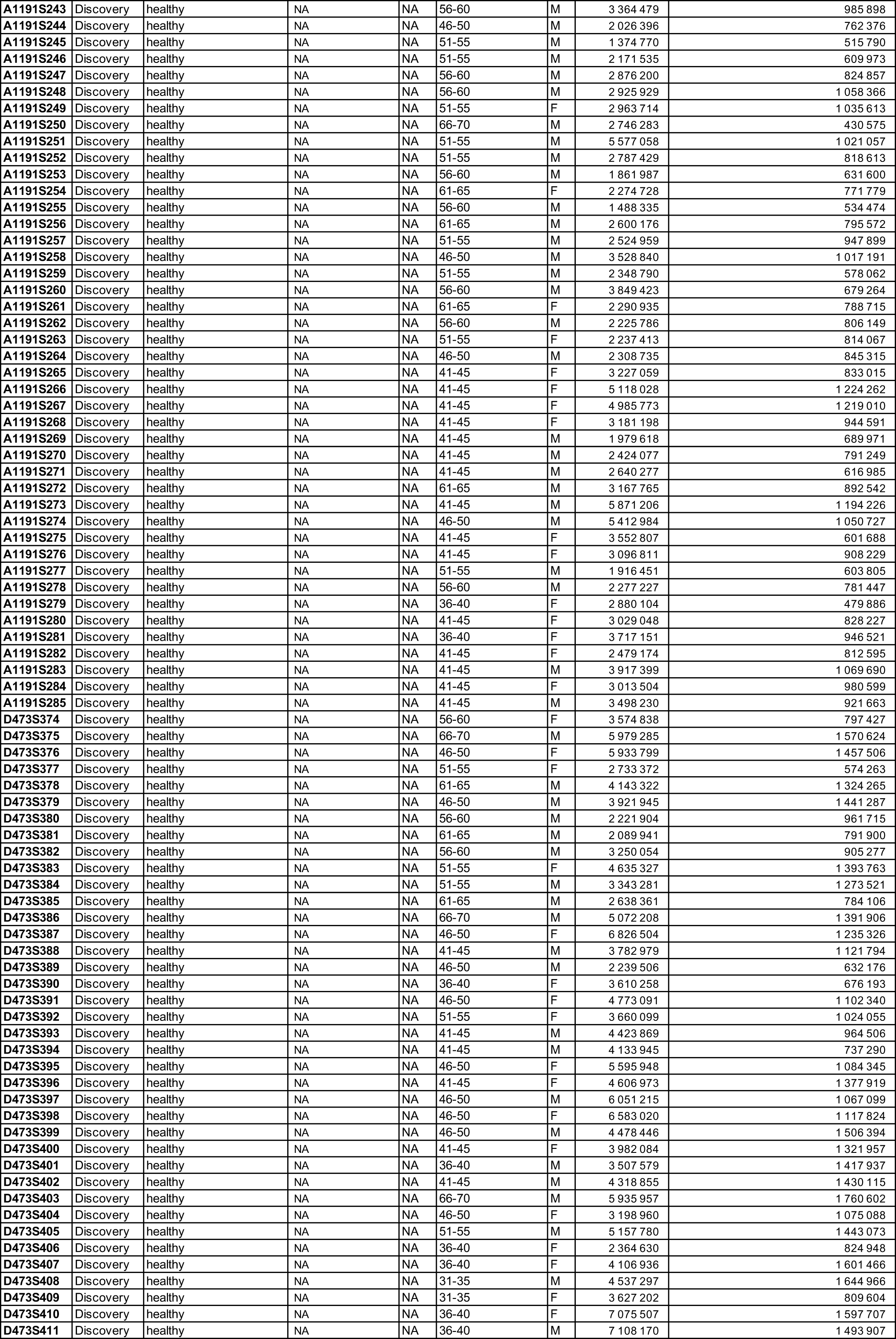
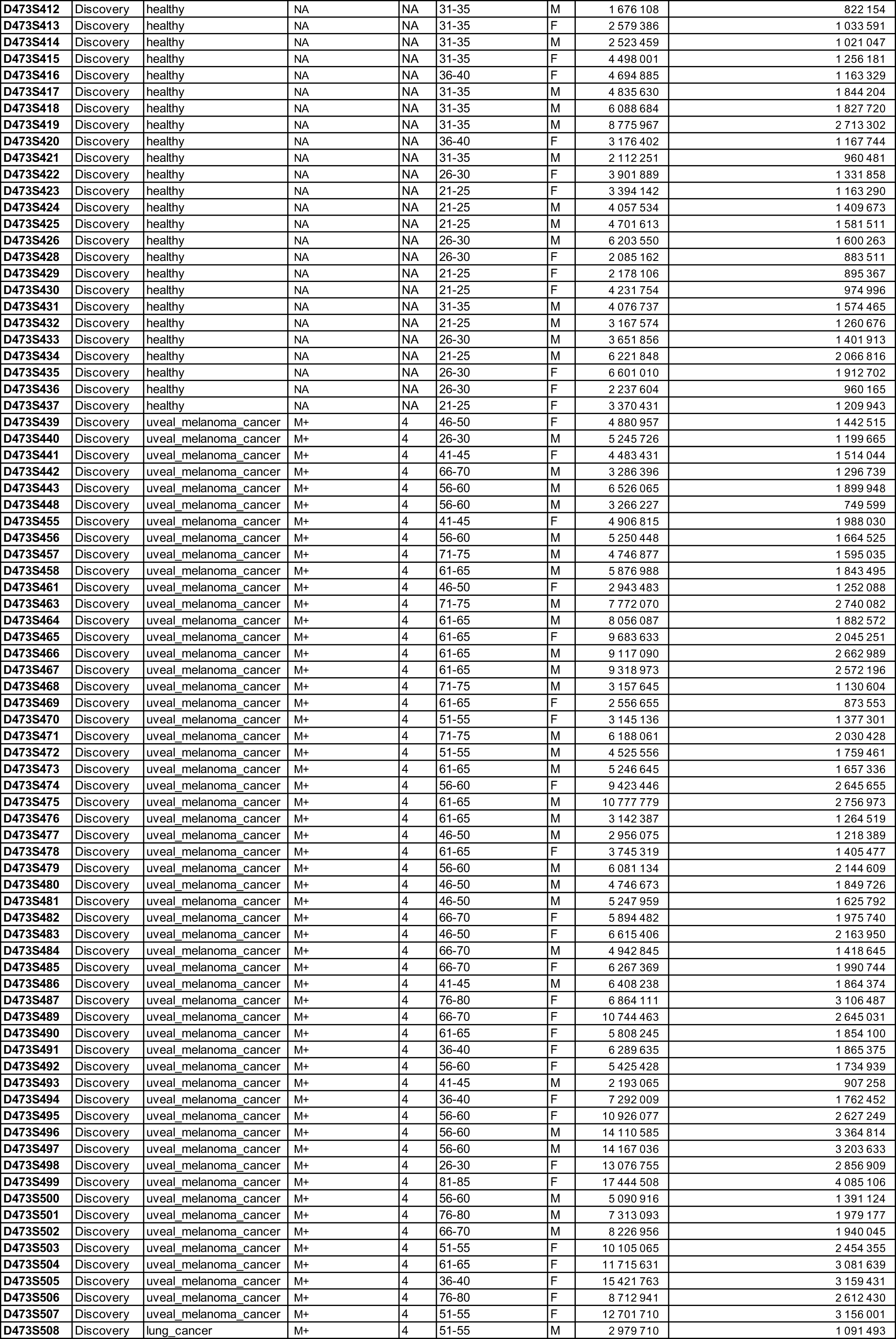
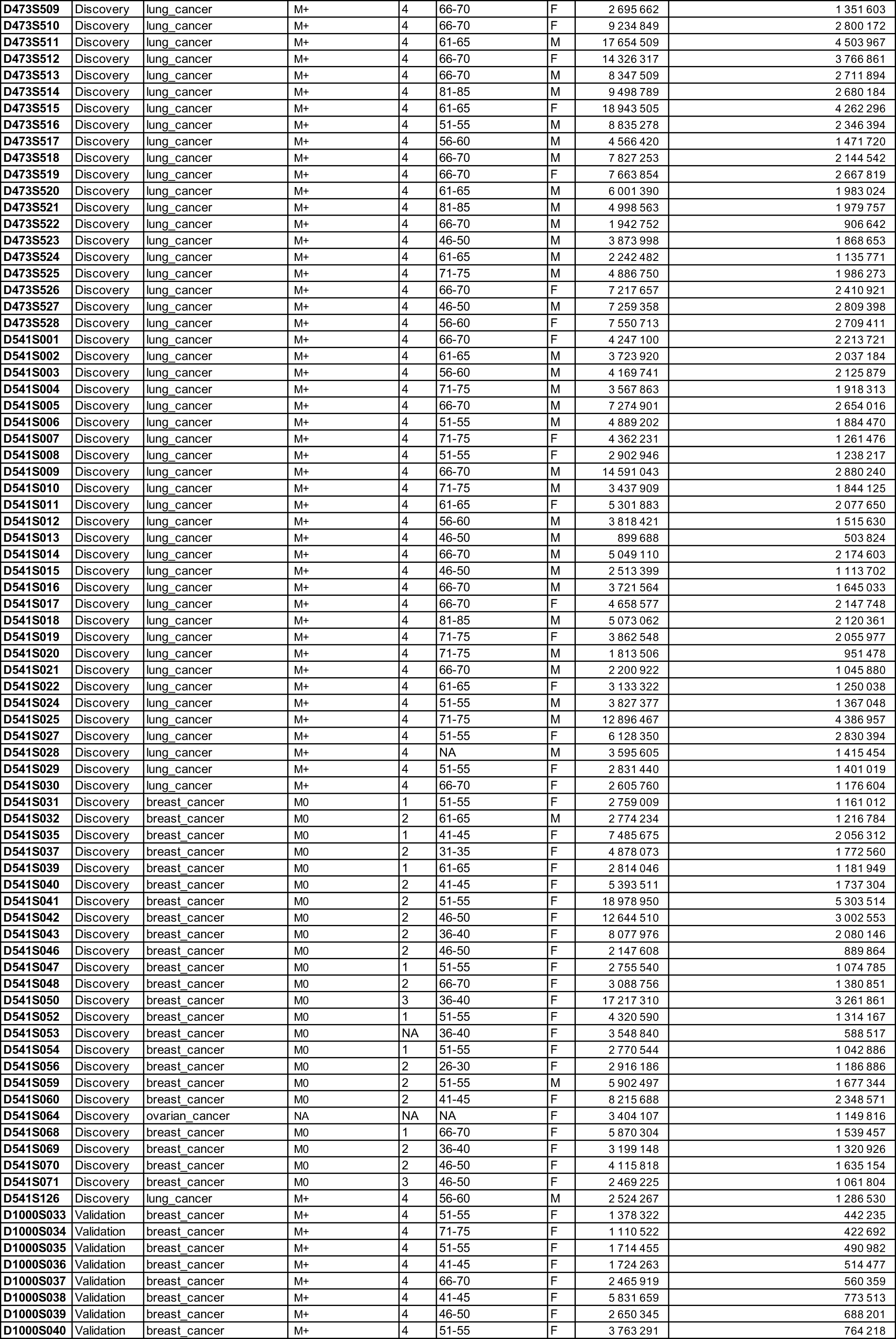
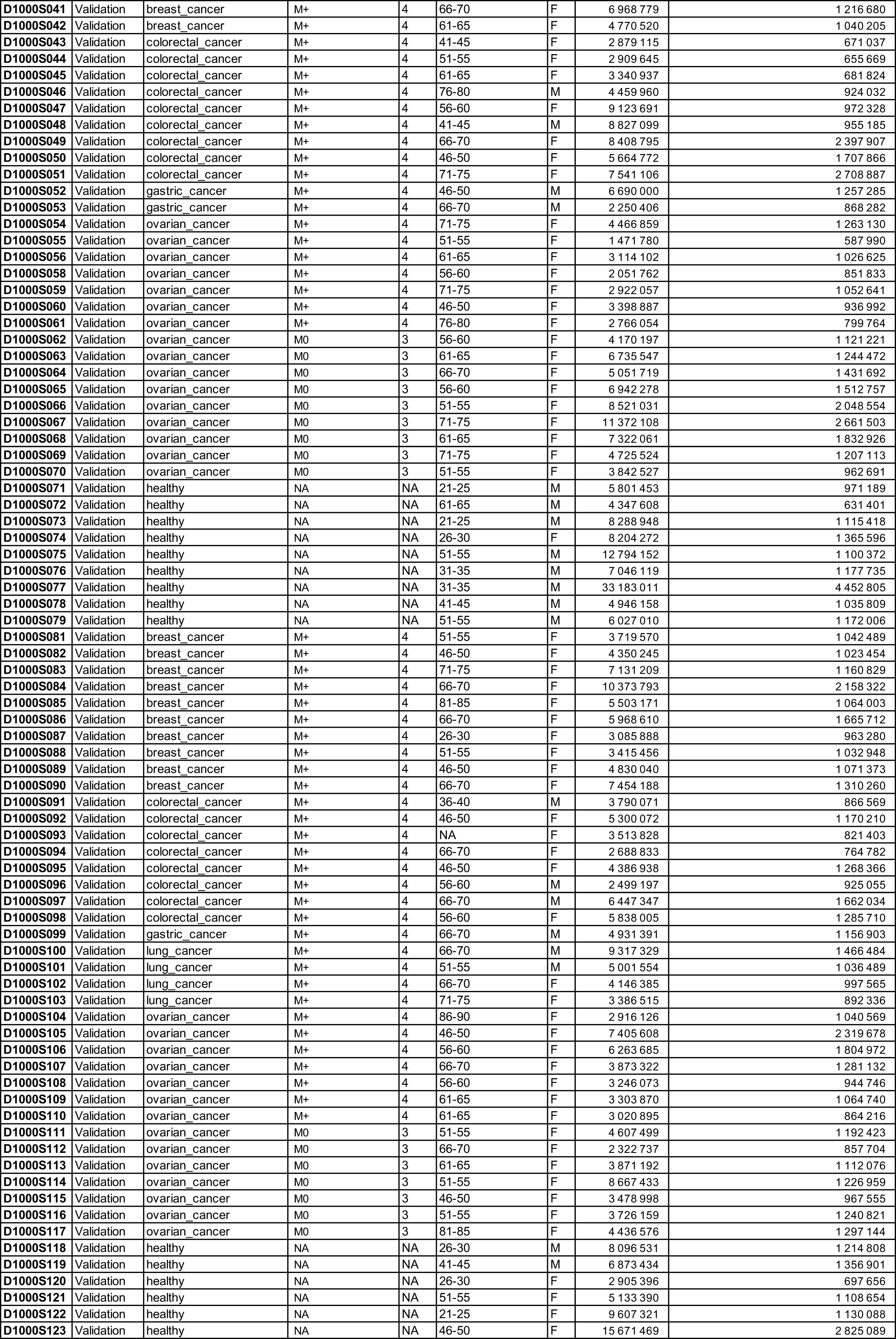
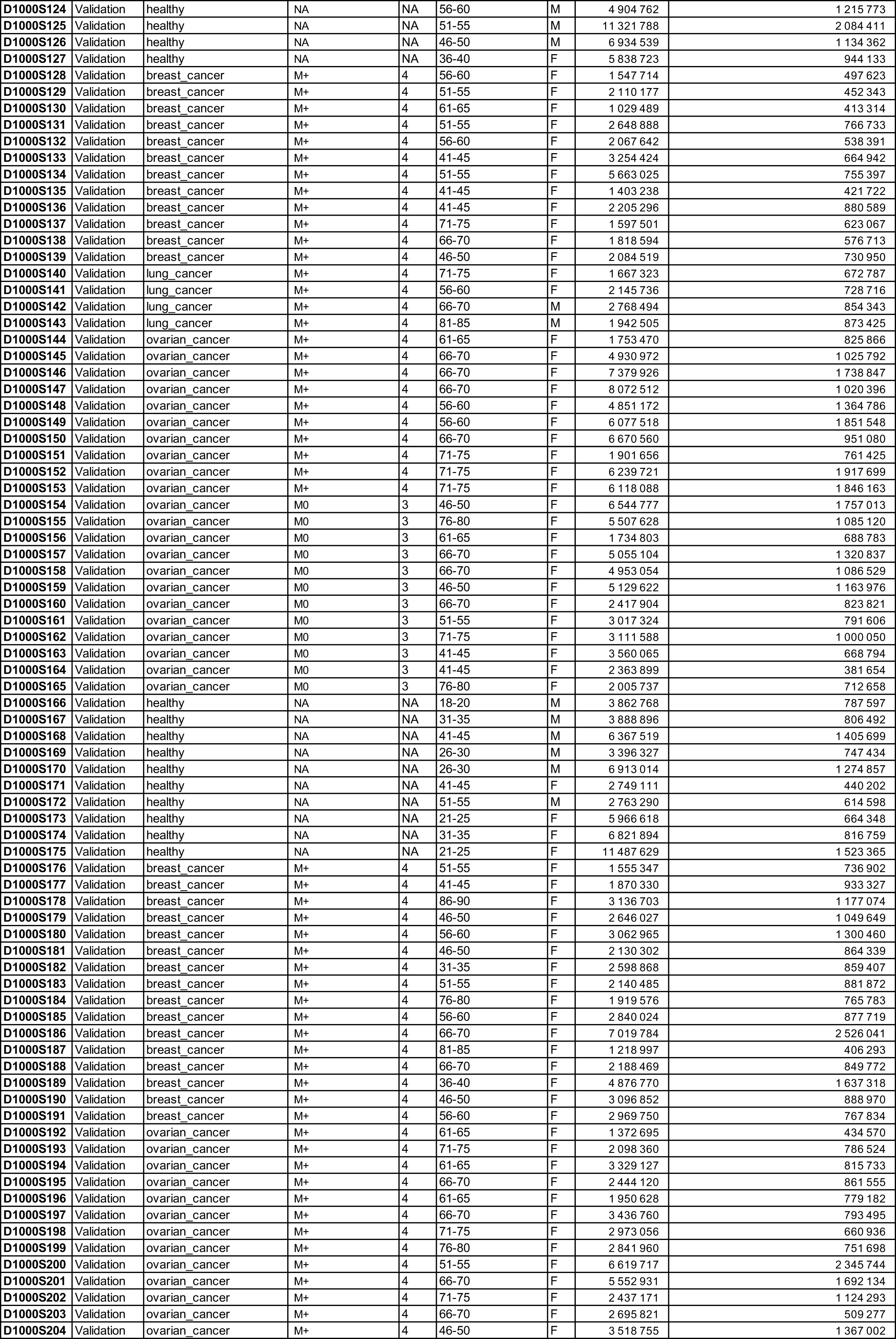
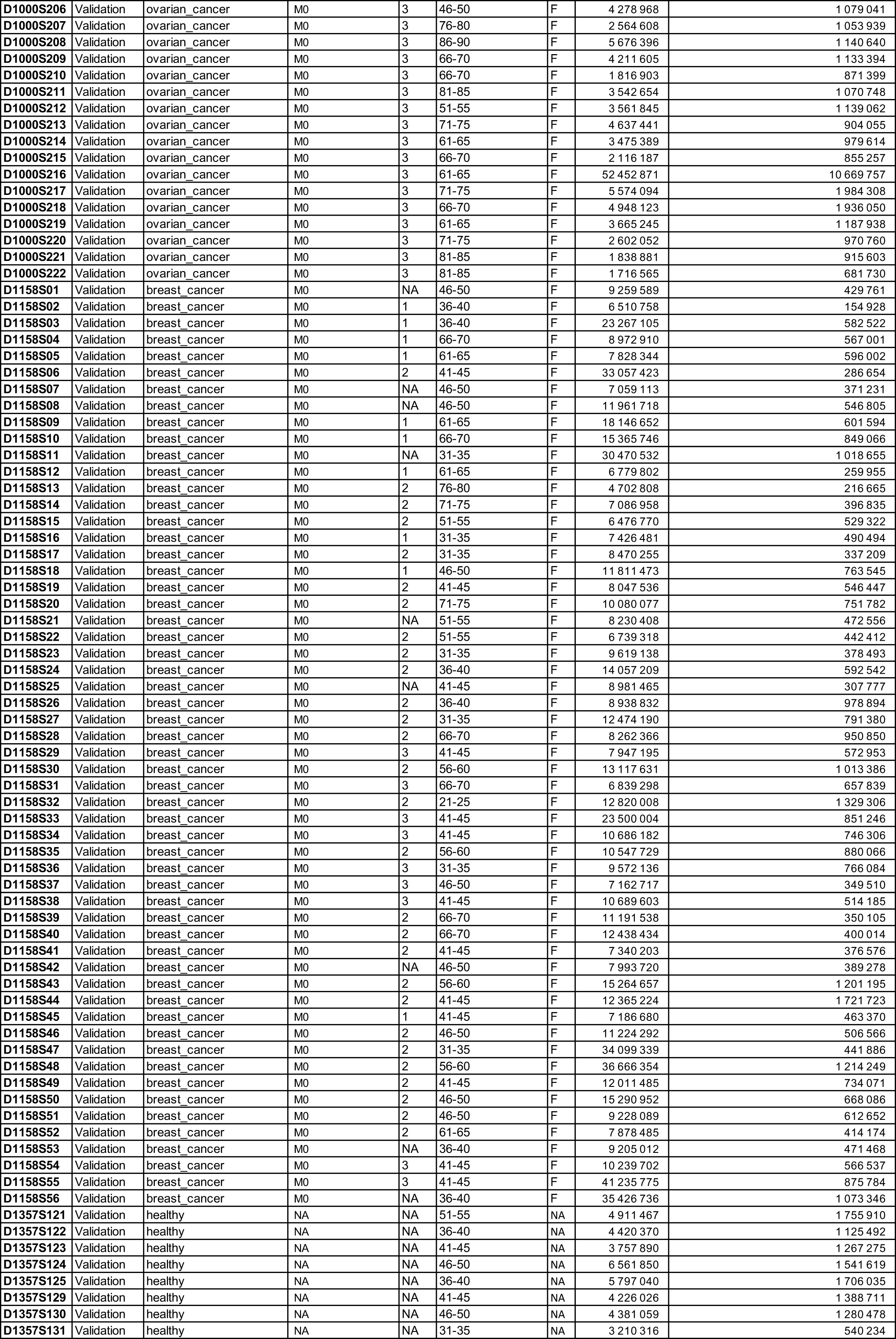
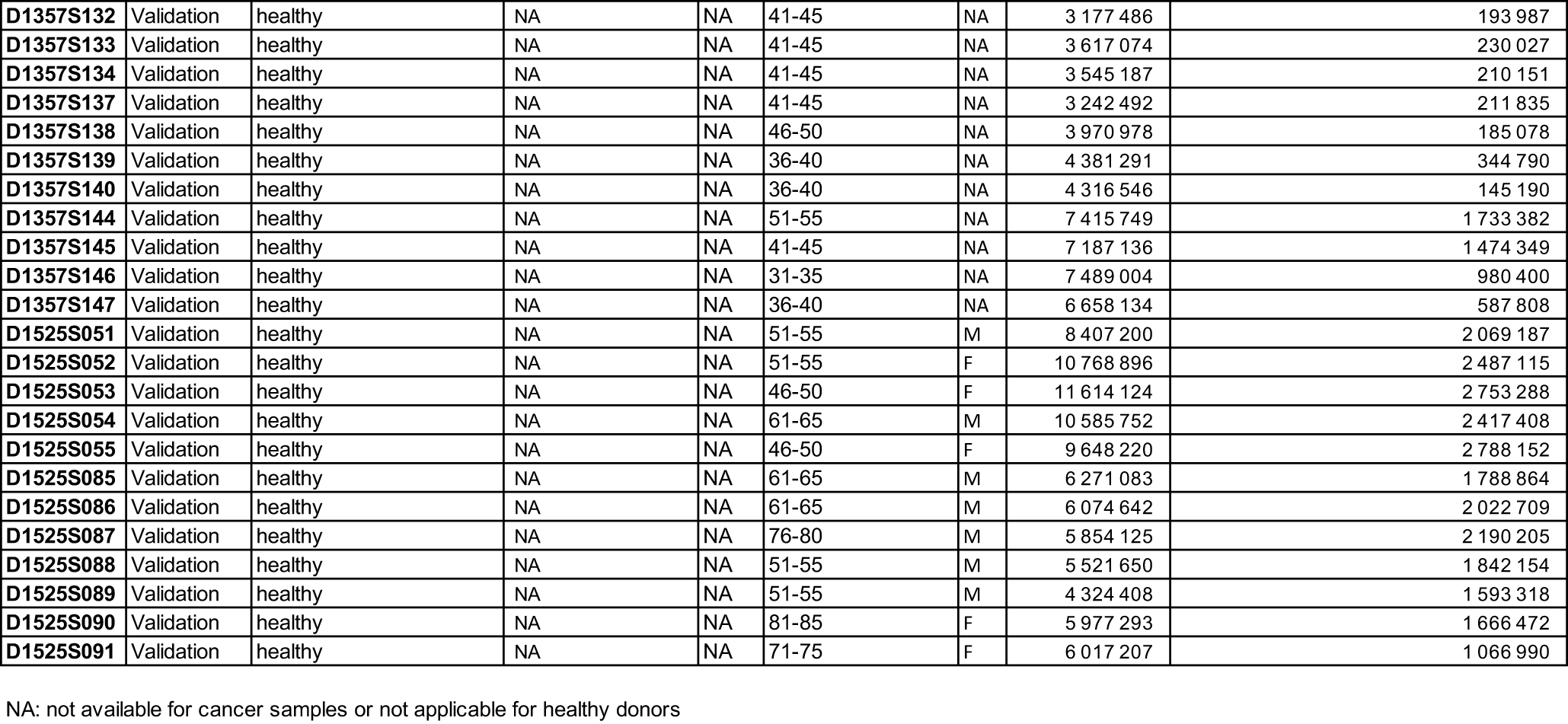
List of plasma samples. Sample IDs were not known to anyone outside the research group.

**Table S5.**
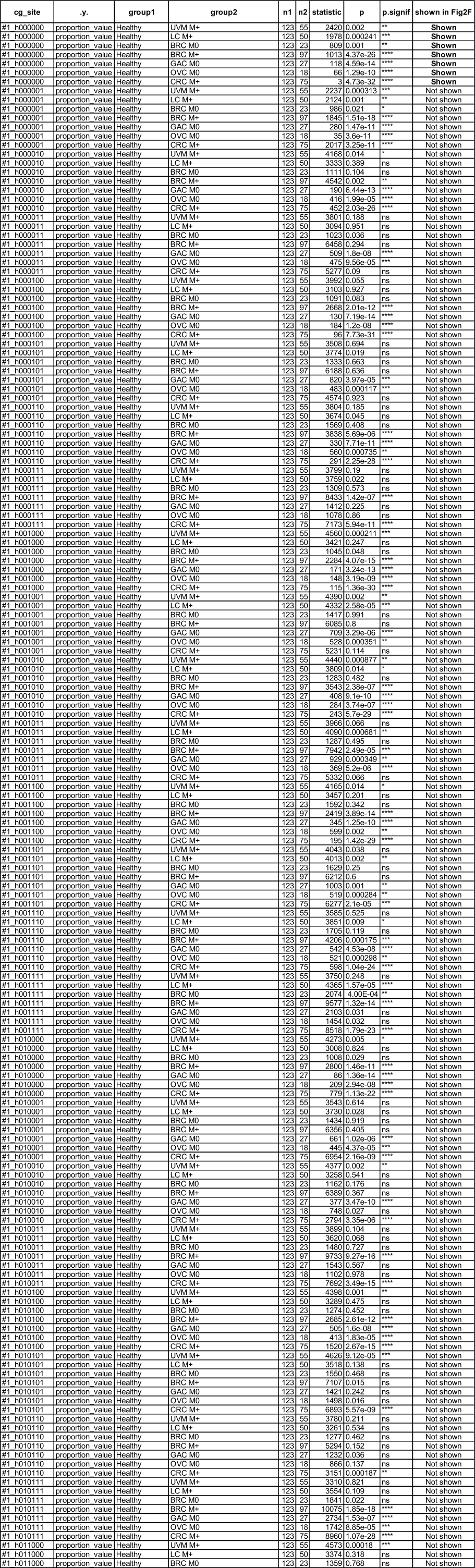
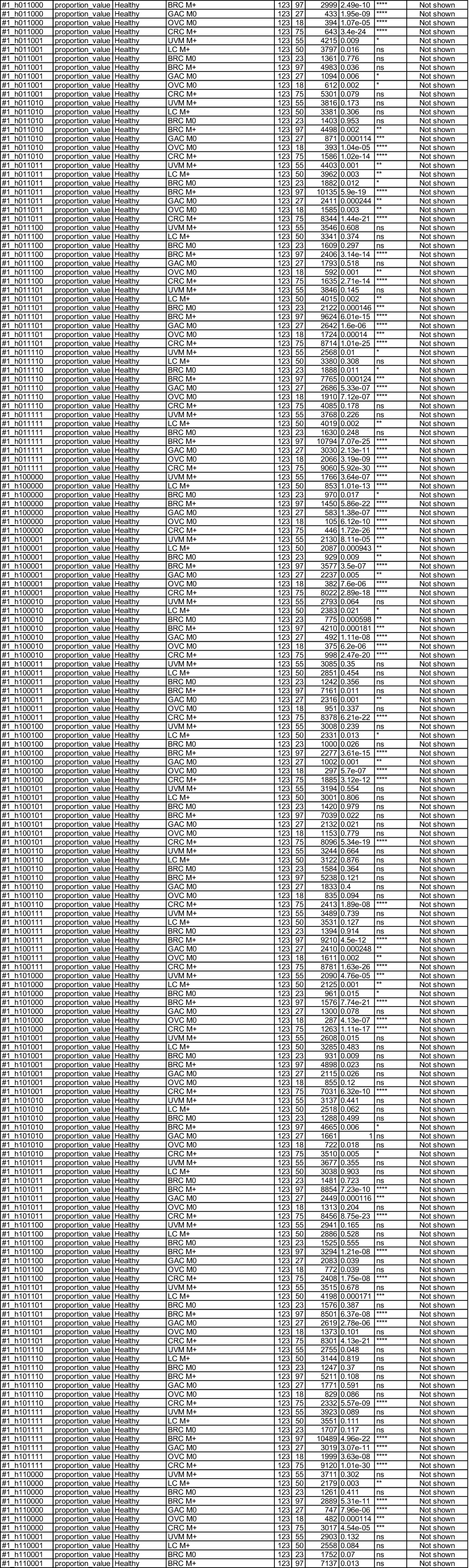
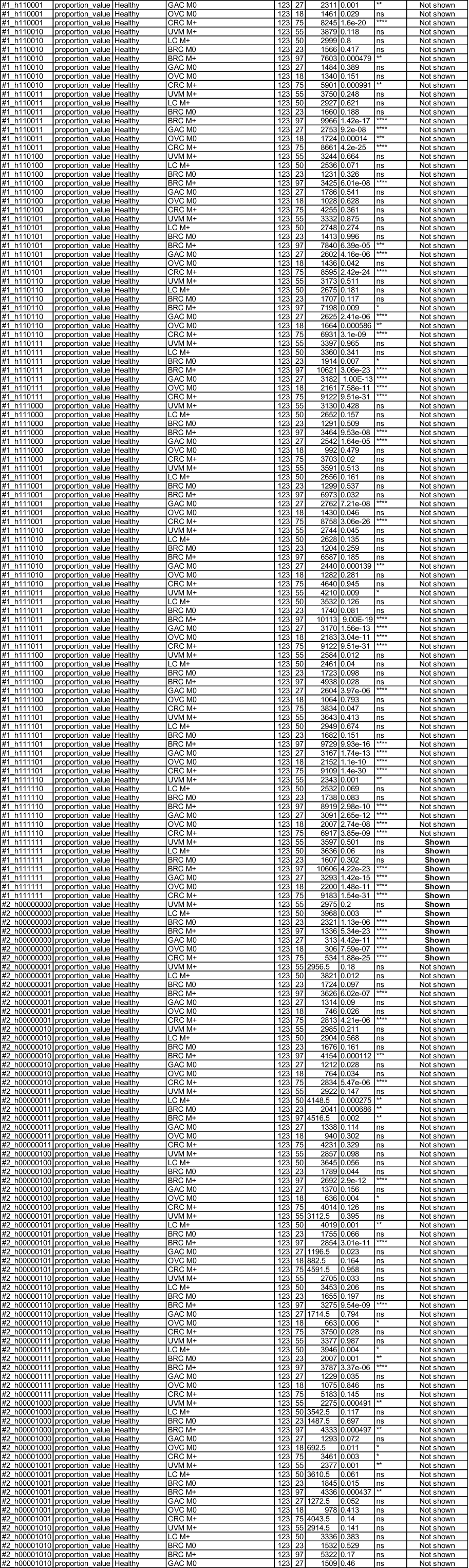
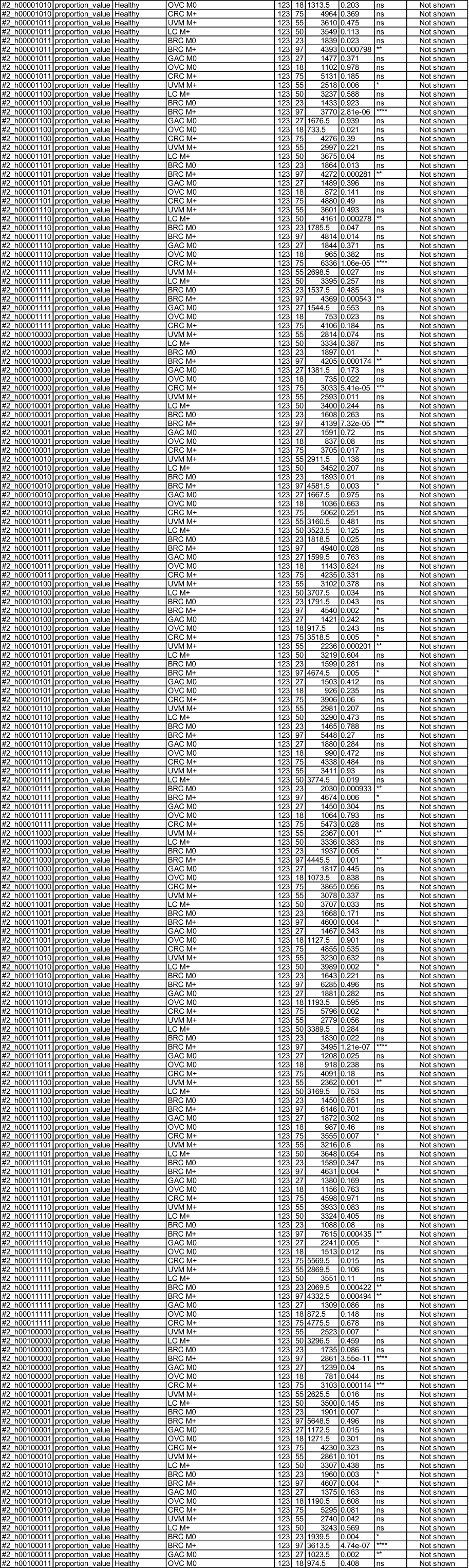
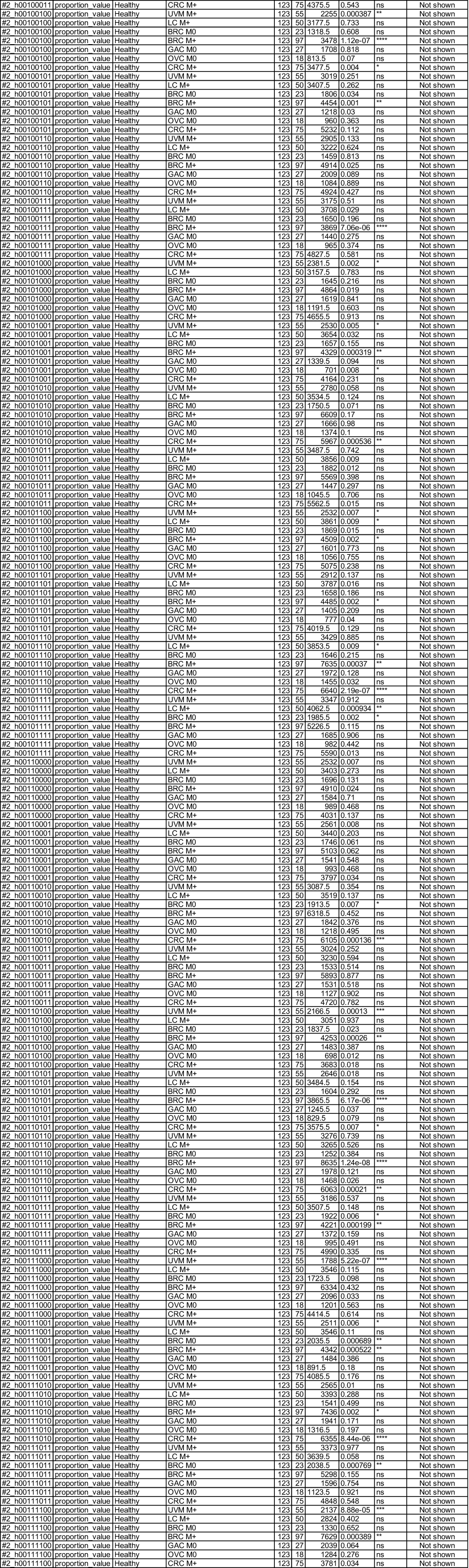
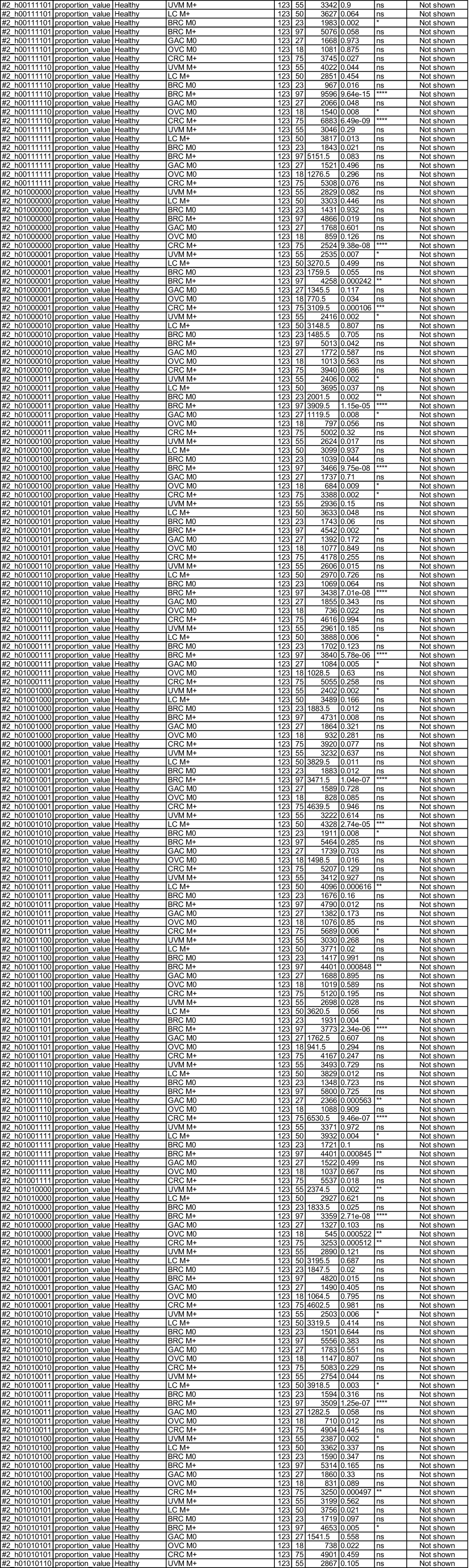
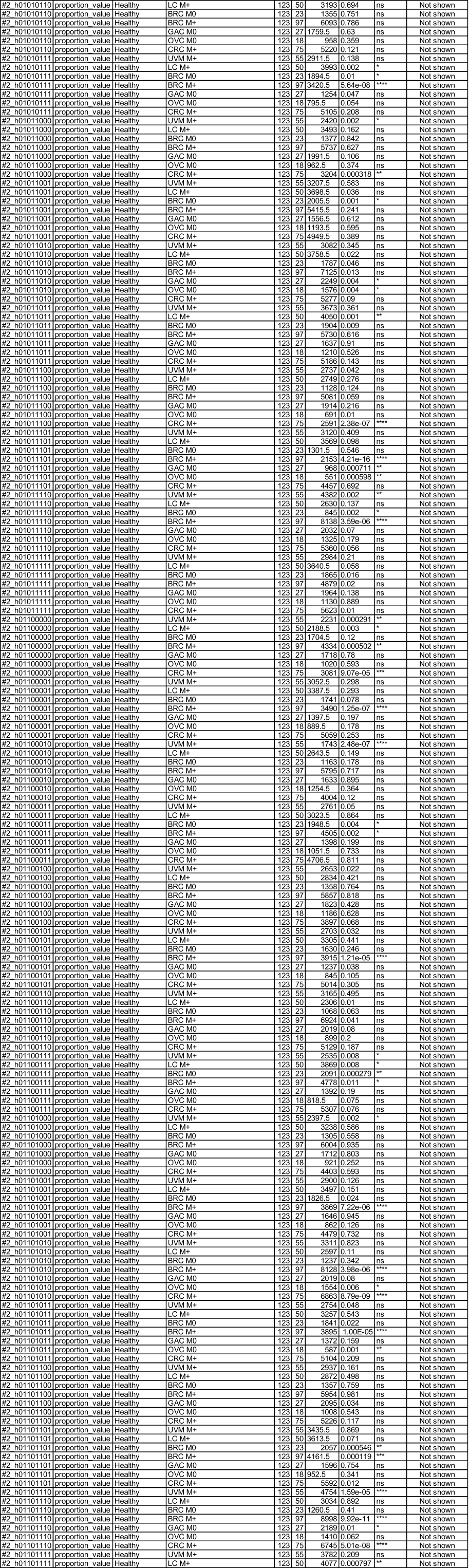
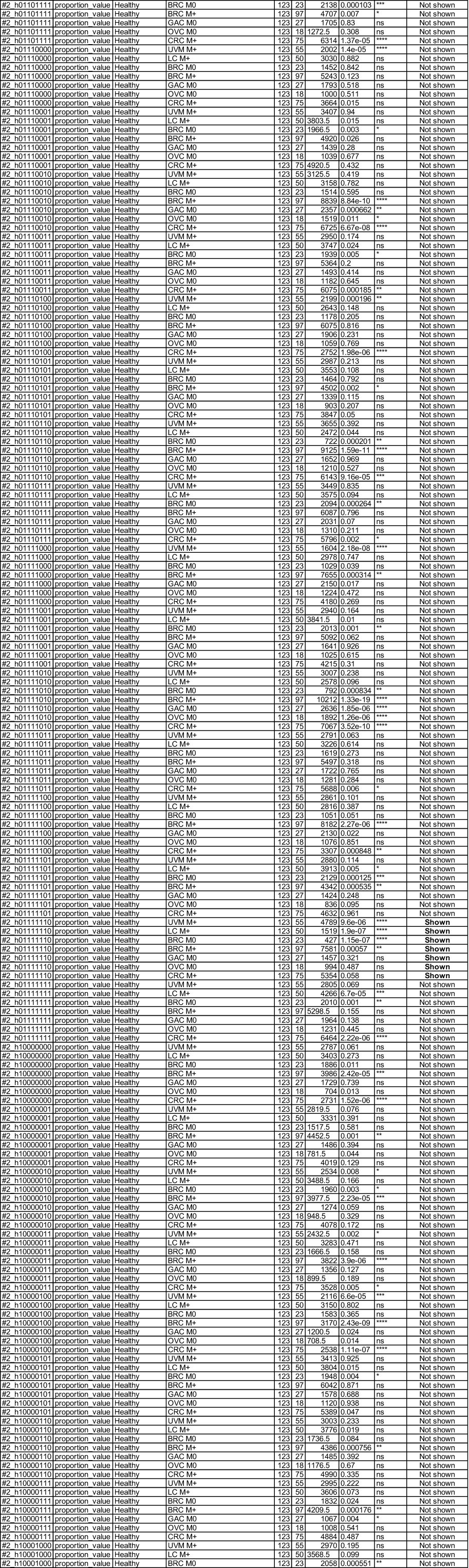
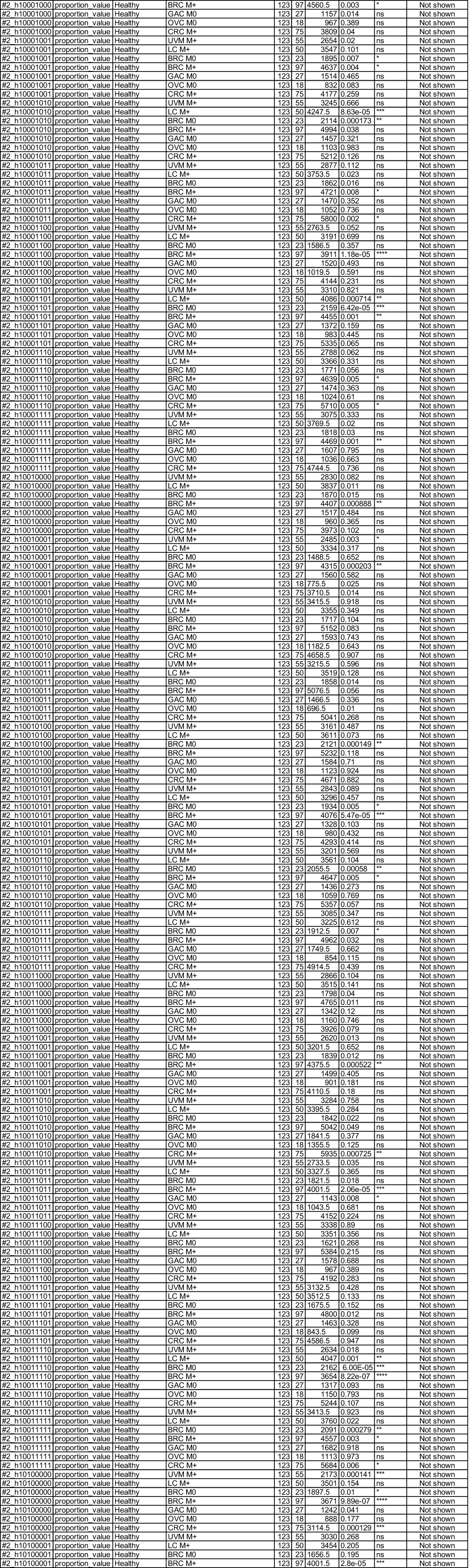
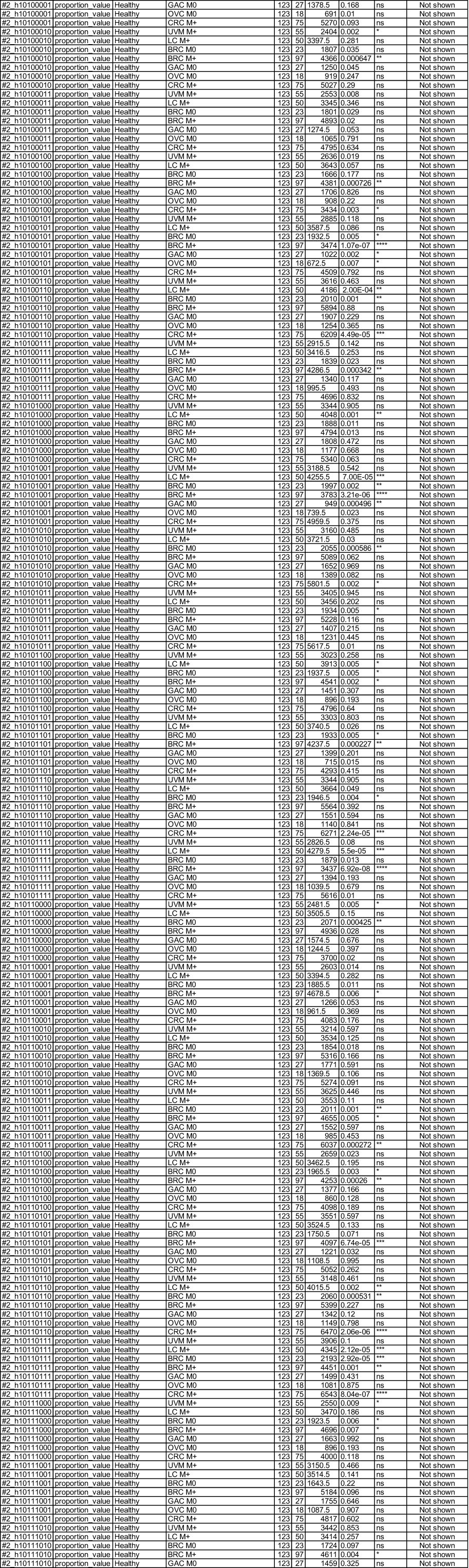
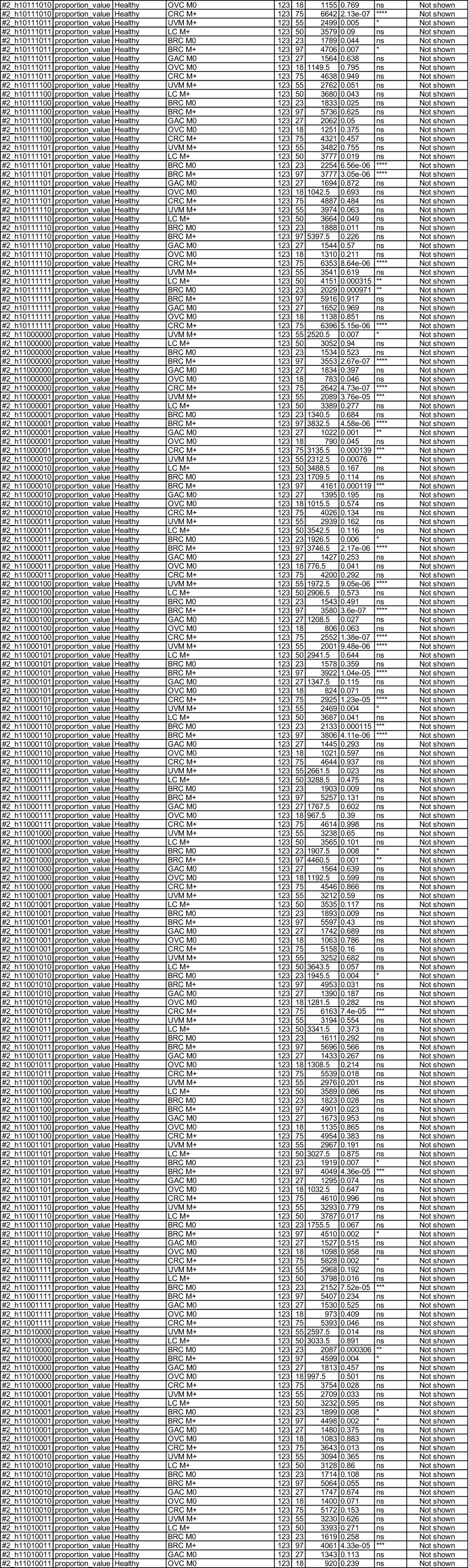
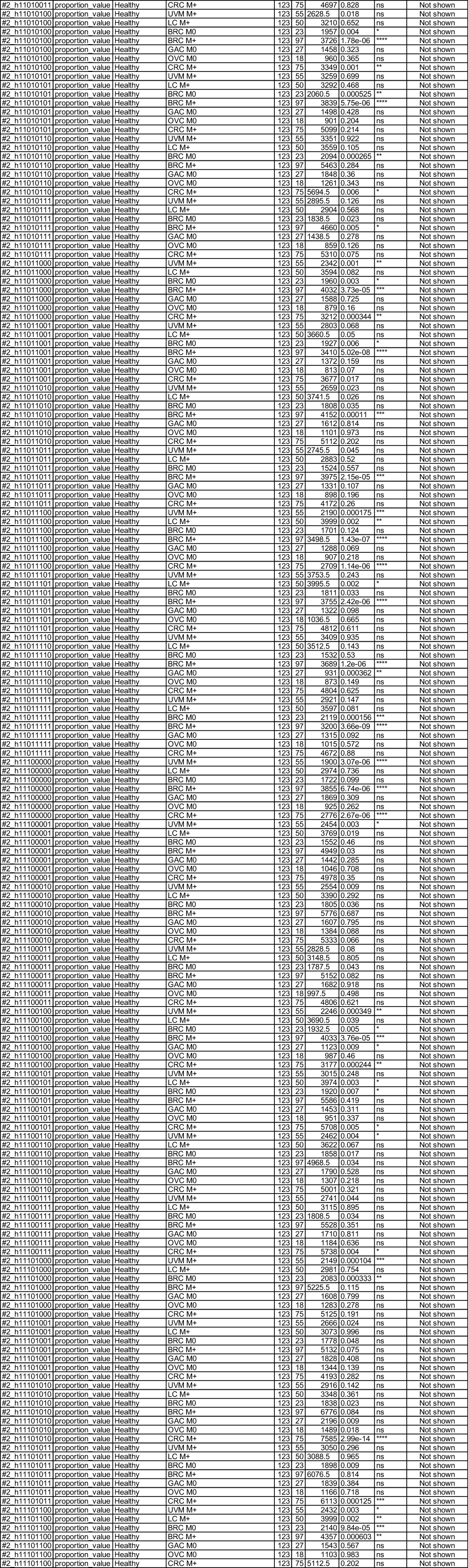
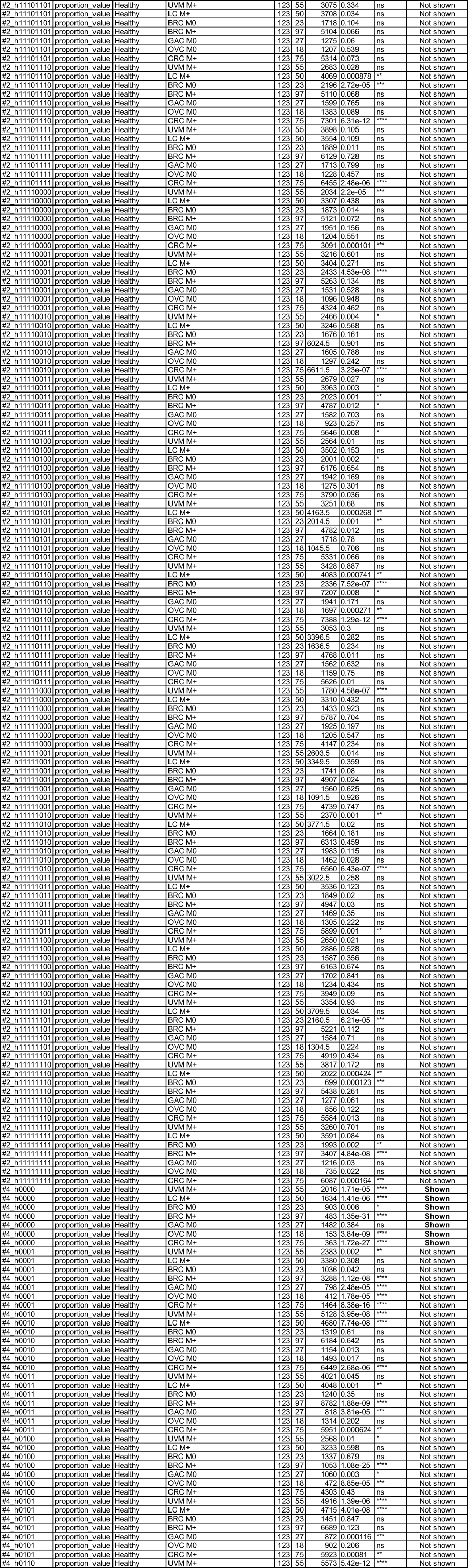
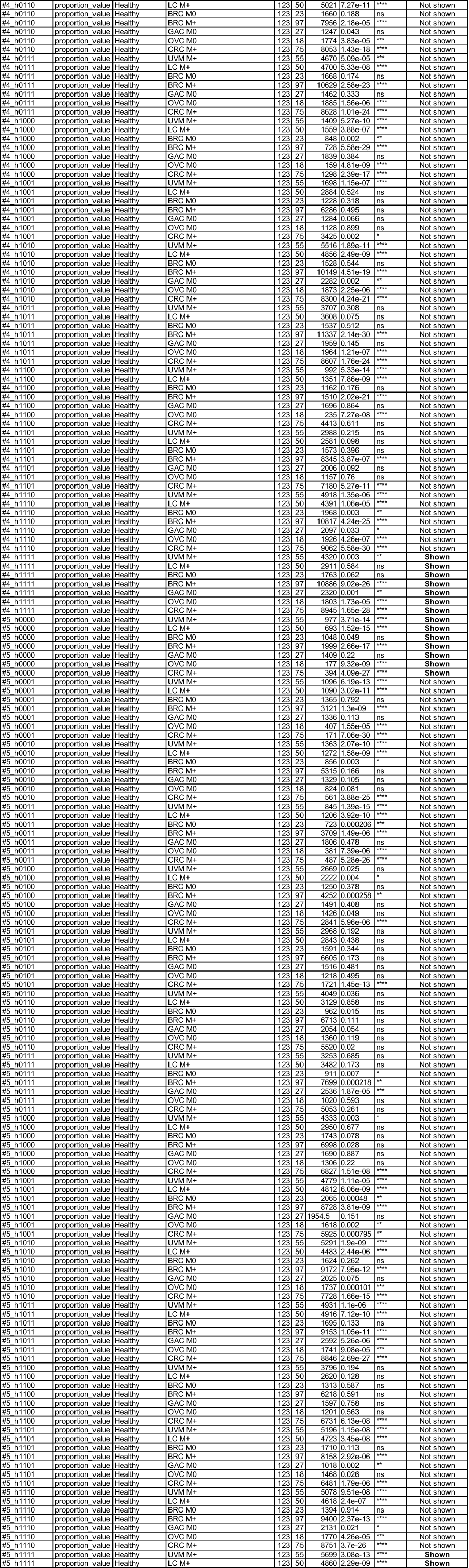
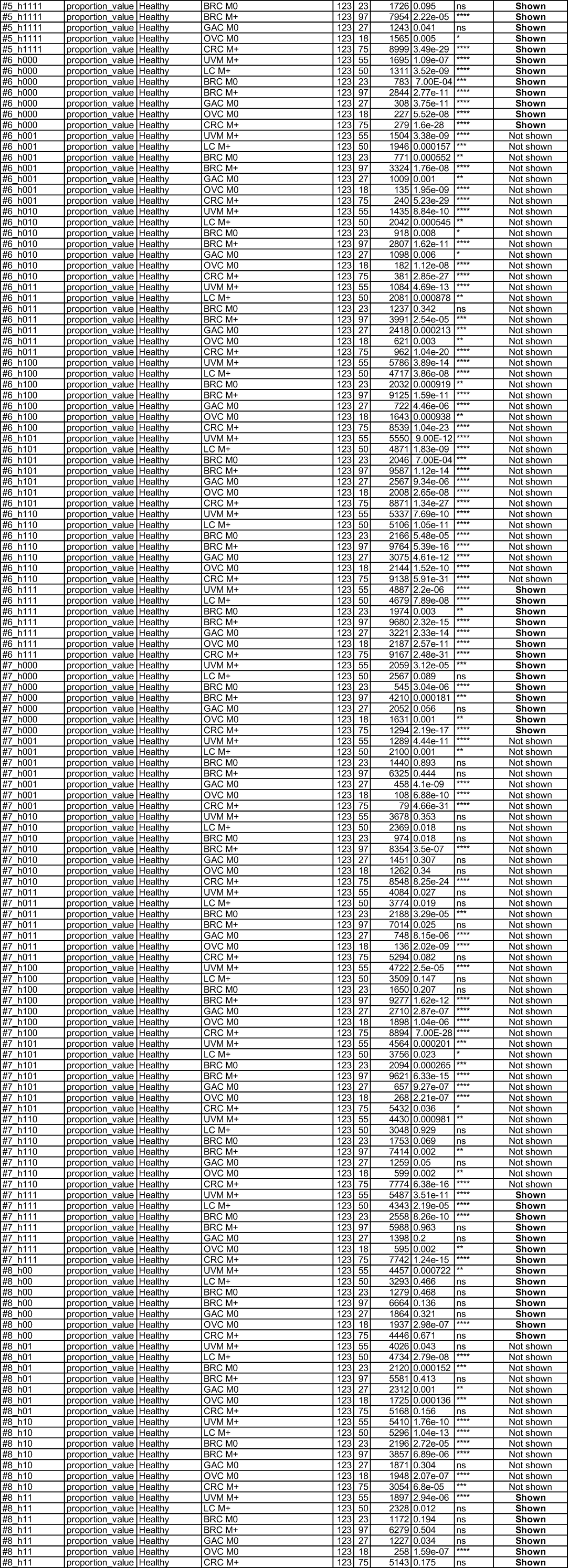
Statistical results of proportion differences between healthy samples and cancer subgroups for the 372 haplotypes using Mann–Whitney U test.

**Table S6.**
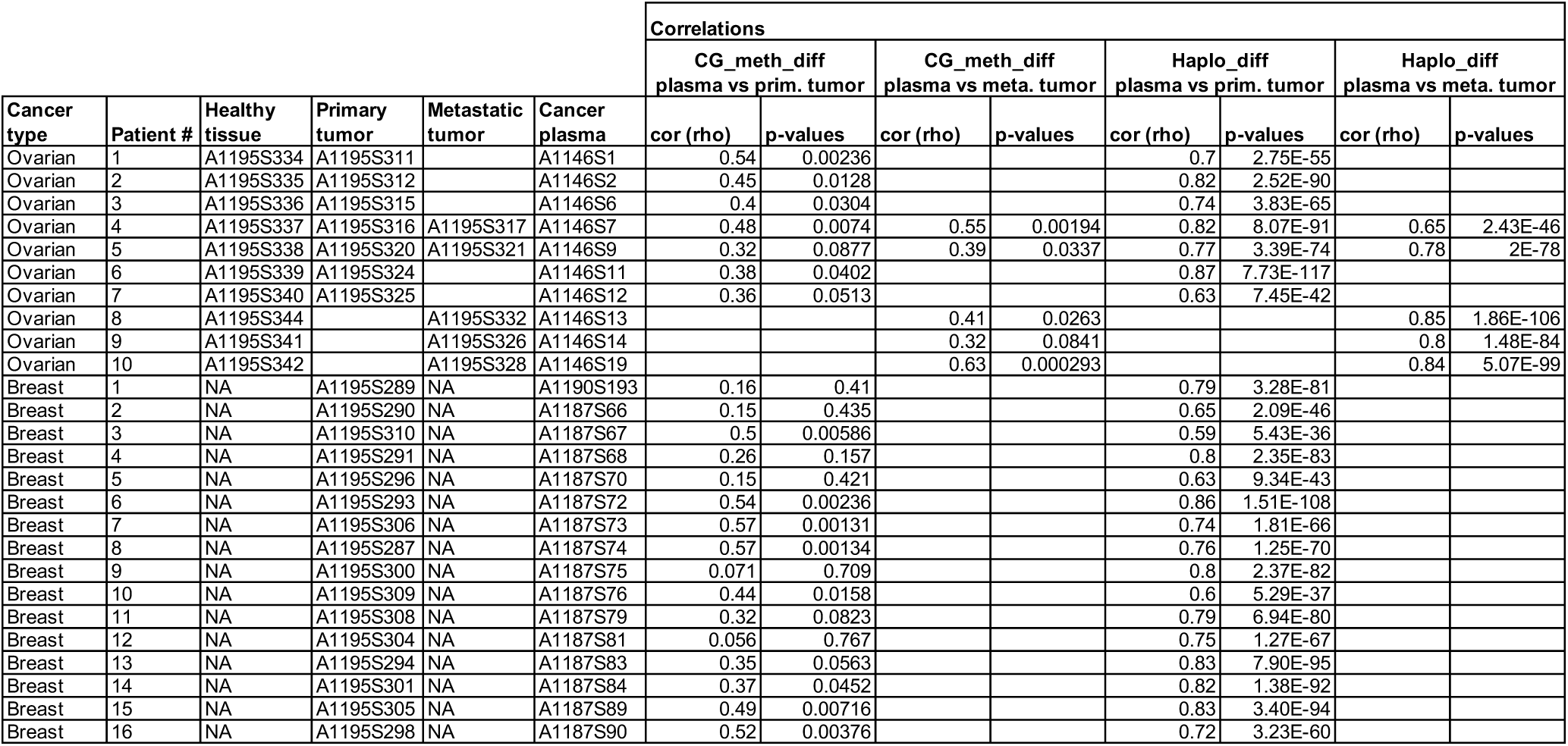
List of paired plasma and tissues samples and their spearman correlation for various features.

| Cancer type | Patient # | Healthy tissue | Primary tumor | Metastatic tumor | Cancer plasma | Correlations |  |  |  |  |  |  |  |
| --- | --- | --- | --- | --- | --- | --- | --- | --- | --- | --- | --- | --- | --- |
|  |  |  |  |  |  | CG_meth_diff plasma vs prim. tumor |  | CG_meth_diff plasma vs meta. tumor |  | Haplo_diff plasma vs prim. tumor |  | Haplo_diff plasma vs meta. tumor |  |
|  |  |  |  |  |  | cor (rho) | p-values | cor (rho) | p-values | cor (rho) | p-values | cor (rho) | p-values |
| Ovarian | 1 | A1195S334 | A1195S311 |  | A1146S1 | 0.54 | 0.00236 |  |  | 0.7 | 2.75E-55 |  |  |
| Ovarian | 2 | A1195S335 | A1195S312 |  | A1146S2 | 0.45 | 0.0128 |  |  | 0.82 | 2.52E-90 |  |  |
| Ovarian | 3 | A1195S336 | A1195S315 |  | A1146S6 | 0.4 | 0.0304 |  |  | 0.74 | 3.83E-65 |  |  |
| Ovarian | 4 | A1195S337 | A1195S316 | A1195S317 | A1146S7 | 0.48 | 0.0074 | 0.55 | 0.00194 | 0.82 | 8.07E-91 | 0.65 | 2.43E-46 |
| Ovarian | 5 | A1195S338 | A1195S320 | A1195S321 | A1146S9 | 0.32 | 0.0877 | 0.39 | 0.0337 | 0.77 | 3.39E-74 | 0.78 | 2E-78 |
| Ovarian | 6 | A1195S339 | A1195S324 |  | A1146S11 | 0.38 | 0.0402 |  |  | 0.87 | 7.73E-117 |  |  |
| Ovarian | 7 | A1195S340 | A1195S325 |  | A1146S12 | 0.36 | 0.0513 |  |  | 0.63 | 7.45E-42 |  |  |
| Ovarian | 8 | A1195S344 |  | A1195S332 | A1146S13 |  |  | 0.41 | 0.0263 |  |  | 0.85 | 1.86E-106 |
| Ovarian | 9 | A1195S341 |  | A1195S326 | A1146S14 |  |  | 0.32 | 0.0841 |  |  | 0.8 | 1.48E-84 |
| Ovarian | 10 | A1195S342 |  | A1195S328 | A1146S19 |  |  | 0.63 | 0.000293 |  |  | 0.84 | 5.07E-99 |
| Breast | 1 | NA | A1195S289 | NA | A1190S193 | 0.16 | 0.41 |  |  | 0.79 | 3.28E-81 |  |  |
| Breast | 2 | NA | A1195S290 | NA | A1187S66 | 0.15 | 0.435 |  |  | 0.65 | 2.09E-46 |  |  |
| Breast | 3 | NA | A1195S310 | NA | A1187S67 | 0.5 | 0.00586 |  |  | 0.59 | 5.43E-36 |  |  |
| Breast | 4 | NA | A1195S291 | NA | A1187S68 | 0.26 | 0.157 |  |  | 0.8 | 2.35E-83 |  |  |
| Breast | 5 | NA | A1195S296 | NA | A1187S70 | 0.15 | 0.421 |  |  | 0.63 | 9.34E-43 |  |  |
| Breast | 6 | NA | A1195S293 | NA | A1187S72 | 0.54 | 0.00236 |  |  | 0.86 | 1.51E-108 |  |  |
| Breast | 7 | NA | A1195S306 | NA | A1187S73 | 0.57 | 0.00131 |  |  | 0.74 | 1.81E-66 |  |  |
| Breast | 8 | NA | A1195S287 | NA | A1187S74 | 0.57 | 0.00134 |  |  | 0.76 | 1.25E-70 |  |  |
| Breast | 9 | NA | A1195S300 | NA | A1187S75 | 0.071 | 0.709 |  |  | 0.8 | 2.37E-82 |  |  |
| Breast | 10 | NA | A1195S309 | NA | A1187S76 | 0.44 | 0.0158 |  |  | 0.6 | 5.29E-37 |  |  |
| Breast | 11 | NA | A1195S308 | NA | A1187S79 | 0.32 | 0.0823 |  |  | 0.79 | 6.94E-80 |  |  |
| Breast | 12 | NA | A1195S304 | NA | A1187S81 | 0.056 | 0.767 |  |  | 0.75 | 1.27E-67 |  |  |
| Breast | 13 | NA | A1195S294 | NA | A1187S83 | 0.35 | 0.0563 |  |  | 0.83 | 7.90E-95 |  |  |
| Breast | 14 | NA | A1195S301 | NA | A1187S84 | 0.37 | 0.0452 |  |  | 0.82 | 1.38E-92 |  |  |
| Breast | 15 | NA | A1195S305 | NA | A1187S89 | 0.49 | 0.00716 |  |  | 0.83 | 3.40E-94 |  |  |
| Breast | 16 | NA | A1195S298 | NA | A1187S90 | 0.52 | 0.00376 |  |  | 0.72 | 3.23E-60 |  |  |

**Table S7.** Breast cancer subtype composition of M0 and M+ subgroups in C1 and C2.

|  | Discovery |  |  |  |  | Validation |  |  |  |  | C1+C2 |  |  |  |  |
| --- | --- | --- | --- | --- | --- | --- | --- | --- | --- | --- | --- | --- | --- | --- | --- |
|  | HR+HER2- | TNBC | HER2+<br>(HR-/HR+) | ND | total | HR+HER2- | TNBC | HER2+<br>(HR-/HR+) | ND | total | HR+HER2- | TNBC | HER2+<br>(HR-/HR+) | ND | total |
| M0 | 11 | 6 | 5 | 1 | 23 | 0 | 56 | 0 | 0 | 56 | 11 | 62 | 5 | 1 | 79 |
|  | 48% | 26% | 22% | 4% | 100% | 0% | 100% | 0% | 0% | 100% | 14% | 78% | 6% | 1% | 100% |
| M+ | 97 | 0 | 0 | 0 | 97 | 28 | 16 | 4 | 0 | 48 | 125 | 16 | 4 | 0 | 145 |
|  | 100% | 0% | 0% | 0% | 100% | 58% | 33% | 8% | 0% | 100% | 86% | 11% | 3% | 0% | 100% |

**Table S8.**
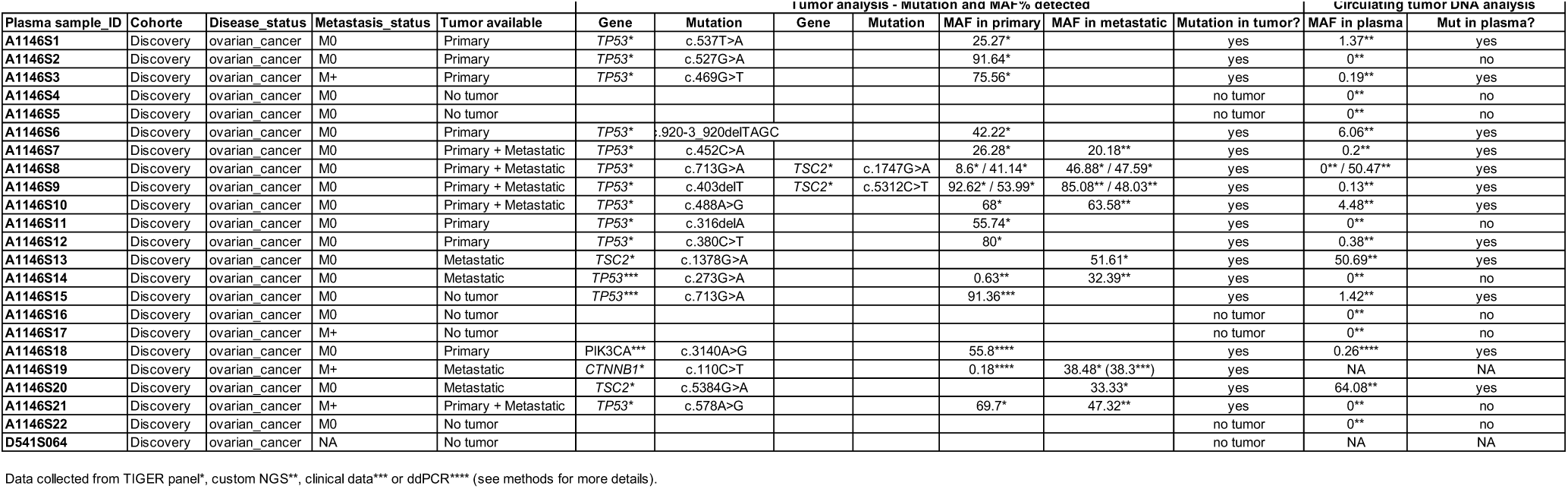
Mutation screening of ovarian cancer samples from the discovery cohort.

**Table S9.** Statistical results of proportion differences between healthy samples from C1 and C2 for 14 haplotypes selected using Mann–Whitney U test.

| Haplotypes | .y. | group1 | group2 | n1 | n2 | statistic | p | p.signif |
| --- | --- | --- | --- | --- | --- | --- | --- | --- |
| #1_h000000 | proportion_value | Healthy_C1 | Healthy_C2 | 123 | 60 | 2165 | 5.85e-06 | **** |
| #1_h111111 | proportion_value | Healthy_C1 | Healthy_C2 | 123 | 60 | 2236 | 1.55e-05 | **** |
| #2_h00000000 | proportion_value | Healthy_C1 | Healthy_C2 | 123 | 60 | 3062 | 6.21e-02 | ns |
| #2_h01111110 | proportion_value | Healthy_C1 | Healthy_C2 | 123 | 60 | 3650 | 9.07e-01 | ns |
| #4_h0000 | proportion_value | Healthy_C1 | Healthy_C2 | 123 | 60 | 2416 | 1.53e-04 | *** |
| #4_h1111 | proportion_value | Healthy_C1 | Healthy_C2 | 123 | 60 | 2132 | 3.66e-06 | **** |
| #5_h0000 | proportion_value | Healthy_C1 | Healthy_C2 | 123 | 60 | 2500 | 4.06e-04 | *** |
| #5_h1111 | proportion_value | Healthy_C1 | Healthy_C2 | 123 | 60 | 2683 | 2.77e-03 | ** |
| #6_h000 | proportion_value | Healthy_C1 | Healthy_C2 | 123 | 60 | 2694 | 3.08e-03 | ** |
| #6_h111 | proportion_value | Healthy_C1 | Healthy_C2 | 123 | 60 | 3622 | 8.41e-01 | ns |
| #7_h000 | proportion_value | Healthy_C1 | Healthy_C2 | 123 | 60 | 3945 | 4.49e-01 | ns |
| #7_h111 | proportion_value | Healthy_C1 | Healthy_C2 | 123 | 60 | 3765 | 8.25e-01 | ns |
| #8_h00 | proportion_value | Healthy_C1 | Healthy_C2 | 123 | 60 | 1651 | 1.36e-09 | **** |
| #8_h11 | proportion_value | Healthy_C1 | Healthy_C2 | 123 | 60 | 5641 | 6.70e-09 | **** |

**Table S10.**
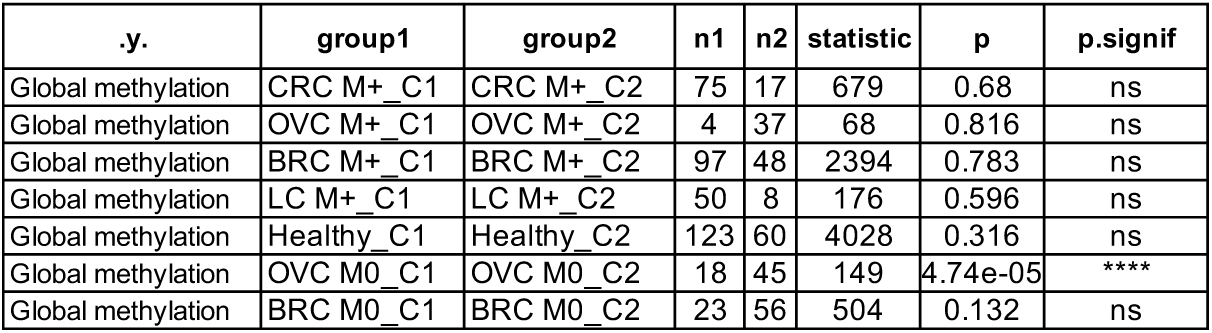
Statistical results of global methylation differences between healthy samples and cancer subgroups from C1 and C2 using Mann–Whitney U test.

**Table S11.**
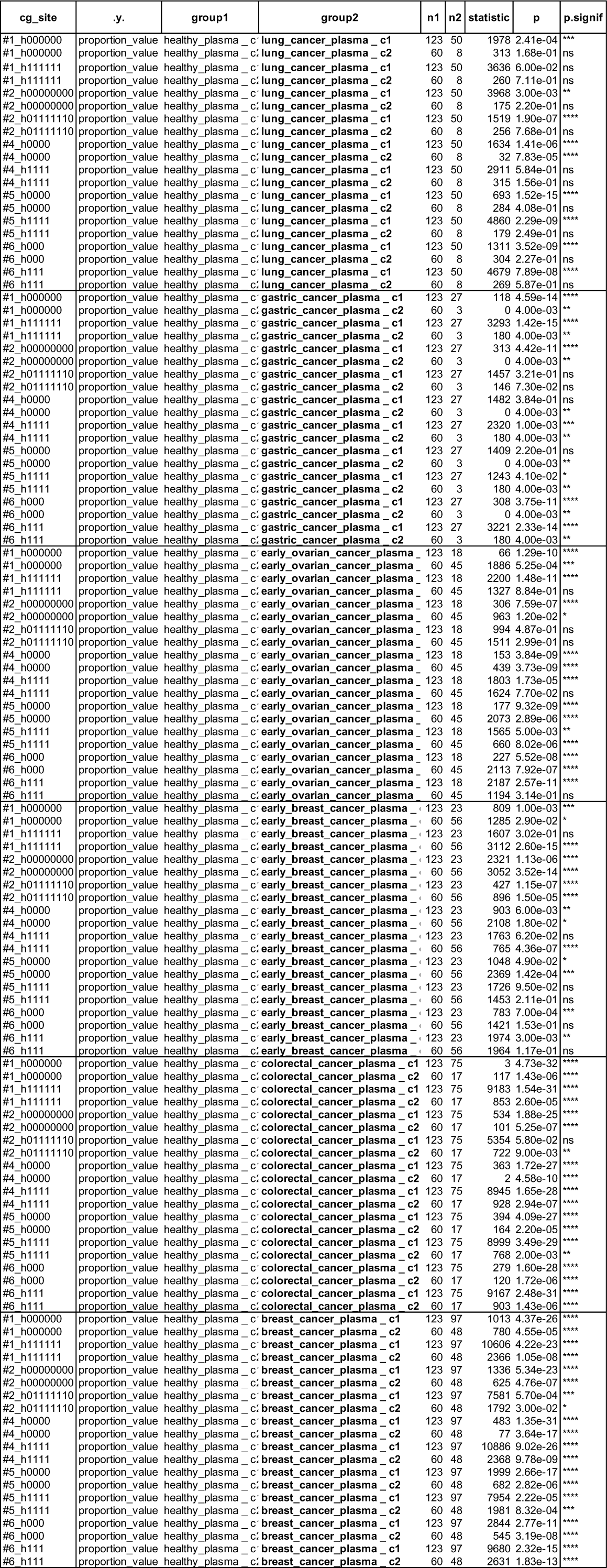
Statistical results of proportion differences between healthy samples and cancer subgroups from C1 and C2 for 14 haplotypes selected using Mann–Whitney U test.

**Table S12.** Statistical results of global methylation differences between original data and age-adjusted data using Mann–Whitney U test.

|  | pvalue_unadjusted | Median difference - 95%CI |  | pvalue_adjusted | Median difference - 95%CI |  |
| --- | --- | --- | --- | --- | --- | --- |
|  |  | lower_unadjusted | upper_unadjusted |  | lower_adjusted | upper_adjusted |
| Healthy vs CRC M+ | 1.0579052012956e-35 | 0.0344045794318254 | 0.0485898838811871 | 1.7897443734997e-33 | 0.0323468589846721 | 0.0470376328481437 |
| Healthy vs BRC M+ | 3.98849696998735e-25 | 0.0147062917799772 | 0.0213450004036148 | 1.15464495255167e-19 | 0.0125436580539427 | 0.0194829734766916 |
| Healthy vs UVM M+ | 7.37313323664488e-06 | 0.00375158485943409 | 0.0103049900893709 | 0.000932832883787769 | 0.00209636623781393 | 0.00896602121839185 |
| Healthy vs NSCLC M+ | 0.862964155819897 | -0.00302065721188611 | 0.00265042706454909 | 0.0523632070508789 | -0.00579373542715967 | 2.79914815230682e-05 |
| Healthy vs GAC M+ | 0.00305315125460983 | 0.0850492895889727 | 0.12512770303051 | 0.00305315125460983 | 0.083141140524617 | 0.122386608575801 |
| Healthy vs OVC M+ | 0.000104790124555637 | 0.00453204067341506 | 0.0142415490819948 | 0.00817381448084698 | 0.00167254696788085 | 0.0120420537412205 |
| Healthy vs OVC M0 | 0.947624990127436 | -0.00372090876019228 | 0.00342785215393217 | 0.105305755371932 | -0.00674368110890361 | 0.000623249146482699 |
| Healthy vs GAC M0 | 1.76054842755586e-08 | 0.00642136151723168 | 0.011838295258056 | 5.27849884994781e-05 | 0.00333037306841255 | 0.00930641545722199 |
| Healthy vs BRC M0 | 0.0176227712287838 | -0.00477528244072236 | -0.000433028832155155 | 0.0090144461485175 | -0.00546047896132789 | -0.000823016158638357 |

**Table S13.** Statistical results of global methylation differences between cancer stages using Mann–Whitney U test.

| .y. | group1 | group2 | n1 | n2 | statistic | p | p.signif |
| --- | --- | --- | --- | --- | --- | --- | --- |
| Global methylation | Cancer_early | Cancer_advanced | 69 | 84 | 3166 | 0.327 | ns |
| Global methylation | Cancer_early | Cancer_meta | 69 | 394 | 21392 | 2.82e-14 | **** |
| Global methylation | Cancer_advanced | Cancer_meta | 84 | 394 | 24180 | 3.14e-11 | **** |
| Global methylation | BRC M0 C1+C2 | BRC M+ C1+C2 | 145 | 79 | 1646.5 | 1.3e-18 | **** |
| Global methylation | GAC M0 C1 | GAC M+ C2 | 27 | 3 | 81.0 | 5.0e-03 | ** |
| Global methylation | OVC M0 C1+C2 | OVC M+ C1+C2 | 63 | 42 | 1742.5 | 6.0e-03 | ** |

**Table S14.**
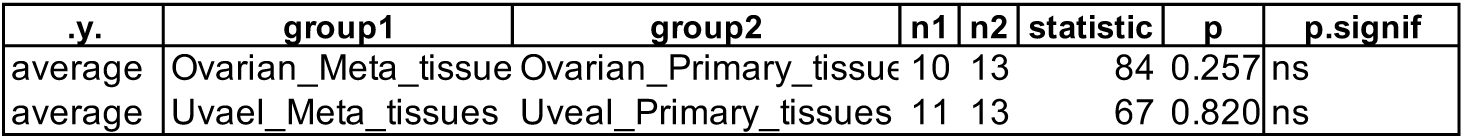
Statistical results of global methylation differences between primary and metastatic tissues using Mann–Whitney U test.

| .y. | group1 | group2 | n1 | n2 | statistic | p | p.signif |
| --- | --- | --- | --- | --- | --- | --- | --- |
| average | Ovarian_Meta_tissue | Ovarian_Primary_tissue | 10 | 13 | 84 | 0.257 | ns |
| average | Uveal_Meta_tissues | Uveal_Primary_tissues | 11 | 13 | 67 | 0.820 | ns |

**Table S15.** Statistical results of Genome-wide z-score differences between healthy samples and cancer subgroups using Mann–Whitney U test.

| .y. | group1 | group2 | n1 | n2 | statistic | p | p.signif |
| --- | --- | --- | --- | --- | --- | --- | --- |
| Gzscore | Healthy | BRC M+ | 120 | 145 | 3213 | 1.01E-18 | **** |
| Gzscore | Healthy | BRC M0 | 120 | 79 | 1407 | 5.12E-17 | **** |
| Gzscore | Healthy | CRC M+ | 120 | 92 | 1644 | 2.05E-18 | **** |
| Gzscore | Healthy | GAC M+ | 120 | 3 | 0 | 0.003 | ** |
| Gzscore | Healthy | GAC M0 | 120 | 27 | 729 | 8.40E-06 | **** |
| Gzscore | Healthy | LC M+ | 120 | 58 | 3575 | 0.769 | ns |
| Gzscore | Healthy | OVC M+ | 120 | 41 | 728 | 1.84E-11 | **** |
| Gzscore | Healthy | OVC M0 | 120 | 63 | 1356 | 1.09E-12 | **** |
| Gzscore | Healthy | OVC NA | 120 | 1 | 5 | 0.119 | ns |
| Gzscore | Healthy | UVM M+ | 120 | 55 | 2872 | 0.169 | ns |

**Table S16.**
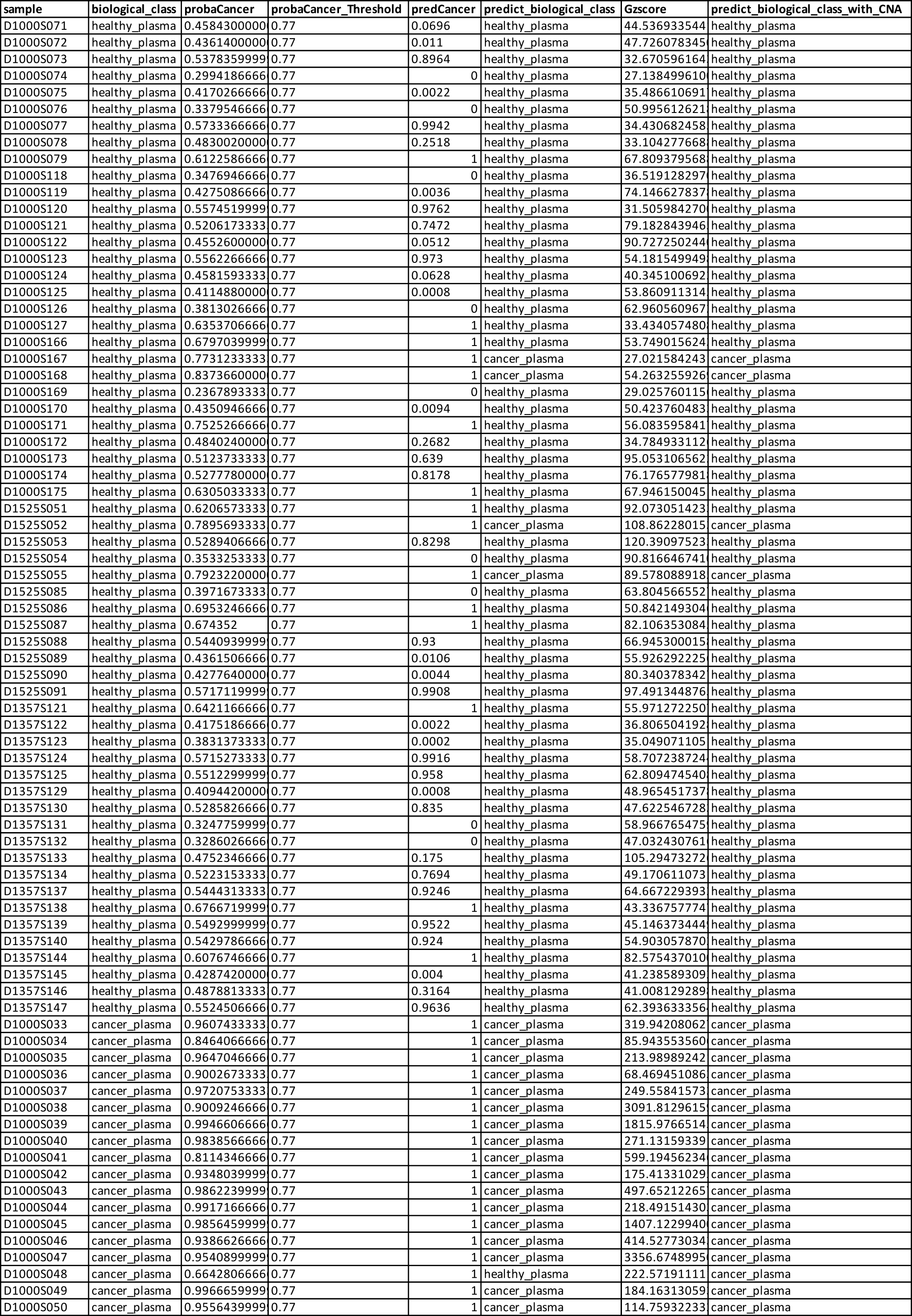
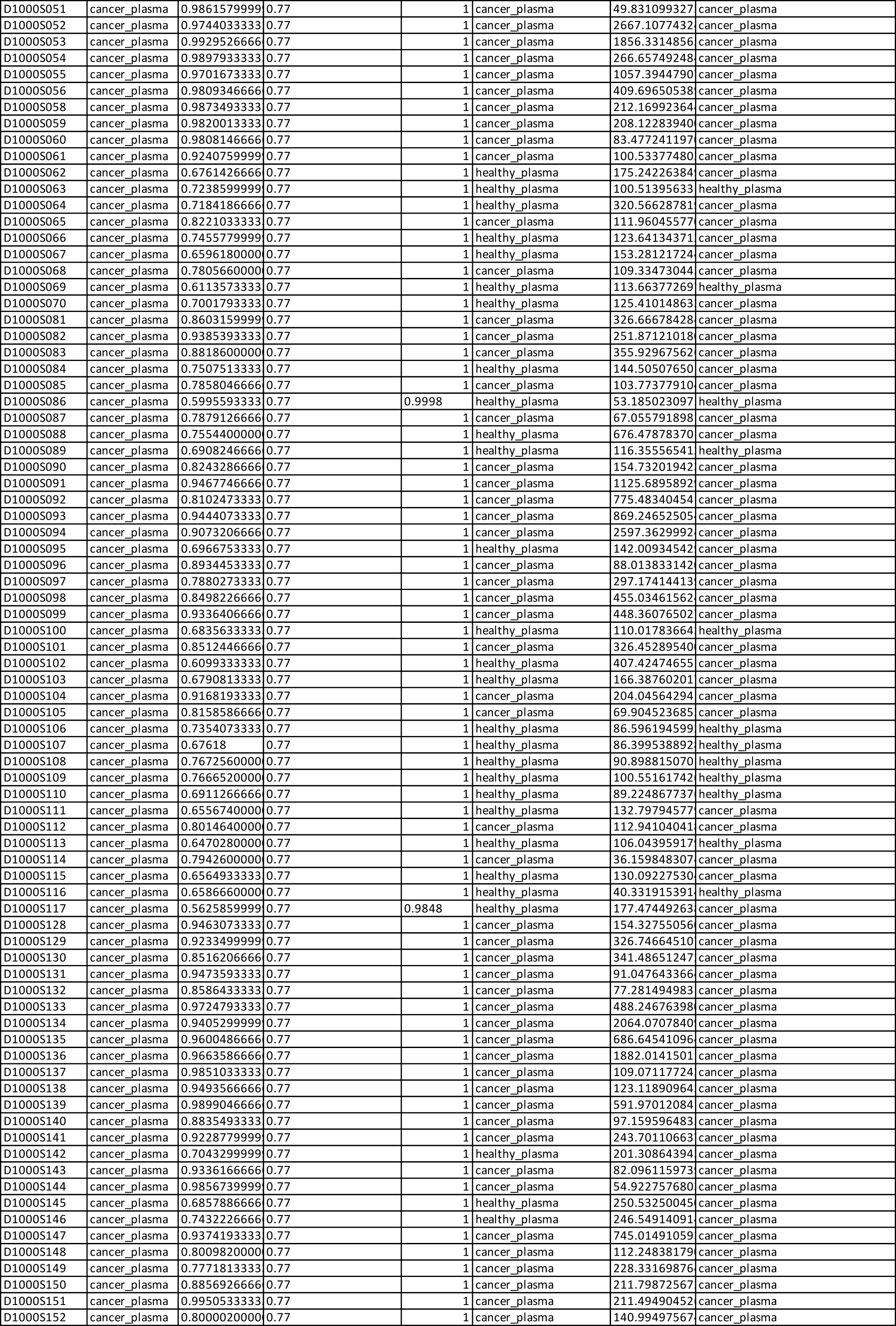
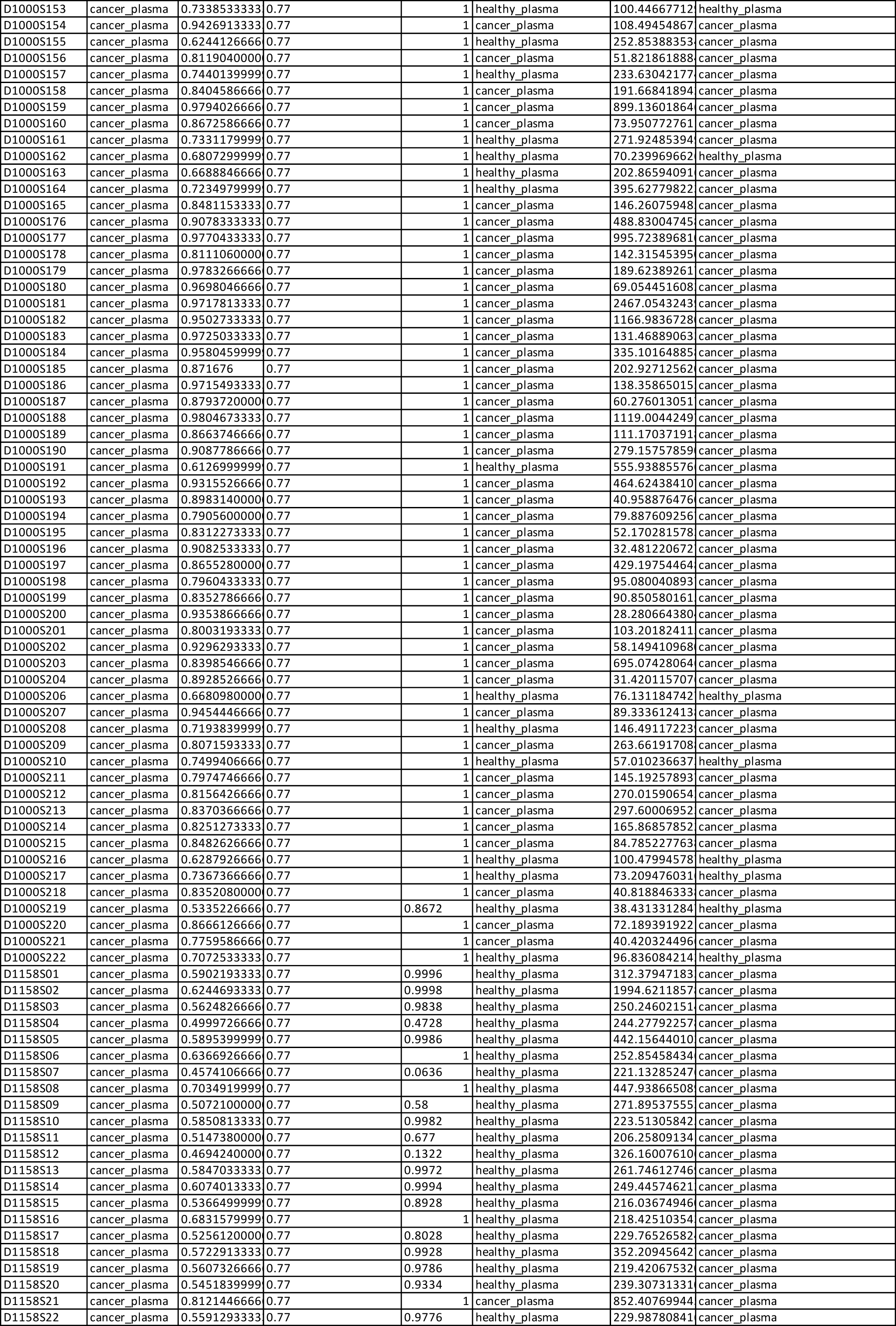
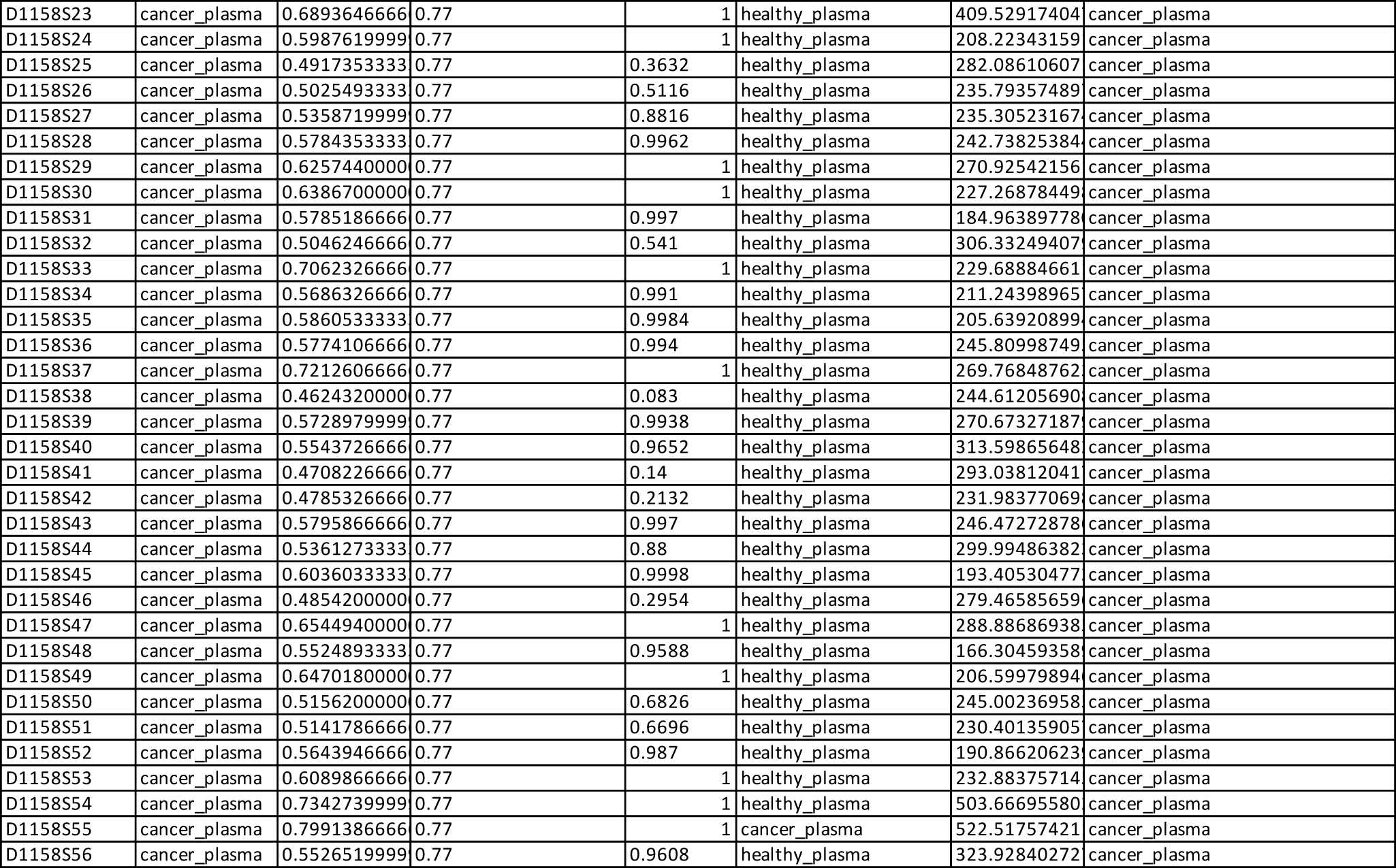
Cancer prediction and sample labelling with the 2-step classification integrating CNA analysis - healthy vs all cancers model.

**Table S17.**
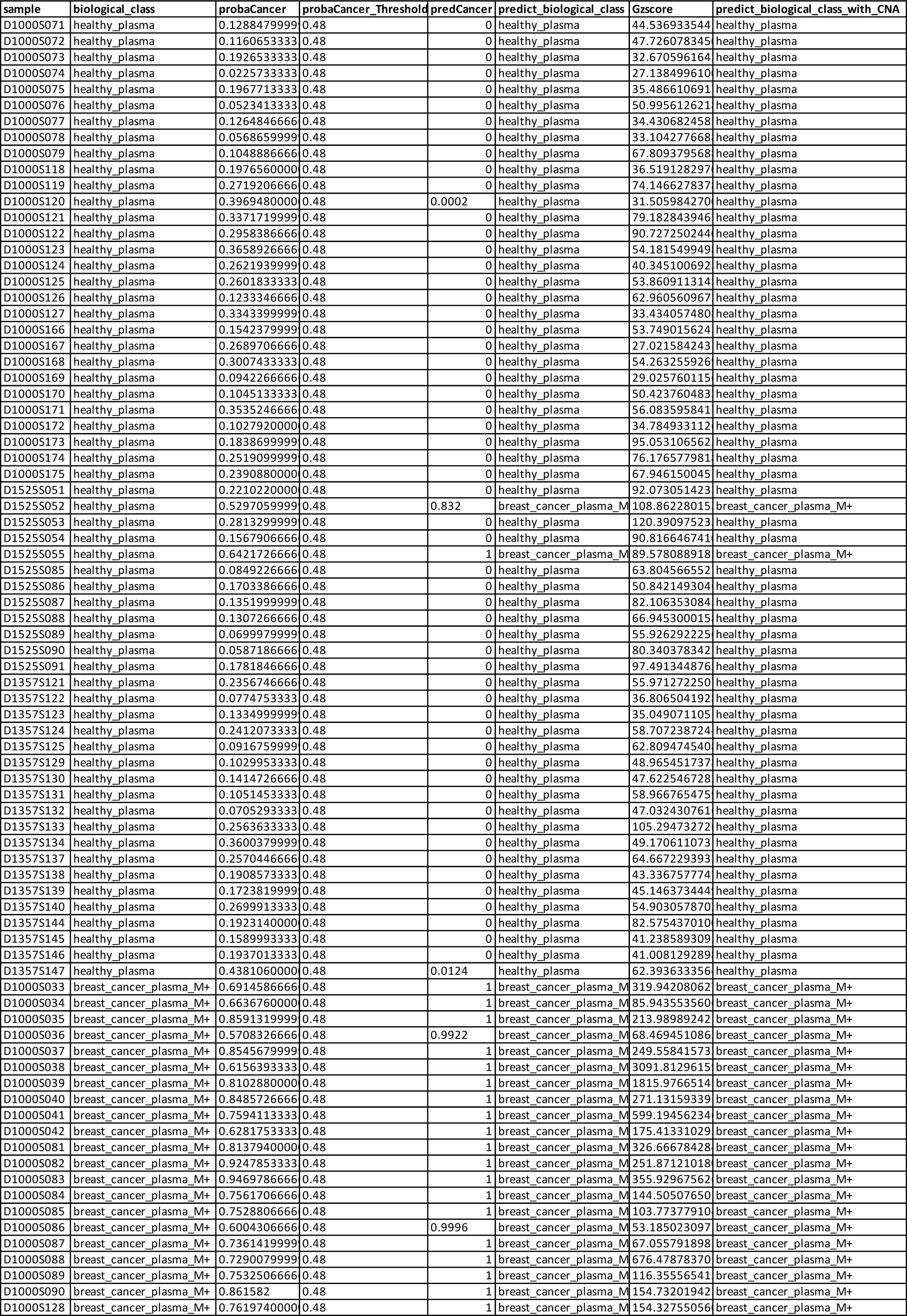
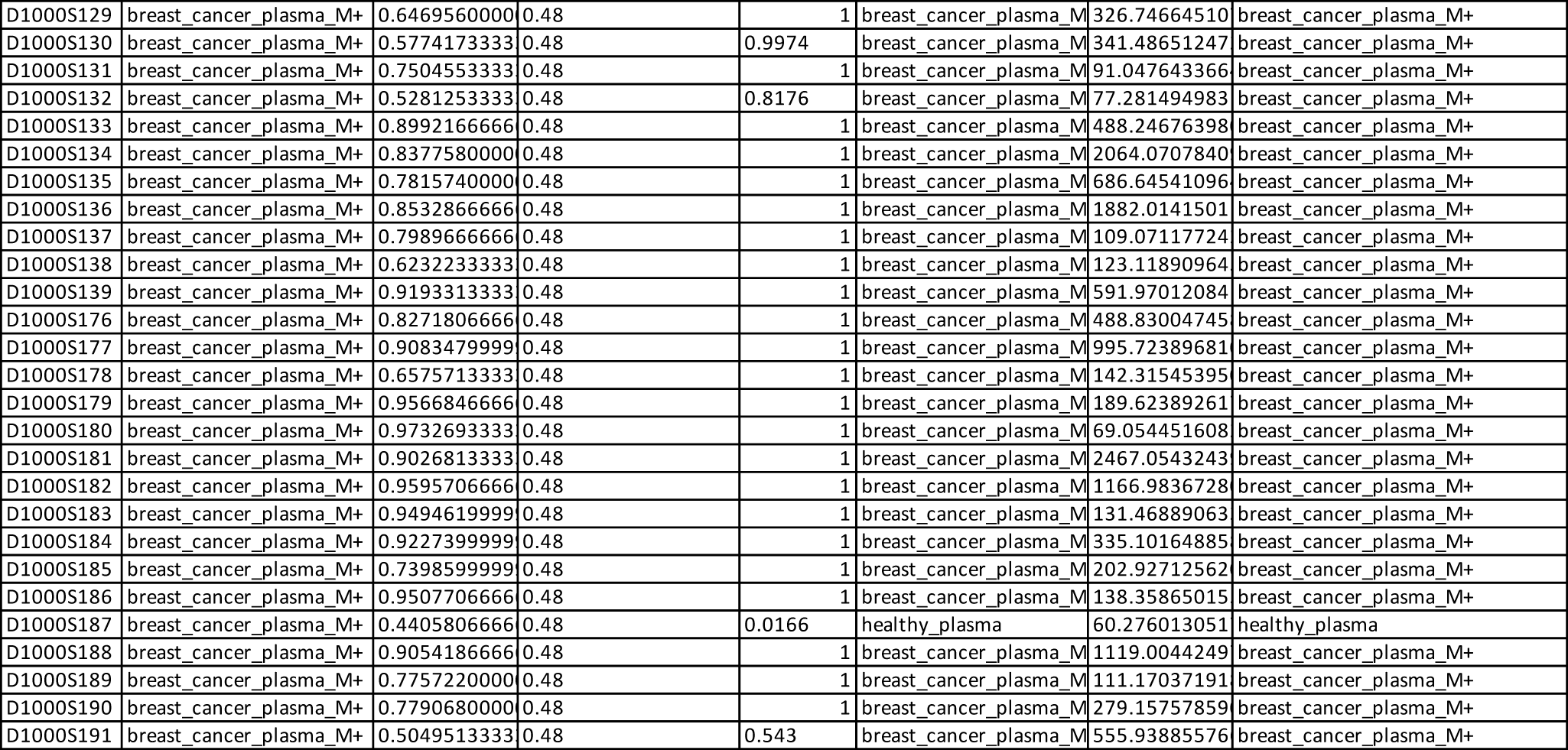
Cancer prediction and sample labelling with the 2-step classification integrating CNA analysis - healthy vs BRC M+ model.

**Table S18.**
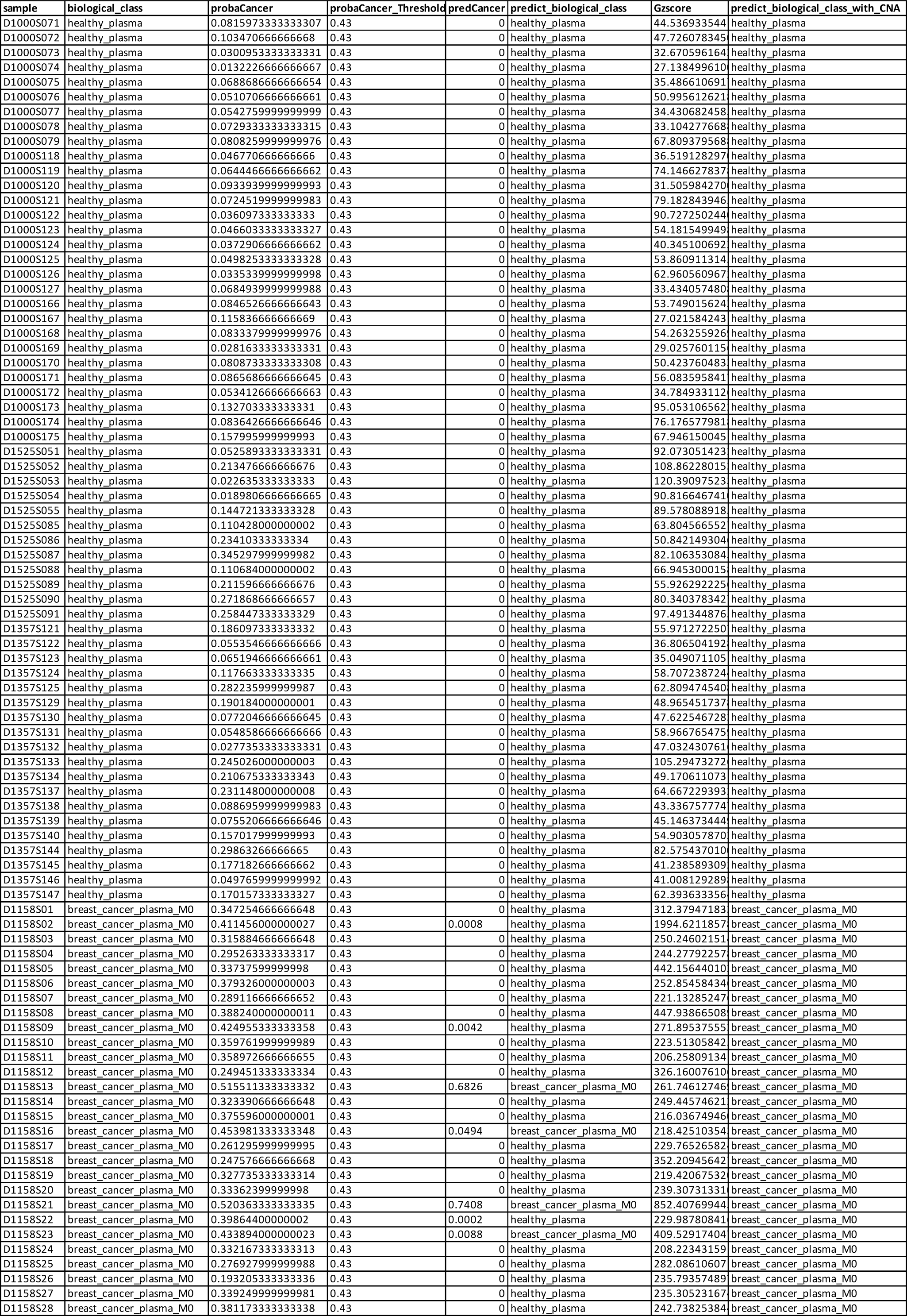
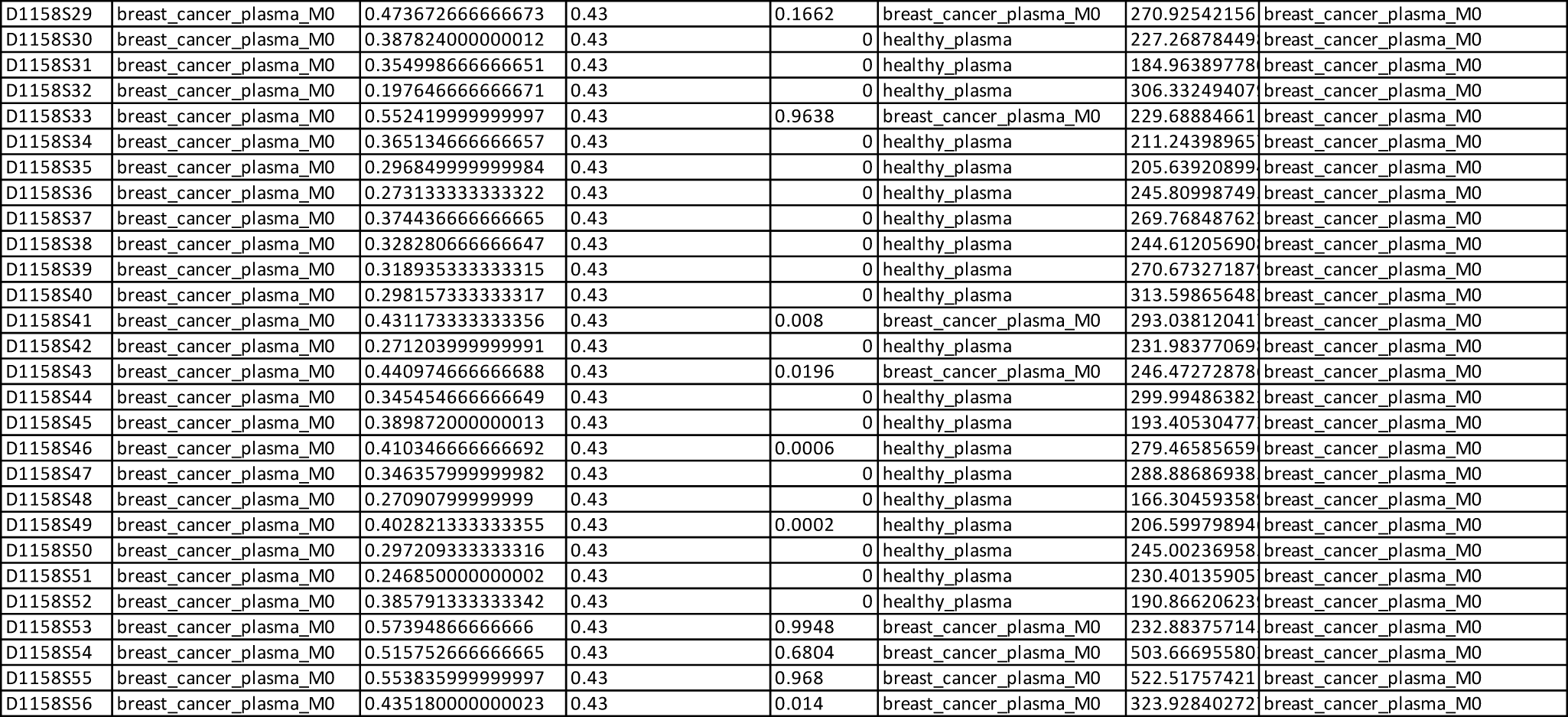
Cancer prediction and sample labelling with the 2-step classification integrating CNA analysis - healthy vs BRC M0 model.

**Table S19.** Cancer prediction and sample labelling with the 2-step classification integrating CNA analysis - healthy vs CRC M+ model.

| sample | biological_class | probaCancer | probaCancer_Threshold | predCancer | predict_biological_class | Gzscore | predict_biological_class_with_CNA |
| --- | --- | --- | --- | --- | --- | --- | --- |
| D1000S071 | healthy_plasma | 0.0159919999 | 0.35 | 0 | healthy_plasma | 44.536933544 | healthy_plasma |
| D1000S072 | healthy_plasma | 0.0210593333 | 0.35 | 0 | healthy_plasma | 47.726078345 | healthy_plasma |
| D1000S073 | healthy_plasma | 0.0204553333 | 0.35 | 0 | healthy_plasma | 32.670596164 | healthy_plasma |
| D1000S074 | healthy_plasma | 0.0004573333 | 0.35 | 0 | healthy_plasma | 27.138499610 | healthy_plasma |
| D1000S075 | healthy_plasma | 0.0516013333 | 0.35 | 0 | healthy_plasma | 35.486610691 | healthy_plasma |
| D1000S076 | healthy_plasma | 0.0042493333 | 0.35 | 0 | healthy_plasma | 50.995612621 | healthy_plasma |
| D1000S077 | healthy_plasma | 0.0520893333 | 0.35 | 0 | healthy_plasma | 34.430682458 | healthy_plasma |
| D1000S078 | healthy_plasma | 0.0060046666 | 0.35 | 0 | healthy_plasma | 33.104277668 | healthy_plasma |
| D1000S079 | healthy_plasma | 0.0231986666 | 0.35 | 0 | healthy_plasma | 67.809379568 | healthy_plasma |
| D1000S118 | healthy_plasma | 0.0200626666 | 0.35 | 0 | healthy_plasma | 36.519128297 | healthy_plasma |
| D1000S119 | healthy_plasma | 0.0238999999 | 0.35 | 0 | healthy_plasma | 74.146627837 | healthy_plasma |
| D1000S120 | healthy_plasma | 0.0774893333 | 0.35 | 0 | healthy_plasma | 31.505984270 | healthy_plasma |
| D1000S121 | healthy_plasma | 0.0641233333 | 0.35 | 0 | healthy_plasma | 79.182843946 | healthy_plasma |
| D1000S122 | healthy_plasma | 0.0233953333 | 0.35 | 0 | healthy_plasma | 90.727250244 | healthy_plasma |
| D1000S123 | healthy_plasma | 0.0240826666 | 0.35 | 0 | healthy_plasma | 54.181549949 | healthy_plasma |
| D1000S124 | healthy_plasma | 0.0253779999 | 0.35 | 0 | healthy_plasma | 40.345100692 | healthy_plasma |
| D1000S125 | healthy_plasma | 0.0215979999 | 0.35 | 0 | healthy_plasma | 53.860911314 | healthy_plasma |
| D1000S126 | healthy_plasma | 0.0116853333 | 0.35 | 0 | healthy_plasma | 62.960560967 | healthy_plasma |
| D1000S127 | healthy_plasma | 0.0191153333 | 0.35 | 0 | healthy_plasma | 33.434057480 | healthy_plasma |
| D1000S166 | healthy_plasma | 0.0274173333 | 0.35 | 0 | healthy_plasma | 53.749015624 | healthy_plasma |
| D1000S167 | healthy_plasma | 0.0278219999 | 0.35 | 0 | healthy_plasma | 27.021584243 | healthy_plasma |
| D1000S168 | healthy_plasma | 0.0621879999 | 0.35 | 0 | healthy_plasma | 54.263255926 | healthy_plasma |
| D1000S169 | healthy_plasma | 0.0052093333 | 0.35 | 0 | healthy_plasma | 29.025760115 | healthy_plasma |
| D1000S170 | healthy_plasma | 0.0120046666 | 0.35 | 0 | healthy_plasma | 50.423760483 | healthy_plasma |
| D1000S171 | healthy_plasma | 0.0510666666 | 0.35 | 0 | healthy_plasma | 56.083595841 | healthy_plasma |
| D1000S172 | healthy_plasma | 0.0316419999 | 0.35 | 0 | healthy_plasma | 34.784933112 | healthy_plasma |
| D1000S173 | healthy_plasma | 0.0181793333 | 0.35 | 0 | healthy_plasma | 95.053106562 | healthy_plasma |
| D1000S174 | healthy_plasma | 0.0361066666 | 0.35 | 0 | healthy_plasma | 76.176577981 | healthy_plasma |
| D1000S175 | healthy_plasma | 0.0172146666 | 0.35 | 0 | healthy_plasma | 67.946150045 | healthy_plasma |
| D1525S051 | healthy_plasma | 0.0368199999 | 0.35 | 0 | healthy_plasma | 92.073051423 | healthy_plasma |
| D1525S052 | healthy_plasma | 0.1839846666 | 0.35 | 0 | healthy_plasma | 108.86228015 | healthy_plasma |
| D1525S053 | healthy_plasma | 0.0164106666 | 0.35 | 0 | healthy_plasma | 120.39097523 | healthy_plasma |
| D1525S054 | healthy_plasma | 0.0097733333 | 0.35 | 0 | healthy_plasma | 90.816646741 | healthy_plasma |
| D1525S055 | healthy_plasma | 0.1737353333 | 0.35 | 0 | healthy_plasma | 89.578088918 | healthy_plasma |
| D1525S085 | healthy_plasma | 0.0356133333 | 0.35 | 0 | healthy_plasma | 63.804566552 | healthy_plasma |
| D1525S086 | healthy_plasma | 0.1028993333 | 0.35 | 0 | healthy_plasma | 50.842149304 | healthy_plasma |
| D1525S087 | healthy_plasma | 0.0695713333 | 0.35 | 0 | healthy_plasma | 82.106353084 | healthy_plasma |
| D1525S088 | healthy_plasma | 0.0337959999 | 0.35 | 0 | healthy_plasma | 66.945300015 | healthy_plasma |
| D1525S089 | healthy_plasma | 0.0154053333 | 0.35 | 0 | healthy_plasma | 55.926292225 | healthy_plasma |
| D1525S090 | healthy_plasma | 0.0161219999 | 0.35 | 0 | healthy_plasma | 80.340378342 | healthy_plasma |
| D1525S091 | healthy_plasma | 0.0739413333 | 0.35 | 0 | healthy_plasma | 97.491344876 | healthy_plasma |
| D1357S121 | healthy_plasma | 0.0663346666 | 0.35 | 0 | healthy_plasma | 55.971272250 | healthy_plasma |
| D1357S122 | healthy_plasma | 0.014398 | 0.35 | 0 | healthy_plasma | 36.806504192 | healthy_plasma |
| D1357S123 | healthy_plasma | 0.0303306666 | 0.35 | 0 | healthy_plasma | 35.049071105 | healthy_plasma |
| D1357S124 | healthy_plasma | 0.0521806666 | 0.35 | 0 | healthy_plasma | 58.707238724 | healthy_plasma |
| D1357S125 | healthy_plasma | 0.0150226666 | 0.35 | 0 | healthy_plasma | 62.809474540 | healthy_plasma |
| D1357S129 | healthy_plasma | 0.0262039999 | 0.35 | 0 | healthy_plasma | 48.965451737 | healthy_plasma |
| D1357S130 | healthy_plasma | 0.0280593333 | 0.35 | 0 | healthy_plasma | 47.622546728 | healthy_plasma |
| D1357S131 | healthy_plasma | 0.0145993333 | 0.35 | 0 | healthy_plasma | 58.966765475 | healthy_plasma |
| D1357S132 | healthy_plasma | 0.0066780000 | 0.35 | 0 | healthy_plasma | 47.032430761 | healthy_plasma |
| D1357S133 | healthy_plasma | 0.0754693333 | 0.35 | 0 | healthy_plasma | 105.29473272 | healthy_plasma |
| D1357S134 | healthy_plasma | 0.1558926666 | 0.35 | 0 | healthy_plasma | 49.170611073 | healthy_plasma |
| D1357S137 | healthy_plasma | 0.100934 | 0.35 | 0 | healthy_plasma | 64.667229393 | healthy_plasma |
| D1357S138 | healthy_plasma | 0.0382099999 | 0.35 | 0 | healthy_plasma | 43.336757774 | healthy_plasma |
| D1357S139 | healthy_plasma | 0.0169746666 | 0.35 | 0 | healthy_plasma | 45.146373444 | healthy_plasma |
| D1357S140 | healthy_plasma | 0.0638093333 | 0.35 | 0 | healthy_plasma | 54.903057870 | healthy_plasma |
| D1357S144 | healthy_plasma | 0.0719786666 | 0.35 | 0 | healthy_plasma | 82.575437010 | healthy_plasma |
| D1357S145 | healthy_plasma | 0.0373913333 | 0.35 | 0 | healthy_plasma | 41.238589309 | healthy_plasma |
| D1357S146 | healthy_plasma | 0.0433499999 | 0.35 | 0 | healthy_plasma | 41.008129289 | healthy_plasma |
| D1357S147 | healthy_plasma | 0.0845999999 | 0.35 | 0 | healthy_plasma | 62.393633356 | healthy_plasma |
| D1000S043 | colorectal_cancer_plasma_M+ | 0.9470673333 | 0.35 | 1 | colorectal_cancer_plasma | 497.65212265 | colorectal_cancer_plasma_M+ |
| D1000S044 | colorectal_cancer_plasma_M+ | 0.8868060000 | 0.35 | 1 | colorectal_cancer_plasma | 218.49151430 | colorectal_cancer_plasma_M+ |
| D1000S045 | colorectal_cancer_plasma_M+ | 0.9825739999 | 0.35 | 1 | colorectal_cancer_plasma | 1407.1229940 | colorectal_cancer_plasma_M+ |
| D1000S046 | colorectal_cancer_plasma_M+ | 0.5529939999 | 0.35 | 0.9612 | colorectal_cancer_plasma | 414.52773034 | colorectal_cancer_plasma_M+ |
| D1000S047 | colorectal_cancer_plasma_M+ | 0.9056359999 | 0.35 | 1 | colorectal_cancer_plasma | 3356.6748995 | colorectal_cancer_plasma_M+ |
| D1000S048 | colorectal_cancer_plasma_M+ | 0.1556066666 | 0.35 | 0 | healthy_plasma | 222.57191111 | colorectal_cancer_plasma_M+ |
| D1000S049 | colorectal_cancer_plasma_M+ | 0.9127806666 | 0.35 | 1 | colorectal_cancer_plasma | 184.16313059 | colorectal_cancer_plasma_M+ |
| D1000S050 | colorectal_cancer_plasma_M+ | 0.6070533333 | 0.35 | 1 | colorectal_cancer_plasma | 114.75932233 | colorectal_cancer_plasma_M+ |
| D1000S051 | colorectal_cancer_plasma_M+ | 0.67357 | 0.35 | 1 | colorectal_cancer_plasma | 49.831099327 | colorectal_cancer_plasma_M+ |
| D1000S091 | colorectal_cancer_plasma_M+ | 0.9017233333 | 0.35 | 1 | colorectal_cancer_plasma | 1125.6895892 | colorectal_cancer_plasma_M+ |
| D1000S092 | colorectal_cancer_plasma_M+ | 0.7177413333 | 0.35 | 1 | colorectal_cancer_plasma | 775.48340454 | colorectal_cancer_plasma_M+ |
| D1000S093 | colorectal_cancer_plasma_M+ | 0.9070199999 | 0.35 | 1 | colorectal_cancer_plasma | 869.24652505 | colorectal_cancer_plasma_M+ |
| D1000S094 | colorectal_cancer_plasma_M+ | 0.9544613333 | 0.35 | 1 | colorectal_cancer_plasma | 2597.3629992 | colorectal_cancer_plasma_M+ |
| D1000S095 | colorectal_cancer_plasma_M+ | 0.2409633333 | 0.35 | 0 | healthy_plasma | 142.00934542 | colorectal_cancer_plasma_M+ |
| D1000S096 | colorectal_cancer_plasma_M+ | 0.6103799999 | 0.35 | 1 | colorectal_cancer_plasma | 88.013833142 | colorectal_cancer_plasma_M+ |
| D1000S097 | colorectal_cancer_plasma_M+ | 0.5130086666 | 0.35 | 0.6484 | colorectal_cancer_plasma | 297.17414413 | colorectal_cancer_plasma_M+ |
| D1000S098 | colorectal_cancer_plasma_M+ | 0.7864646666 | 0.35 | 1 | colorectal_cancer_plasma | 455.03461562 | colorectal_cancer_plasma_M+ |

**Table S20.**
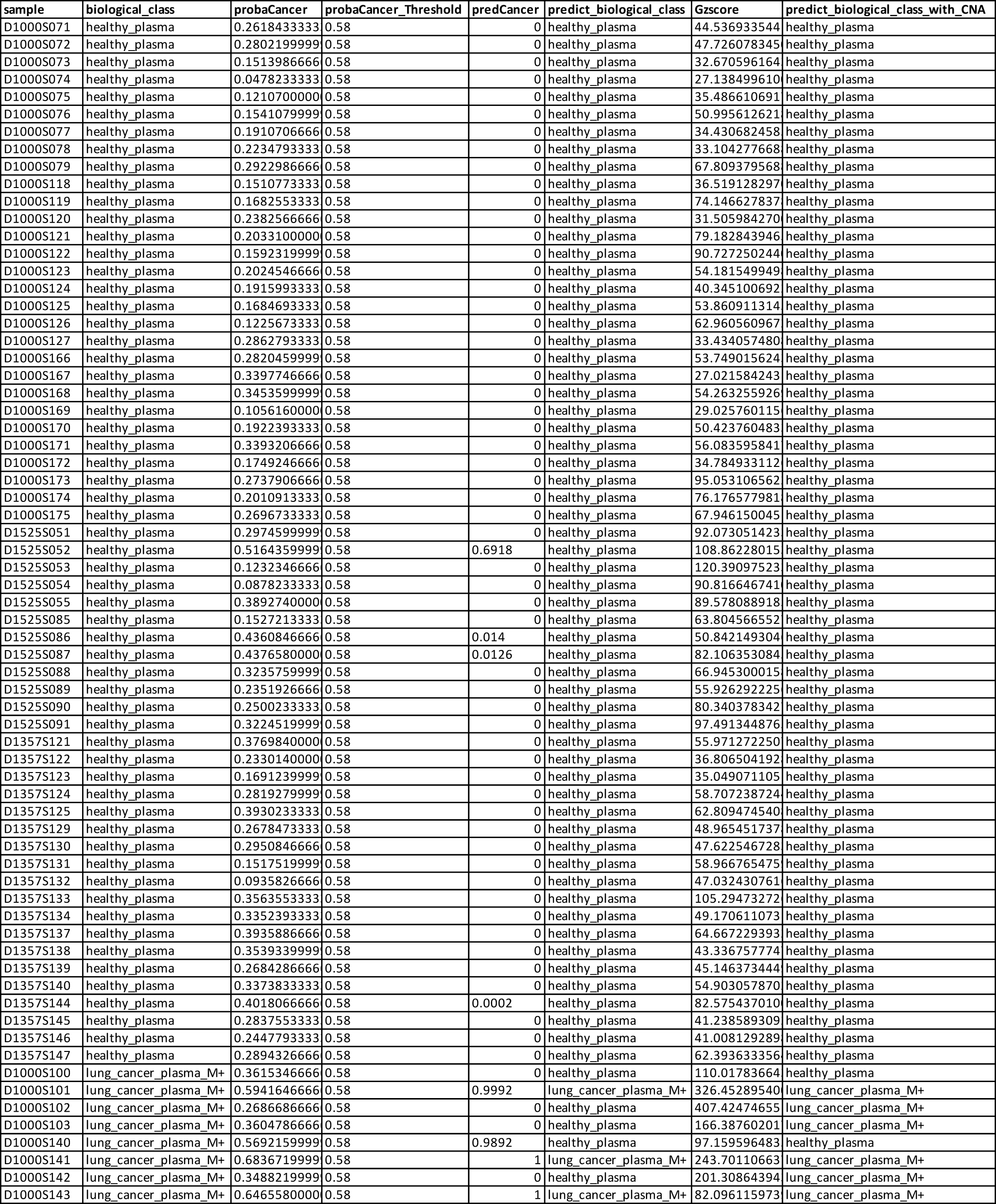
Cancer prediction and sample labelling with the 2-step classification integrating CNA analysis - healthy vs LC M+ model.

| sample | biological_class | probaCancer | probaCancer_Threshold | predCancer | predict_biological_class | Gzscore | predict_biological_class_with_CNA |
| --- | --- | --- | --- | --- | --- | --- | --- |
| D1000S071 | healthy_plasma | 0.2618433333 | 0.58 | 0 | healthy_plasma | 44.536933544 | healthy_plasma |
| D1000S072 | healthy_plasma | 0.2802199999 | 0.58 | 0 | healthy_plasma | 47.726078345 | healthy_plasma |
| D1000S073 | healthy_plasma | 0.1513986666 | 0.58 | 0 | healthy_plasma | 32.670596164 | healthy_plasma |
| D1000S074 | healthy_plasma | 0.0478233333 | 0.58 | 0 | healthy_plasma | 27.138499610 | healthy_plasma |
| D1000S075 | healthy_plasma | 0.1210700000 | 0.58 | 0 | healthy_plasma | 35.486610691 | healthy_plasma |
| D1000S076 | healthy_plasma | 0.1541079999 | 0.58 | 0 | healthy_plasma | 50.995612621 | healthy_plasma |
| D1000S077 | healthy_plasma | 0.1910706666 | 0.58 | 0 | healthy_plasma | 34.430682458 | healthy_plasma |
| D1000S078 | healthy_plasma | 0.2234793333 | 0.58 | 0 | healthy_plasma | 33.104277668 | healthy_plasma |
| D1000S079 | healthy_plasma | 0.2922986666 | 0.58 | 0 | healthy_plasma | 67.809379568 | healthy_plasma |
| D1000S118 | healthy_plasma | 0.1510773333 | 0.58 | 0 | healthy_plasma | 36.519128297 | healthy_plasma |
| D1000S119 | healthy_plasma | 0.1682553333 | 0.58 | 0 | healthy_plasma | 74.146627837 | healthy_plasma |
| D1000S120 | healthy_plasma | 0.2382566666 | 0.58 | 0 | healthy_plasma | 31.505984270 | healthy_plasma |
| D1000S121 | healthy_plasma | 0.2033100000 | 0.58 | 0 | healthy_plasma | 79.182843946 | healthy_plasma |
| D1000S122 | healthy_plasma | 0.1592319999 | 0.58 | 0 | healthy_plasma | 90.727250244 | healthy_plasma |
| D1000S123 | healthy_plasma | 0.2024546666 | 0.58 | 0 | healthy_plasma | 54.181549949 | healthy_plasma |
| D1000S124 | healthy_plasma | 0.1915993333 | 0.58 | 0 | healthy_plasma | 40.345100692 | healthy_plasma |
| D1000S125 | healthy_plasma | 0.1684693333 | 0.58 | 0 | healthy_plasma | 53.860911314 | healthy_plasma |
| D1000S126 | healthy_plasma | 0.1225673333 | 0.58 | 0 | healthy_plasma | 62.960560967 | healthy_plasma |
| D1000S127 | healthy_plasma | 0.2862793333 | 0.58 | 0 | healthy_plasma | 33.434057480 | healthy_plasma |
| D1000S166 | healthy_plasma | 0.2820459999 | 0.58 | 0 | healthy_plasma | 53.749015624 | healthy_plasma |
| D1000S167 | healthy_plasma | 0.3397746666 | 0.58 | 0 | healthy_plasma | 27.021584243 | healthy_plasma |
| D1000S168 | healthy_plasma | 0.3453599999 | 0.58 | 0 | healthy_plasma | 54.263255926 | healthy_plasma |
| D1000S169 | healthy_plasma | 0.1056160000 | 0.58 | 0 | healthy_plasma | 29.025760115 | healthy_plasma |
| D1000S170 | healthy_plasma | 0.1922393333 | 0.58 | 0 | healthy_plasma | 50.423760483 | healthy_plasma |
| D1000S171 | healthy_plasma | 0.3393206666 | 0.58 | 0 | healthy_plasma | 56.083595841 | healthy_plasma |
| D1000S172 | healthy_plasma | 0.1749246666 | 0.58 | 0 | healthy_plasma | 34.784933112 | healthy_plasma |
| D1000S173 | healthy_plasma | 0.2737906666 | 0.58 | 0 | healthy_plasma | 95.053106562 | healthy_plasma |
| D1000S174 | healthy_plasma | 0.2010913333 | 0.58 | 0 | healthy_plasma | 76.176577981 | healthy_plasma |
| D1000S175 | healthy_plasma | 0.2696733333 | 0.58 | 0 | healthy_plasma | 67.946150045 | healthy_plasma |
| D1525S051 | healthy_plasma | 0.2974599999 | 0.58 | 0 | healthy_plasma | 92.073051423 | healthy_plasma |
| D1525S052 | healthy_plasma | 0.5164359999 | 0.58 | 0.6918 | healthy_plasma | 108.86228015 | healthy_plasma |
| D1525S053 | healthy_plasma | 0.1232346666 | 0.58 | 0 | healthy_plasma | 120.39097523 | healthy_plasma |
| D1525S054 | healthy_plasma | 0.0878233333 | 0.58 | 0 | healthy_plasma | 90.816646741 | healthy_plasma |
| D1525S055 | healthy_plasma | 0.3892740000 | 0.58 | 0 | healthy_plasma | 89.578088918 | healthy_plasma |
| D1525S085 | healthy_plasma | 0.1527213333 | 0.58 | 0 | healthy_plasma | 63.804566552 | healthy_plasma |
| D1525S086 | healthy_plasma | 0.4360846666 | 0.58 | 0.014 | healthy_plasma | 50.842149304 | healthy_plasma |
| D1525S087 | healthy_plasma | 0.4376580000 | 0.58 | 0.0126 | healthy_plasma | 82.106353084 | healthy_plasma |
| D1525S088 | healthy_plasma | 0.3235759999 | 0.58 | 0 | healthy_plasma | 66.945300015 | healthy_plasma |
| D1525S089 | healthy_plasma | 0.2351926666 | 0.58 | 0 | healthy_plasma | 55.926292225 | healthy_plasma |
| D1525S090 | healthy_plasma | 0.2500233333 | 0.58 | 0 | healthy_plasma | 80.340378342 | healthy_plasma |
| D1525S091 | healthy_plasma | 0.3224519999 | 0.58 | 0 | healthy_plasma | 97.491344876 | healthy_plasma |
| D1357S121 | healthy_plasma | 0.3769840000 | 0.58 | 0 | healthy_plasma | 55.971272250 | healthy_plasma |
| D1357S122 | healthy_plasma | 0.2330140000 | 0.58 | 0 | healthy_plasma | 36.806504192 | healthy_plasma |
| D1357S123 | healthy_plasma | 0.1691239999 | 0.58 | 0 | healthy_plasma | 35.049071105 | healthy_plasma |
| D1357S124 | healthy_plasma | 0.2819279999 | 0.58 | 0 | healthy_plasma | 58.707238724 | healthy_plasma |
| D1357S125 | healthy_plasma | 0.3930233333 | 0.58 | 0 | healthy_plasma | 62.809474540 | healthy_plasma |
| D1357S129 | healthy_plasma | 0.2678473333 | 0.58 | 0 | healthy_plasma | 48.965451737 | healthy_plasma |
| D1357S130 | healthy_plasma | 0.2950846666 | 0.58 | 0 | healthy_plasma | 47.622546728 | healthy_plasma |
| D1357S131 | healthy_plasma | 0.1517519999 | 0.58 | 0 | healthy_plasma | 58.966765475 | healthy_plasma |
| D1357S132 | healthy_plasma | 0.0935826666 | 0.58 | 0 | healthy_plasma | 47.032430761 | healthy_plasma |
| D1357S133 | healthy_plasma | 0.3563553333 | 0.58 | 0 | healthy_plasma | 105.29473272 | healthy_plasma |
| D1357S134 | healthy_plasma | 0.3352393333 | 0.58 | 0 | healthy_plasma | 49.170611073 | healthy_plasma |
| D1357S137 | healthy_plasma | 0.3935886666 | 0.58 | 0 | healthy_plasma | 64.667229393 | healthy_plasma |
| D1357S138 | healthy_plasma | 0.3539339999 | 0.58 | 0 | healthy_plasma | 43.336757774 | healthy_plasma |
| D1357S139 | healthy_plasma | 0.2684286666 | 0.58 | 0 | healthy_plasma | 45.146373444 | healthy_plasma |
| D1357S140 | healthy_plasma | 0.3373833333 | 0.58 | 0 | healthy_plasma | 54.903057870 | healthy_plasma |
| D1357S144 | healthy_plasma | 0.4018066666 | 0.58 | 0.0002 | healthy_plasma | 82.575437010 | healthy_plasma |
| D1357S145 | healthy_plasma | 0.2837553333 | 0.58 | 0 | healthy_plasma | 41.238589309 | healthy_plasma |
| D1357S146 | healthy_plasma | 0.2447793333 | 0.58 | 0 | healthy_plasma | 41.008129289 | healthy_plasma |
| D1357S147 | healthy_plasma | 0.2894326666 | 0.58 | 0 | healthy_plasma | 62.393633356 | healthy_plasma |
| D1000S100 | lung_cancer_plasma_M+ | 0.3615346666 | 0.58 | 0 | healthy_plasma | 110.01783664 | healthy_plasma |
| D1000S101 | lung_cancer_plasma_M+ | 0.5941646666 | 0.58 | 0.9992 | lung_cancer_plasma_M+ | 326.45289540 | lung_cancer_plasma_M+ |
| D1000S102 | lung_cancer_plasma_M+ | 0.2686686666 | 0.58 | 0 | healthy_plasma | 407.42474655 | lung_cancer_plasma_M+ |
| D1000S103 | lung_cancer_plasma_M+ | 0.3604786666 | 0.58 | 0 | healthy_plasma | 166.38760201 | lung_cancer_plasma_M+ |
| D1000S140 | lung_cancer_plasma_M+ | 0.5692159999 | 0.58 | 0.9892 | healthy_plasma | 97.159596483 | healthy_plasma |
| D1000S141 | lung_cancer_plasma_M+ | 0.6836719999 | 0.58 | 1 | lung_cancer_plasma_M+ | 243.70110663 | lung_cancer_plasma_M+ |
| D1000S142 | lung_cancer_plasma_M+ | 0.3488219999 | 0.58 | 0 | healthy_plasma | 201.30864394 | lung_cancer_plasma_M+ |
| D1000S143 | lung_cancer_plasma_M+ | 0.6465580000 | 0.58 | 1 | lung_cancer_plasma_M+ | 82.096115973 | lung_cancer_plasma_M+ |

**Table S21.** Cancer prediction and sample labelling with the 2-step classification integrating CNA analysis - healthy vs OVC M+ model.

| sample | biological_class | probaCancer | probaCancer_Threshold | predCancer | predict_biological_class | Gzscore | predict_biological_class_with_CNA |
| --- | --- | --- | --- | --- | --- | --- | --- |
| D1000S071 | healthy_plasma | 0.0066573333 | 0.09 | 0 | healthy_plasma | 44.536933544 | healthy_plasma |
| D1000S072 | healthy_plasma | 0.0078733333 | 0.09 | 0 | healthy_plasma | 47.726078345 | healthy_plasma |
| D1000S073 | healthy_plasma | 0.0052586666 | 0.09 | 0 | healthy_plasma | 32.670596164 | healthy_plasma |
| D1000S074 | healthy_plasma | 0.0002033333 | 0.09 | 0 | healthy_plasma | 27.138499610 | healthy_plasma |
| D1000S075 | healthy_plasma | 0.0353959999 | 0.09 | 0 | healthy_plasma | 35.486610691 | healthy_plasma |
| D1000S076 | healthy_plasma | 0.0011033333 | 0.09 | 0 | healthy_plasma | 50.995612621 | healthy_plasma |
| D1000S077 | healthy_plasma | 0.0402546666 | 0.09 | 0 | healthy_plasma | 34.430682458 | healthy_plasma |
| D1000S078 | healthy_plasma | 0.0009759999 | 0.09 | 0 | healthy_plasma | 33.104277668 | healthy_plasma |
| D1000S079 | healthy_plasma | 0.0048760000 | 0.09 | 0 | healthy_plasma | 67.809379568 | healthy_plasma |
| D1000S118 | healthy_plasma | 0.0190453333 | 0.09 | 0 | healthy_plasma | 36.519128297 | healthy_plasma |
| D1000S119 | healthy_plasma | 0.0092740000 | 0.09 | 0 | healthy_plasma | 74.146627837 | healthy_plasma |
| D1000S120 | healthy_plasma | 0.0382059999 | 0.09 | 0 | healthy_plasma | 31.505984270 | healthy_plasma |
| D1000S121 | healthy_plasma | 0.0460526666 | 0.09 | 0 | healthy_plasma | 79.182843946 | healthy_plasma |
| D1000S122 | healthy_plasma | 0.0213519999 | 0.09 | 0 | healthy_plasma | 90.727250244 | healthy_plasma |
| D1000S123 | healthy_plasma | 0.0277619999 | 0.09 | 0 | healthy_plasma | 54.181549949 | healthy_plasma |
| D1000S124 | healthy_plasma | 0.0140353333 | 0.09 | 0 | healthy_plasma | 40.345100692 | healthy_plasma |
| D1000S125 | healthy_plasma | 0.0117913333 | 0.09 | 0 | healthy_plasma | 53.860911314 | healthy_plasma |
| D1000S126 | healthy_plasma | 0.0092246666 | 0.09 | 0 | healthy_plasma | 62.960560967 | healthy_plasma |
| D1000S127 | healthy_plasma | 0.0347586666 | 0.09 | 0 | healthy_plasma | 33.434057480 | healthy_plasma |
| D1000S166 | healthy_plasma | 0.0138453333 | 0.09 | 0 | healthy_plasma | 53.749015624 | healthy_plasma |
| D1000S167 | healthy_plasma | 0.0014613333 | 0.09 | 0 | healthy_plasma | 27.021584243 | healthy_plasma |
| D1000S168 | healthy_plasma | 0.0268539999 | 0.09 | 0 | healthy_plasma | 54.263255926 | healthy_plasma |
| D1000S169 | healthy_plasma | 0.0079800000 | 0.09 | 0 | healthy_plasma | 29.025760115 | healthy_plasma |
| D1000S170 | healthy_plasma | 0.0059426666 | 0.09 | 0 | healthy_plasma | 50.423760483 | healthy_plasma |
| D1000S171 | healthy_plasma | 0.0204926666 | 0.09 | 0 | healthy_plasma | 56.083595841 | healthy_plasma |
| D1000S172 | healthy_plasma | 0.0076726666 | 0.09 | 0 | healthy_plasma | 34.784933112 | healthy_plasma |
| D1000S173 | healthy_plasma | 0.0068753333 | 0.09 | 0 | healthy_plasma | 95.053106562 | healthy_plasma |
| D1000S174 | healthy_plasma | 0.0054613333 | 0.09 | 0 | healthy_plasma | 76.176577981 | healthy_plasma |
| D1000S175 | healthy_plasma | 0.0033206666 | 0.09 | 0 | healthy_plasma | 67.946150045 | healthy_plasma |
| D1525S051 | healthy_plasma | 0.0098953333 | 0.09 | 0 | healthy_plasma | 92.073051423 | healthy_plasma |
| D1525S052 | healthy_plasma | 0.0561173333 | 0.09 | 0 | healthy_plasma | 108.86228015 | healthy_plasma |
| D1525S053 | healthy_plasma | 0.0073880000 | 0.09 | 0 | healthy_plasma | 120.39097523 | healthy_plasma |
| D1525S054 | healthy_plasma | 0.0092413333 | 0.09 | 0 | healthy_plasma | 90.816646741 | healthy_plasma |
| D1525S055 | healthy_plasma | 0.0727559999 | 0.09 | 0 | healthy_plasma | 89.578088918 | healthy_plasma |
| D1525S085 | healthy_plasma | 0.0107033333 | 0.09 | 0 | healthy_plasma | 63.804566552 | healthy_plasma |
| D1525S086 | healthy_plasma | 0.01241 | 0.09 | 0 | healthy_plasma | 50.842149304 | healthy_plasma |
| D1525S087 | healthy_plasma | 0.01259 | 0.09 | 0 | healthy_plasma | 82.106353084 | healthy_plasma |
| D1525S088 | healthy_plasma | 0.0124513333 | 0.09 | 0 | healthy_plasma | 66.945300015 | healthy_plasma |
| D1525S089 | healthy_plasma | 0.0092300000 | 0.09 | 0 | healthy_plasma | 55.926292225 | healthy_plasma |
| D1525S090 | healthy_plasma | 0.0039126666 | 0.09 | 0 | healthy_plasma | 80.340378342 | healthy_plasma |
| D1525S091 | healthy_plasma | 0.0287573333 | 0.09 | 0 | healthy_plasma | 97.491344876 | healthy_plasma |
| D1357S121 | healthy_plasma | 0.0407153333 | 0.09 | 0 | healthy_plasma | 55.971272250 | healthy_plasma |
| D1357S122 | healthy_plasma | 0.0039000000 | 0.09 | 0 | healthy_plasma | 36.806504192 | healthy_plasma |
| D1357S123 | healthy_plasma | 0.0202379999 | 0.09 | 0 | healthy_plasma | 35.049071105 | healthy_plasma |
| D1357S124 | healthy_plasma | 0.0247573333 | 0.09 | 0 | healthy_plasma | 58.707238724 | healthy_plasma |
| D1357S125 | healthy_plasma | 0.0020106666 | 0.09 | 0 | healthy_plasma | 62.809474540 | healthy_plasma |
| D1357S129 | healthy_plasma | 0.0100486666 | 0.09 | 0 | healthy_plasma | 48.965451737 | healthy_plasma |
| D1357S130 | healthy_plasma | 0.0102053333 | 0.09 | 0 | healthy_plasma | 47.622546728 | healthy_plasma |
| D1357S131 | healthy_plasma | 0.0033760000 | 0.09 | 0 | healthy_plasma | 58.966765475 | healthy_plasma |
| D1357S132 | healthy_plasma | 0.0028033333 | 0.09 | 0 | healthy_plasma | 47.032430761 | healthy_plasma |
| D1357S133 | healthy_plasma | 0.0337826666 | 0.09 | 0 | healthy_plasma | 105.29473272 | healthy_plasma |
| D1357S134 | healthy_plasma | 0.0829446666 | 0.09 | 0 | healthy_plasma | 49.170611073 | healthy_plasma |
| D1357S137 | healthy_plasma | 0.0653899999 | 0.09 | 0 | healthy_plasma | 64.667229393 | healthy_plasma |
| D1357S138 | healthy_plasma | 0.0196693333 | 0.09 | 0 | healthy_plasma | 43.336757774 | healthy_plasma |
| D1357S139 | healthy_plasma | 0.0014033333 | 0.09 | 0 | healthy_plasma | 45.146373444 | healthy_plasma |
| D1357S140 | healthy_plasma | 0.0183493333 | 0.09 | 0 | healthy_plasma | 54.903057870 | healthy_plasma |
| D1357S144 | healthy_plasma | 0.0304199999 | 0.09 | 0 | healthy_plasma | 82.575437010 | healthy_plasma |
| D1357S145 | healthy_plasma | 0.0072926666 | 0.09 | 0 | healthy_plasma | 41.238589309 | healthy_plasma |
| D1357S146 | healthy_plasma | 0.0157953333 | 0.09 | 0 | healthy_plasma | 41.008129289 | healthy_plasma |
| D1357S147 | healthy_plasma | 0.0739339999 | 0.09 | 0 | healthy_plasma | 62.393633356 | healthy_plasma |
| D1000S054 | ovarian_cancer_plasma_M+ | 0.5677386666 | 0.09 | 0.9896 | ovarian_cancer_plasma_M+ | 266.65749248 | ovarian_cancer_plasma_M+ |
| D1000S055 | ovarian_cancer_plasma_M+ | 0.511266 | 0.09 | 0.624 | ovarian_cancer_plasma_M+ | 1057.3944790 | ovarian_cancer_plasma_M+ |
| D1000S056 | ovarian_cancer_plasma_M+ | 0.3664306666 | 0.09 | 0 | ovarian_cancer_plasma_M+ | 409.69650538 | ovarian_cancer_plasma_M+ |
| D1000S058 | ovarian_cancer_plasma_M+ | 0.3917333333 | 0.09 | 0 | ovarian_cancer_plasma_M+ | 212.16992364 | ovarian_cancer_plasma_M+ |
| D1000S059 | ovarian_cancer_plasma_M+ | 0.3018146666 | 0.09 | 0 | ovarian_cancer_plasma_M+ | 208.12283940 | ovarian_cancer_plasma_M+ |
| D1000S060 | ovarian_cancer_plasma_M+ | 0.2584379999 | 0.09 | 0 | ovarian_cancer_plasma_M+ | 83.477241197 | ovarian_cancer_plasma_M+ |
| D1000S061 | ovarian_cancer_plasma_M+ | 0.2216273333 | 0.09 | 0 | ovarian_cancer_plasma_M+ | 100.53377480 | ovarian_cancer_plasma_M+ |
| D1000S104 | ovarian_cancer_plasma_M+ | 0.3853546666 | 0.09 | 0 | ovarian_cancer_plasma_M+ | 204.04564294 | ovarian_cancer_plasma_M+ |
| D1000S105 | ovarian_cancer_plasma_M+ | 0.2081980000 | 0.09 | 0 | ovarian_cancer_plasma_M+ | 69.904523685 | ovarian_cancer_plasma_M+ |
| D1000S106 | ovarian_cancer_plasma_M+ | 0.0634319999 | 0.09 | 0 | healthy_plasma | 86.596194599 | healthy_plasma |
| D1000S107 | ovarian_cancer_plasma_M+ | 0.060984 | 0.09 | 0 | healthy_plasma | 86.399538892 | healthy_plasma |
| D1000S108 | ovarian_cancer_plasma_M+ | 0.1498459999 | 0.09 | 0 | ovarian_cancer_plasma_M+ | 90.898815070 | ovarian_cancer_plasma_M+ |
| D1000S109 | ovarian_cancer_plasma_M+ | 0.1417539999 | 0.09 | 0 | ovarian_cancer_plasma_M+ | 100.55161742 | ovarian_cancer_plasma_M+ |
| D1000S110 | ovarian_cancer_plasma_M+ | 0.1087360000 | 0.09 | 0 | ovarian_cancer_plasma_M+ | 89.224867737 | ovarian_cancer_plasma_M+ |
| D1000S144 | ovarian_cancer_plasma_M+ | 0.3491579999 | 0.09 | 0 | ovarian_cancer_plasma_M+ | 54.922757680 | ovarian_cancer_plasma_M+ |
| D1000S145 | ovarian_cancer_plasma_M+ | 0.0529086666 | 0.09 | 0 | healthy_plasma | 250.53250045 | ovarian_cancer_plasma_M+ |
| D1000S146 | ovarian_cancer_plasma_M+ | 0.0698639999 | 0.09 | 0 | healthy_plasma | 246.54914091 | ovarian_cancer_plasma_M+ |
| D1000S147 | ovarian_cancer_plasma_M+ | 0.2437366666 | 0.09 | 0 | ovarian_cancer_plasma_M+ | 745.01491059 | ovarian_cancer_plasma_M+ |
| D1000S148 | ovarian_cancer_plasma_M+ | 0.0703366666 | 0.09 | 0 | healthy_plasma | 112.24838179 | healthy_plasma |
| D1000S149 | ovarian_cancer_plasma_M+ | 0.1336999999 | 0.09 | 0 | ovarian_cancer_plasma_M+ | 228.33169876 | ovarian_cancer_plasma_M+ |
| D1000S150 | ovarian_cancer_plasma_M+ | 0.1046600000 | 0.09 | 0 | ovarian_cancer_plasma_M+ | 211.79872567 | ovarian_cancer_plasma_M+ |
| D1000S151 | ovarian_cancer_plasma_M+ | 0.5399833333 | 0.09 | 0.906 | ovarian_cancer_plasma_M+ | 211.49490452 | ovarian_cancer_plasma_M+ |
| D1000S152 | ovarian_cancer_plasma_M+ | 0.2033306666 | 0.09 | 0 | ovarian_cancer_plasma_M+ | 140.99497567 | ovarian_cancer_plasma_M+ |
| D1000S153 | ovarian_cancer_plasma_M+ | 0.0930613333 | 0.09 | 0 | ovarian_cancer_plasma_M+ | 100.44667712 | ovarian_cancer_plasma_M+ |
| D1000S192 | ovarian_cancer_plasma_M+ | 0.4540926666 | 0.09 | 0.0464 | ovarian_cancer_plasma_M+ | 464.62438410 | ovarian_cancer_plasma_M+ |
| D1000S193 | ovarian_cancer_plasma_M+ | 0.1371866666 | 0.09 | 0 | ovarian_cancer_plasma_M+ | 40.958876476 | ovarian_cancer_plasma_M+ |
| D1000S194 | ovarian_cancer_plasma_M+ | 0.0997980000 | 0.09 | 0 | ovarian_cancer_plasma_M+ | 79.887609256 | ovarian_cancer_plasma_M+ |
| D1000S195 | ovarian_cancer_plasma_M+ | 0.0756706666 | 0.09 | 0 | healthy_plasma | 52.170281578 | healthy_plasma |
| D1000S196 | ovarian_cancer_plasma_M+ | 0.2911133333 | 0.09 | 0 | ovarian_cancer_plasma_M+ | 32.481220672 | ovarian_cancer_plasma_M+ |
| D1000S197 | ovarian_cancer_plasma_M+ | 0.1052020000 | 0.09 | 0 | ovarian_cancer_plasma_M+ | 429.19754464 | ovarian_cancer_plasma_M+ |
| D1000S198 | ovarian_cancer_plasma_M+ | 0.126574 | 0.09 | 0 | ovarian_cancer_plasma_M+ | 95.080040893 | ovarian_cancer_plasma_M+ |
| D1000S199 | ovarian_cancer_plasma_M+ | 0.0697199999 | 0.09 | 0 | healthy_plasma | 90.850580161 | healthy_plasma |
| D1000S200 | ovarian_cancer_plasma_M+ | 0.3166219999 | 0.09 | 0 | ovarian_cancer_plasma_M+ | 28.280664380 | ovarian_cancer_plasma_M+ |
| D1000S201 | ovarian_cancer_plasma_M+ | 0.0865486666 | 0.09 | 0 | healthy_plasma | 103.20182411 | healthy_plasma |
| D1000S202 | ovarian_cancer_plasma_M+ | 0.2275780000 | 0.09 | 0 | ovarian_cancer_plasma_M+ | 58.149410968 | ovarian_cancer_plasma_M+ |
| D1000S203 | ovarian_cancer_plasma_M+ | 0.2220733333 | 0.09 | 0 | ovarian_cancer_plasma_M+ | 695.07428064 | ovarian_cancer_plasma_M+ |
| D1000S204 | ovarian_cancer_plasma_M+ | 0.1456159999 | 0.09 | 0 | ovarian_cancer_plasma_M+ | 31.420115707 | ovarian_cancer_plasma_M+ |

**Table S22.**
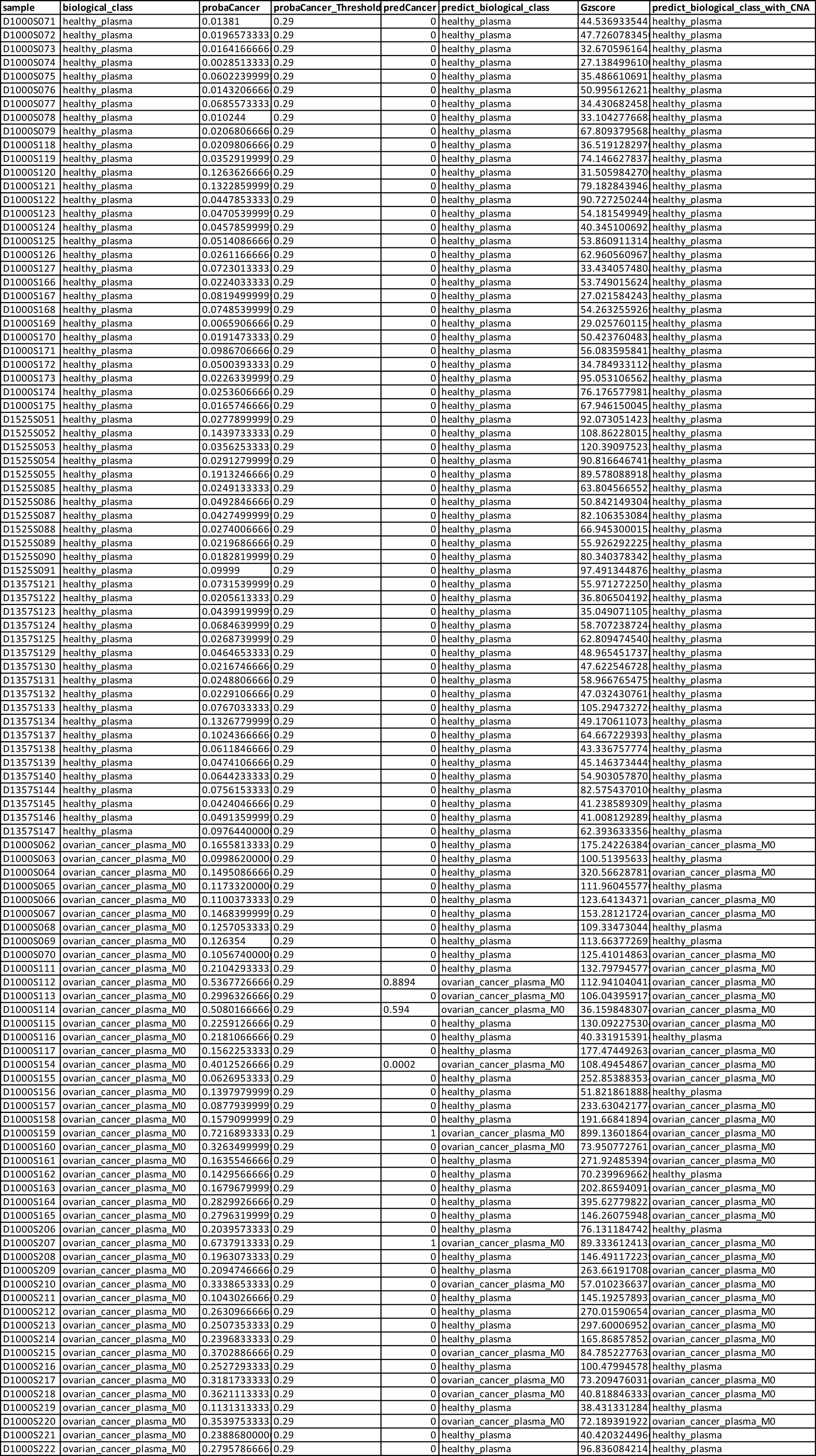
Cancer prediction and sample labelling with the 2-step classification integrating CNA analysis - healthy vs OVC M0 model.

